# Integrated Genomic and Immune Profiling Reveals PIK3CA-Mutant HPV16-Positive Cervical Cancer as a Clinically Aggressive and Immunologically Distinct Subtype in Patients from Eastern India

**DOI:** 10.64898/2026.10.27.26366457

**Authors:** Abhisikta Ghosh, Arnab Ghosh, Abarna Sinha, Sonia Mathai, Jaydip Bhaumik, Asima Mukhopadhyay, Sreyosi Majumder, Arindam Maitra, Nidhan K. Biswas, Koushik Mondal, Partha P. Majumder, Sharmila Sengupta

**Author notes:** To whom correspondence and reprint requests should be addressed: *Present Address: **Sharmila Sengupta, PhD*** Saroj Gupta Cancer Centre & Research Institute, Kolkata, West Bengal, India. Present Address: National Centre for Biological Sciences (NCBS), Tata Institute of Fundamental Research (TIFR), Bengaluru, Karnataka, India.

## Abstract

**Background:** HPV16-driven cervical cancer (CaCx) is a leading cause of cancer-related mortality among Indian women. Identifying population-specific genomic and immune determinants may improve therapeutic strategies.

**Methods:** We performed whole-exome sequencing on 49 HPV16-positive cervical tumors and matched blood samples. Available RNA-seq data from 29 out of 49 tumors were integrated, to identify recurrent mutations, altered pathways, and immune microenvironment features.

**Results:** We identified 6,462 somatic mutations enriched in cancer-related pathways, particularly PI3K-Akt signaling. *PIK3CA* was the most recurrently mutated gene (42%), significantly higher frequency (p < 0.04) than in HPV16-positive TCGA-CESC tumors (26.21%). Unlike TCGA-CESC, where *PIK3CA* mutations were associated with early-stage disease, Indian *PIK3CA*-mutant tumors were enriched in advanced-stage disease. These tumors exhibited lower viral loads, higher tumor mutation burden, and more aggressive molecular phenotypes than *PIK3CA*-wild tumors. CIBERSORT-based immune profiling showed comparable immune-cell infiltration, including CD8+ T cells, between *PIK3CA*-mutant tumors and normal tissues, whereas *PIK3CA*-wild tumors showed significantly enhanced infiltration, compared to normal tissues. Both tumor subtypes portrayed mature tertiary lymphoid structures at higher levels than normal tissues. Compared to normal tissues, *CD274* (PD-L1) expression was elevated significantly in both subtypes, while *PDCD1* (PD-1) upregulation and PD-1/PD-L1 transcriptional coupling at significant levels, occurred exclusively in *PIK3CA*-mutant tumors. The CCL3-CCR1 axis was selectively activated in *PIK3CA*-mutant tumors and correlated with cytotoxic, interferon-response, myeloid, and immune checkpoint markers, indicating an immunosuppressive microenvironment.

**Conclusion:** *PIK3CA* mutations define a molecularly aggressive, immunologically distinct subtype of HPV16-positive CaCx in Indian patients, underscoring their potential for targeted therapeutic interventions.

## Introduction

Cervical cancer (CaCx) continues to be a major public health concern in low- and middle-income countries (LMICs), with India and China together accounting for about one-third of the global disease burden. According to Global Cancer Observatory (GLOBOCAN) 2022 report (https://gco.iarc.who.int/media/globocan/factsheets/populations/356-india-fact-sheet.pdf), CaCx continues to be the second most common cancer among Indian women both in terms of incidence and mortality. Late presentation of affected women in clinics, lack of an organized cervical screening program involving HPV testing and absence of awareness for HPV vaccination program at the National level (https://www.uicc.org/news/globocan-2022-latest-global-cancer-data-shows-rising-incidence-and-stark-inequities) are likely to be the major contributing factors. Almost all the CaCx cases develop under the impact of persistent infection with high-risk Human PappilomaVirus (HR-HPV), especially HPV16 (1,2). Although HPV infection plays a crucial role in tumor pathogenesis, but it is not sufficient. Several other cofactors like host genetic and epigenetic alterations also play important roles in disease progression.

The role of somatic mutations and copy number alterations in CaCx pathogenesis has been well documented (3,4) through candidate gene or single-gene mutation studies. More recently, several studies (5–7) have provided comprehensive insights into the genomic landscape of CaCx, relevant not only for disease pathogenesis, but also of clinical importance. Such findings have contributed immensely to the development of novel therapeutic approaches, such as targeted therapy and immunotherapy, which, in many cancer types, hold the promise for improved efficacy by selectively modulating cancer or immune cells, while minimizing toxicity and drug resistance. Besides, differential selection pressure and environmental conditions across different geographical regions, have resulted in population-specific genetic architecture and immune response. These variations influence the frequency and impact of genetic alterations, underscoring the importance of conducting population-specific genomic studies. Although there are several Whole Exome Sequencing (WES) studies on CaCx, in various populations, such studies from India are limited (8,9). High genetic diversity among Indian populations is well established, and as genetic variations can influence disease susceptibility and therapeutic response, it is important to investigate molecular drivers of CaCx within Indian populations.

In this study, we aimed to characterise the mutational landscape of treatment-naive, HPV16-positive cervical tumors of female patients from Eastern India. We employed WES to identify the driver mutations associated with the disease and potential actionable genomic markers. We identified Phosphatidylinositol 4,5-bisphosphate 3-kinase catalytic subunit alpha (PIK3CA) gene with oncogenic mutations to be the major driver for HPV16-related CaCx pathogenesis, among Indian patients. The majority of such PIK3CA mutations are classified as actionable alterations and the U.S. Food and Drug Administration (FDA) has approved targeted therapies against the mutations in PIK3CA gene in other cancer types, such as breast cancer. To understand more deeply the clinical and biological relevance of these findings, we compared PIK3CA-mutant and -wild tumors in terms of clinical parameters, gene expression profile-based cancer phenotypes, and viral factors. We also explored the tumor immune microenvironment (TIME) of the PIK3CA-mutant and -wild tumors, in comparison with healthy individuals, considering the (i) Tumor Infiltrating Immune cells (TIICs), (ii) immune active component through Tertiary Lymphoid Structures (TLS) enrichment and maturation, and (iii) the immune suppressive axis, mediated by Programmed Death-1 (PD-1) and Programmed Death-Ligand 1 (PD-L1) axis. Through such analysis, we aimed to define clinically relevant molecular subtypes of HPV16-positive CaCx and predict their potential responsiveness to immunotherapeutic interventions.

## Methods

### Patient recruitment, sample collection and ethical approval

This study was approved by the Ethics Committees of Tata Medical Center (TMC), Kolkata, and National Institute of Biomedical Genomics (NIBMG), Kalyani. With written informed consent, we collected CaCx primary tissue biopsies and matched blood samples from treatment-naive married patients (age range: 36-78 years; median age: 58 years) who were diagnosed with the disease (confirmed as malignant by histopathology) at TMC. Similarly, we collected, non-malignant (confirmed by histopathology) cervical tissue biopsies from married women (age range: 28-50 years; median age: 43 years) who underwent hysterectomy for reasons other than cancer from the College of Medicine and JNM Hospital, Kalyani. We also collected relevant clinical data from each woman recruited into this study and provided a summary of this in **Supplementary Table S1**.

### Sample processing, Whole-exome sequence (WES) data generation and analysis

We isolated DNA from each biopsied tissue and blood sample, using the QIAamp DNA mini kit (Cat No: 51304) and QIASYMPHONY DSP DNA Midi Kit (Qiagen, Cat No-937255), respectively. We screened DNA isolated from cervical tissue samples of patients and healthy individuals for the presence of HPV, using DNA-based HPV-type specific PCR primers as mentioned in our earlier studies (10). As majority of the HPV infected CaCx patients (about 72.22%) carried HPV16 genotype, therefore, we focused our attention on HPV16-positive samples only for subsequent analysis. We performed WES for HPV16-positive CaCx tissue and paired blood samples (n=49), using TruSeq Exome Library Prep kit (Illumina) followed by sequencing in Nova-Seq 6000 (Illumina) at 90X depth. We have provided the details of sequencing quality in **Supplementary Table S2** and described the methods in detail in **Supplementary Methods (Section S1)**

We analysed the sequenced data to identify the somatic alterations in HPV16-positive CaCx patients, following the approach previously published by our group (11); details provided in **Supplementary Methods (Sections S2 and S3)**. Briefly, we used a suite of packages that included FastQC for quality control (QC), Burrows-Wheeler Aligner-Maximal Exact Match (BWA-MEM-0.7.17) for alignment (reference genome version used for alignment - “hs37d5”), PICARD-2.17.11 (https://github.com/broadinstitute/picard) and Genomic Analysis Tool Kit (GATK-3.8) for post-alignment processing. For estimating the coverage and median insert size, we used the Qualimap-2.2.1 tool.

Thereafter, we performed somatic variant calling using multiple variant callers: GATK-4 Mutect2, Strelka-2, VarScan2 and MuSE (for Single Nucleotide Variants or SNV detection) and GATK-4 Mutect2, Strelka2, VarScan2 (for Insertion and Deletion or INDEL detection). For further analysis, we accepted only those variants that were detected by at least 2 out of the 4 above-mentioned variant callers. Subsequently, we performed the filtering of the variants for sequencing artifacts such as low frequency, strand-bias, OXO-G, low complexity regions, etc, and polymorphic genomic loci in GenomeAsia100K (detailed in **Supplementary Methods, Section S3**), followed by the generation of detailed functional annotations for the filtered set of variants using Oncotator-1.9.9.0.

We determined the functional impact of the SNVs on the protein functionality using 3 mutation prediction algorithms, such as Mutation Assessor, Polyphen (Polyphen2_HVAR and Polyphen2_HDIV), and Sorting Intolerant from Tolerant (SIFT). The SNVs were categorized as deleterious, damaging, putatively damaging, and benign by these algorithms. For further analysis, we considered only those SNVs that were predicted to be deleterious or damaging by any 2 out of the 3 prediction algorithms. INDELs, on the other hand, were considered pathogenic.

To validate the identified somatic mutations, we manually inspected the sequencing reads corresponding to the genomic regions through Integrative Genomics Viewer (IGV) (12), using hg19 as the reference genome. Binary Alignment Map (BAM) files from tumor tissue samples, their corresponding paired blood samples, as well as from tissues of healthy individuals were examined to verify the presence or absence of identified variants. Variants were considered as tumor-specific when detected only in tumor tissue samples but absent from corresponding blood samples and healthy individuals.

### Identification of actionable alterations using precision oncology resources

We determined the clinical actionability of individual genomic variants using a precision oncology and FDA-recognised Human Genetic Variant Database called OncoKB-version 4.15 on 21.3.2024 (www.oncokb.org; **RRID**: SCR_014782) (13,14). OncoKB annotates the biological and oncogenic impact as well as the prognostic and predictive significance of somatic mutation alterations in cancer. It provides evidence-based information on drug response for specific molecular alterations (Level of evidence-version V2). We provided the details of the level of evidence in **Supplementary Methods (Section S4)**.

### Whole-transcriptome sequence data generation and analysis

For this study, we utilised a subset of whole-transcriptome sequencing data generated and published by our group (15,16). Briefly, we isolated total RNA from HPV16-positive CaCx tissue and normal cervical tissue biopsies using Qiagen RNeasy Mini Kit (Cat. No-74104) and performed Whole transcriptome sequencing (see **Supplementary Methods Section S5)**. We analysed the RNA-seq data as per the GDC pipeline described in detail in our previous publications (11,15,16) and **Supplementary Methods (Section S6)**, employing a suite of packages including FastQC-0.11.7, STAR aligner (STAR-2.6.0c, **RRID:** SCR_004463) and HTSeq (HTSeq-0.11.0) for QC, alignment, and count generation, respectively. We performed differential gene expression analysis between the comparison groups using DESeq2 and generated the TPM (Transcripts per million) values using TPMCalculator tool. We have provided the sequencing quality for RNA-seq samples used in this study (n=29) in Supplementary Table S3.

### Identification of Tertiary Lymphoid Structures (TLS) from bulk RNA-seq data based on pre-existing classical TLS Signature

In order to assess the presence of TLS in HPV16-positive CaCx patients and healthy individuals, we curated a panel of 75 TLS-associated genes from the literature (17–22). These genes were involved in the formation and maturation of TLS, as described in **Supplementary Table S4**. To ensure reliable analysis, we performed quality filtering and retained only those genes with non-zero median expression (TPM) that were detectable in at least 20% of our study cohort (i.e, both CaCx patients and healthy individuals). Then we calculated the TLS Enrichment Score for each sample using the geometric mean of the expression (TPM) of the selected gene signatures, as described by Mughal et. al (23). We subsequently stratified the samples into TLS-High, TLS-Low, and TLS-intermediate based on tertile cutoffs (33rd and 66th percentiles) of the median TLS enrichment score i.e, TLS-Low if the TLS median Enrichment Score is ≤ 3.2, TLS-Intermediate if the score is >3.2 and <6.5, and TLS-High if the score ≥ 6.5.

### TLS stratification based on the maturity of TLS in healthy individuals and HPV16-positive CaCx patients

The TLS gene signatures were further categorised into early, intermediate, and mature TLS signatures according to their roles during different stages of TLS development (**Supplementary Table S4**). For each sample, we calculated a stage-specific TLS enrichment score using the geometric mean of the expression (TPM) of the stage-specific genes (23). Further, to enable relative comparison across samples irrespective of absolute gene expression scales, we compared the TLS scores across each sample by computing z-scores in R (https://search.r-project.org/CRAN/refmans/hacksig/html/hack_sig.html) and assigned each sample a dominant TLS stage (Early, Intermediate and Mature) based on the highest z-score among the three stages.

### Analysis of Immune cell composition in the Tumor microenvironment from bulk RNA-seq data

For estimating the immune cell abundance in normal cervical and CaCx tissues, we employed Cell-type Identification by Estimating Relative Subsets of RNA Transcripts (CIBERSORT)-absolute deconvolution algorithm with default parameters. We utilised gene expression data (TPM values) to generate a proportion matrix of 22 Tumor Infiltrating Immune Cells (TIICs), in these tissues (24).

### Estimation of HPV16 load, physical status and E7 expression in the HPV16-positive CaCx tissues

We estimated the viral load (defined as the number of copies of viral genome per hundred nanograms of genomic DNA and represented as natural log transformed) and viral physical status (episomal, concomitant, or integrated) in HPV16-positive CaCx tissues, employing TaqMan-based qRT-PCR, as described in our earlier studies (15,25). We calculated relative HPV16 E7 expression in CaCx tissues using Taqman based qRT-PCR, with 18s rRNA as internal control. The detailed protocol is described in our earlier publication (26). We provided the amplification curves from qRT-PCR in **Supplementary Figure S1.**

### Bioinformatics and Statistical analysis: Network and pathway analysis, Data visualisation

We generated heatmaps using the R pheatmap package (https://cran.r-project.org/web/packages/pheatmap/pheatmap.pdf; **RRID**: SCR_016418); lollipop plot using Mutation Mapper tool available in cBioportal (https://www.cbioportal.org/; **RRID**: SCR_014555) (27); box plots representing the distribution across different groups, using R programming. We performed the statistical analysis in R and GraphPad Prism 9.0 (**RRID:** SCR_002798). Comparisons between two groups were carried out using unpaired Student’s t-test and chi-square test, as appropriate. All the statistical tests were two-sided with p<0.05 was considered as statistically significant. We performed Protein-protein interaction network analysis of Significantly Mutated Genes (SMGs) previously reported in the different studies on CaCx employing the STRING (Search Tool for the Retrieval of Interacting Genes/Proteins) database (**RRID**: SCR_005223) Version 11.0 (28) and visualised the data using Cytoscape (version 3.8.2); **RRID:** SCR_003032. Genes with the highest degree of connectivity in the network i.e., the largest number of interactions with other genes in the network, were identified as hub genes. We performed functional enrichment analysis of the genes harbouring deleterious mutations, including both INDELs and SNVs using DAVID (Database for Annotation, Visualisation and Integrated Discovery) (29,30) and considered the processes and pathways with adjusted p<0.05 (after FDR correction), as statistically enriched. For determining the association between patient survival and PIK3CA mutation status, we employed the Survival package in R. Clinical and genomic data corresponding to HPV16-positive cervical squamous cell carcinoma and endocervical adenocarcinoma from the TCGA-CESC cohort was downloaded from Genomic Data Commons (GDC) (https://portal.gdc.cancer.gov/) and cBioportal.

## Results

### Somatic mutational landscape of Indian patients with HPV16-positive CaCx

To identify the landscape of genomic alterations prevalent among the Indian patients, we generated and analysed WES data of 49 HPV16-positive CaCx patients on their paired samples, blood and tissue. We recorded median coverage of 126X (range: 70-211X), spanning a target region of 39 Mb (exons of 19396 genes), with the median insert size of 160 bp (range: 156-193bp), details provided in **Supplementary Table S2**.

We recorded a total of 6,462 somatic variants identified across 4,598 genes, comprising 1,703 synonymous variants and 4,759 non-synonymous variants. These variants affected 3,585 genes with at least one non-synonymous alteration and 1,530 genes with at least one synonymous alteration. Of the non-synonymous variants, we identified 246 INDELs and the remaining were SNVs. Among the SNVs, 4,020 were missense, 343 nonsense, 142 splice site variants, 5 start codon variants and 3 nonstop variants. Among the INDELs, 128 were frame-shift deletions, 61 inframe deletions, 7 inframe insertions, 1 stop codon deletion and 43 frame shift insertions. In addition, we also identified 4 de novo start inframe and 2 out-of-frame mutations. We have provided the details of the types of mutations recorded in each sample, in **Supplementary Table S5** and pictorially represented in **Figure 1A**.

**Figure 1:**
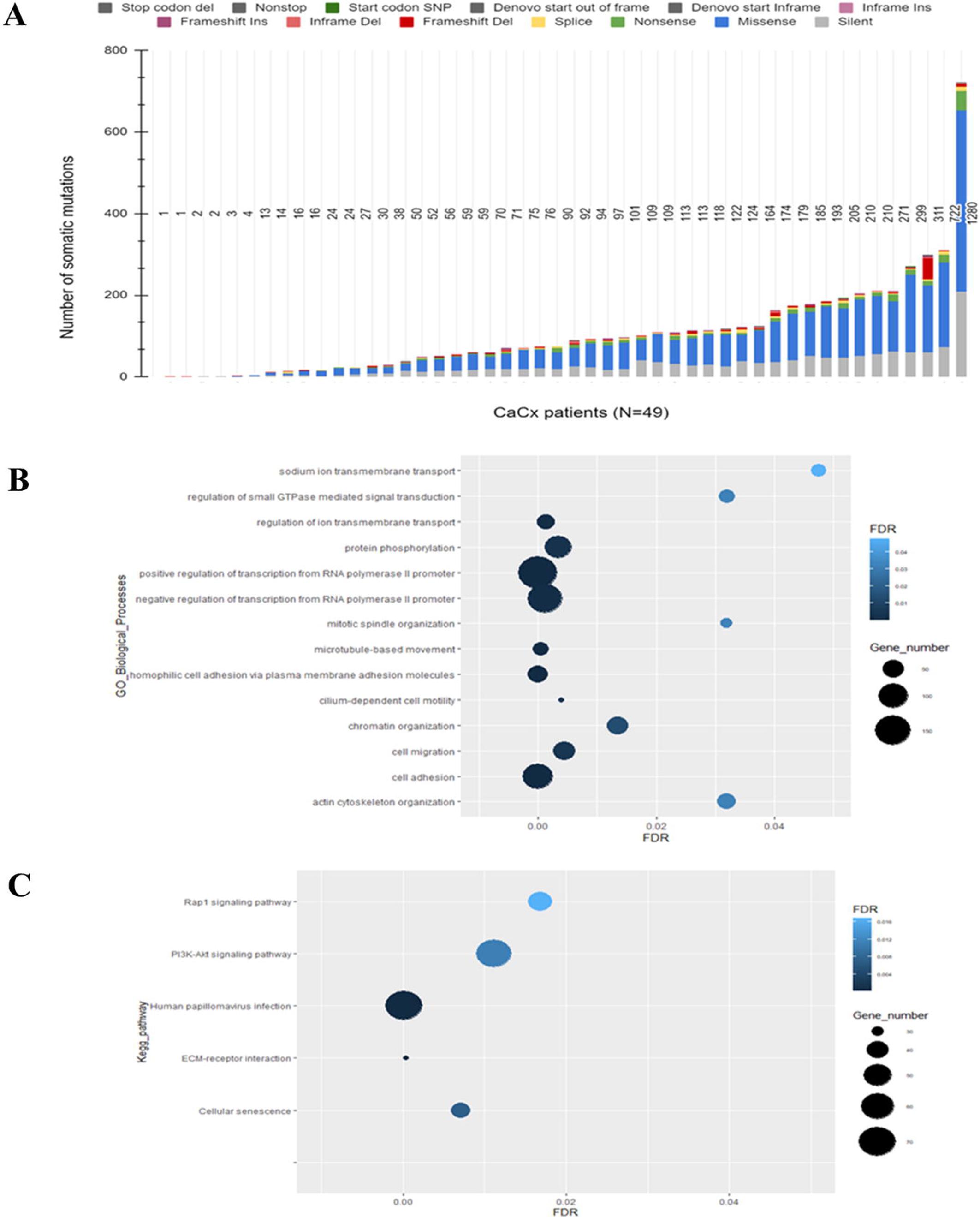
**(A)** Landscape of somatic mutations in HPV16-positive CaCx patients (n=49). The mutation types are colour coded as mentioned in top part of the figure. **(B-C)** Functional enrichment analysis of the mutated genes in HPV16-positive CaCx patients. **(B)** GO-BPs and **(C)** KEGG pathways corresponding to the mutated genes having functional impact on the protein functions. The Y axis represents the names the GO-BPs and KEGG pathways and X axis represent the FDR cut-off <0.05. The colour of the bubble represents the FDR value as per the colour intensity scale provided, while the size of the bubble is correlated with the number of genes belonging to the pathway.

Across the 49 tumor samples, the mean number of somatic variants (SNVs and INDELs) per sample was 132 (range:1-1280) including synonymous variants, and 97.2 (range: 1-931) excluding synonymous variants. The average mutation rate per Mb or Tumor Mutation Burden (TMB), including and excluding synonymous variants was 4.66 and 3.22, respectively, which is nearly concordant with the TCGA-CESC dataset (average total mutation rate/mb = 4.04). We have summarized the descriptive statistics related to the mutation types, in **Supplementary Table S6**.

### Cellular pathways and GO processes associated with the genes functionally impacted by deleterious mutations in CaCx

We identified 2185 genes harbouring deleterious mutations (n=2822 mutations, including both SNVs and INDELs) in HPV16-positive CaCx patients. To decipher the functional role of these mutations in CaCx pathogenesis, we identified the biological processes and pathways associated with the mutated genes, employing DAVID. Some important KEGG pathways (FDR[p]<0.05) identified by such analysis included PI3K-Akt signalling pathway, focal adhesion, Human Papillomavirus infection, ECM receptor interaction and pathways reported in various cancers like small cell lung cancer, prostate cancer, cellular senescence, etc. The GO-BPs, which were significantly (FDR[p]<0.05) associated with these mutated genes, included homophilic cell adhesion, microtubule-based movement, cilium dependent cell motility, cell migration, chromatin organisation, regulation of RNA Pol II promoter, etc. We have provided the details of the KEGG pathways and GO-BP terms in **Supplementary Tables S7 and S8,** respectively, and represented the topmost significant GO-BPs and KEGG pathways, in **Figures 1B and 1C**, respectively.

### Mutation frequency of established Significantly Mutated Genes (SMGs) and driver genes in Indian HPV16-positive CaCx patients

To characterise the mutational landscape of HPV16-positive CaCx in our cohort, we compiled a list of known driver genes and SMGs associated with CaCx reported in various landmark studies, including those by Ojesina et. al, TCGA and other reports (5–7,31–33). The curated gene set comprised PIK3CA, EP300, FBXW7, HLA-B, PTEN, ARID1A, NFE2L2, KRAS, MAPK1, SHKBP1, ERBB3, CASP8, HLA-A, TGFBR2, TP53, STK11, ERBB2, KMT2C, KMT2D, FAT1, MACF1, HUWE1, SPTA1, MED12, CHD8, BCOR, ATXN3, ASXL1, APC, MTOR and ZFHX3. We profiled the somatic mutations in these driver genes in our cohort (n=49) as depicted in **Figure 2**. When multiple types of mutations were recorded in the same gene within an individual patient, we prioritized the most deleterious mutation according to the following hierarchy: Splice site > Frame shift indels > nonsense > missense hotspot > other missense mutations. Among the curated genes, we identified that PIK3CA, KMT2C, KMT2D, EP300, FAT1, MACF1 and HUWE1 harbored somatic mutations in at least 10% (5 out of 49) of the patients and therefore considered these as recurrently mutated genes in our cohort. PIK3CA was the most frequently mutated gene with mutations detected in 43% (21 out of 49) patients, as represented in **Figure 2**. We have provided the details of the somatic mutations identified in these recurrently mutated genes, arranged according to their observed mutation frequency in our cohort, in **Supplementary Table S9.**

**Figure 2:**
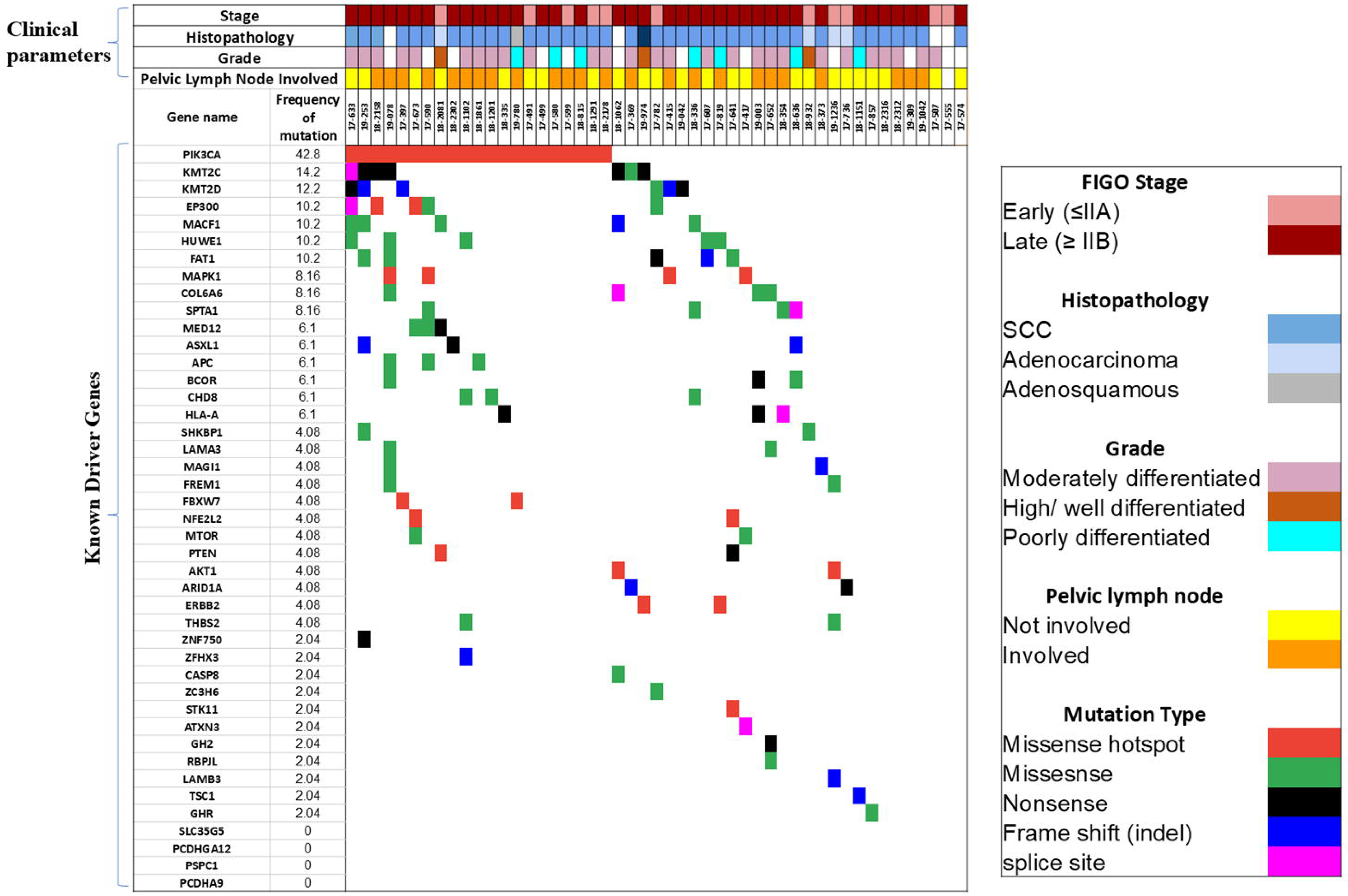
Mutational landscape of known SMGs and recurrently mutated CaCx driver genes in *HPV16-positive* CaCx patients. The SMGs and driver genes are ordered by overall mutation frequency detected in the sample cohort. The rows represent the SMGs and driver genes whereas the columns represent the CaCx patients. The types of alterations across the samples are colour coded as indicated on right side of the figure. Hotspot missense mutations were identified by Mutation Mapper. The clinical parameters of the samples in the cohort are also colour coded as indicated in the figure.

To evaluate population-specific differences, we compared the mutation frequencies of these driver genes between our patient cohort and the TCGA-CESC cohort. Of the 26 genes reported as significantly mutated in the TCGA PanCancer Atlas study, 13 showed higher observed mutation frequencies in our Indian cohort (N=49) compared with the TCGA cohort (N=291). We provided a comparison of the mutation frequency between the two cohorts in **Supplementary Table S10**.

### Identification of potential actionable alterations in HPV16-positive CaCx patients

We assessed the clinical actionability of the curated gene panel employing OncoKB (13). The somatic alterations identified in these genes were considered as potential actionable alterations based on established evidence supporting their role as predictive biomarkers for drug sensitivity to FDA-approved or investigational agents. Overall, 29 patients (59.18%) harboured at least 1 potential actionable alteration across 6 genes. Notably, almost all the alterations in the PIK3CA gene were annotated as actionable alterations. Among 21 patients with PIK3CA-mutations in our cohort, 20 harboured at least one therapeutically actionable alteration, categorised under OncoKB level 4 evidence (biological evidence predicting drug response) (**Table 1**). Some of these alterations were also annotated as level 1 therapeutic evidence (the alterations can predict the response to FDA-approved drugs) in breast cancer. Detailed description of these evidence levels are provided in **Supplementary Methods-Section S4**. In addition to PIK3CA, actionable alterations were identified in PTEN, ARID1A, AKT1, TSC1 and STK11 across 9 patients. These genes also appear to be part of multiple significantly enriched pathways in CaCx in our cohort. Hence targeting these alterations could simultaneously modulate several oncogenic pathways, thereby underscoring the clinical relevance of the corresponding genes in CaCx pathogenesis. Consequently, the patients with such alterations could be categorised for off-label targeted therapy where FDA-approved drugs can be repurposed for CaCx patients and considered for ongoing clinical research on CaCx.

**Table 1:**
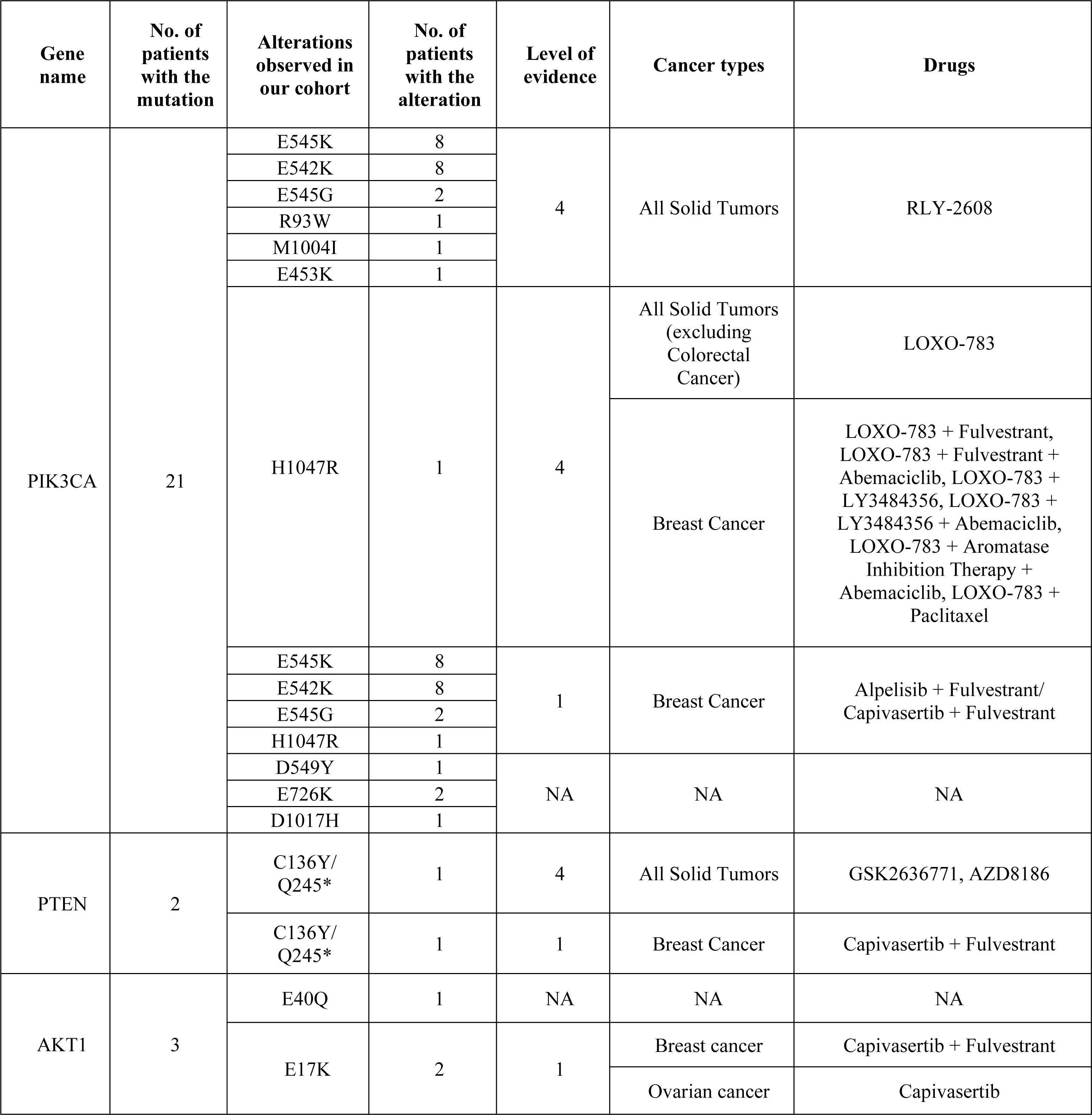
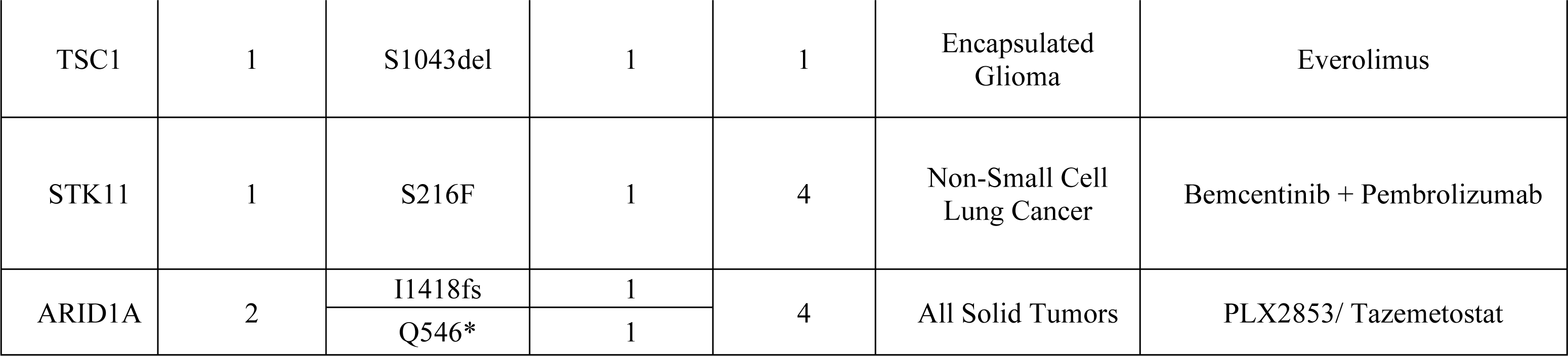
Details of the potentially actionable oncogenic mutations in our cohort and the potentially targeting drugs identified through OncoKB.

### Mutations in PIK3CA gene: most recurrent in Indian cohort of CaCx patients

We identified that ∼43% of the patients harboured mutations in the PIK3CA gene, which were classified as ‘missense’ and were located in the cancer hotspot regions, as represented in **Figure 3A**. About 73.0% (19/26) of these mutations were in the region encoding the helical domain (HD) of PIK3CA protein i.e., Exon 9: E542K (8/26), E545K/G (10/26) and D549Y (1/26). The remaining mutations were observed in the kinase domain (KD), C2 domain and Adapter Binding Domain (ABD) of the PIK3CA protein. Complete details of PIK3CA mutations identified in our cohort are provided in **Supplementary Table S11.** We validated the somatic origin of these PIK3CA mutations using IGV. Representative IGV snapshots (**Figure 3B**) demonstrated the presence of PIK3CA mutations exclusively in mutant tumor tissues, whereas the corresponding blood samples lacked these variants, thereby confirming their somatic origin. These mutations were not detected in PIK3CA-wild tumors. In addition, we also examined the corresponding genomic regions in tissues from healthy individuals and our data revealed that none of the healthy individuals harbored any of these PIK3CA mutations (**Figure 3B)**, confirming their tumor-specific origin.

**Figure 3:**
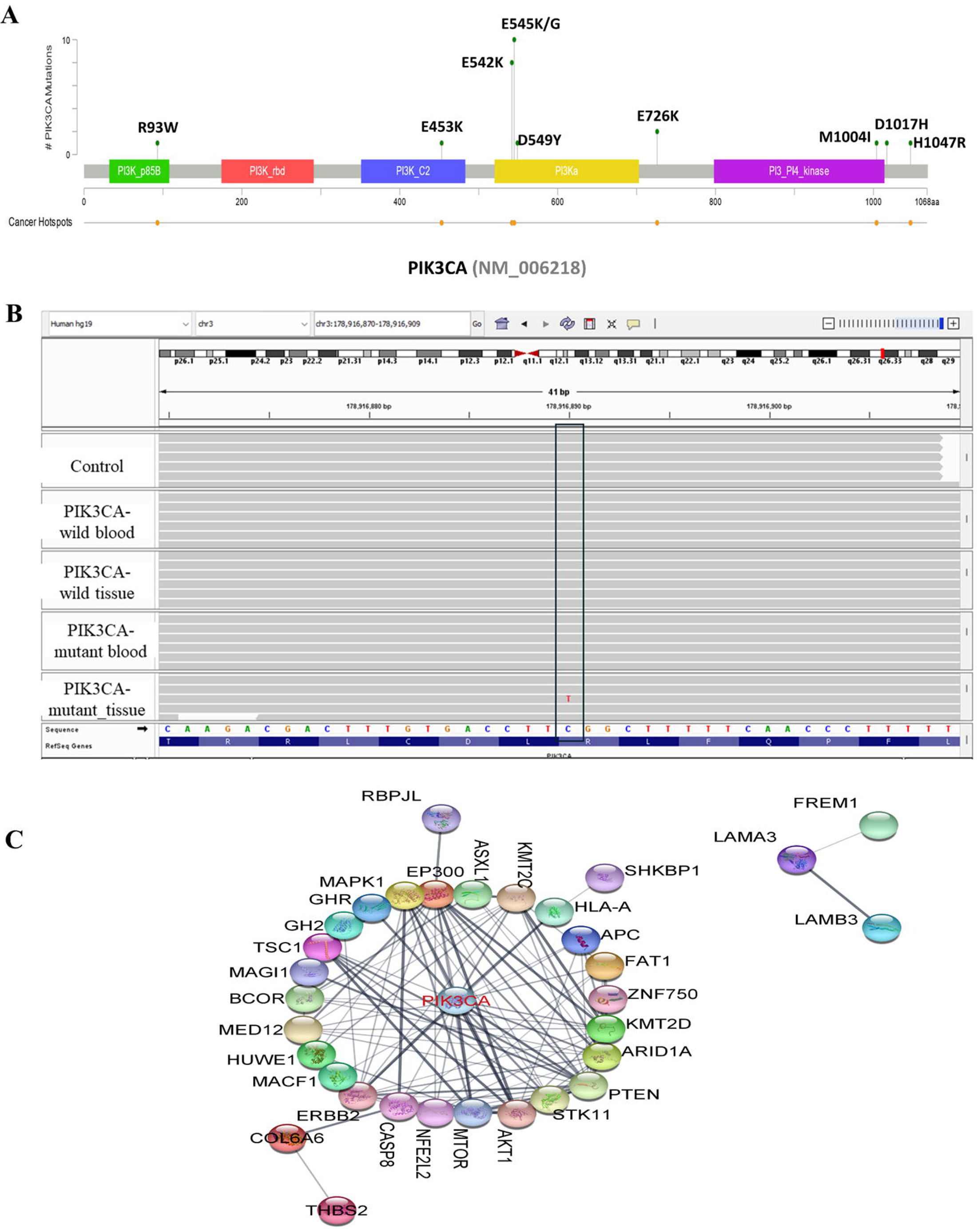
**(A)** Lollipop plot representing the overview of mutations in PIK3CA in *HPV16-positive* CaCx patients of Indian cohort. **(B)** Visualisation of the presence of somatic mutations in PIK3CA gene in healthy individuals (control), PIK3CA-wild and PIK3CA-mutant tumors and blood through IGV. **(C)** Protein-protein Interaction analysis of SMGs by STRING database and visualised through Cytoscape.

Next, we examined the interaction and co-occurrence dynamics among the curated genes within our cohort. Approximately 17% (7/39) of these genes showed moderate levels of co-occurrence with PIK3CA mutations (**Figure 2**). Protein-protein interaction (PPI) network analysis between these genes using Cytoscape revealed that the majority of these genes interacted through PIK3CA, which exhibited highest degree of connectivity between the genes and emerged as the central hub gene. We have provided the details of the degrees of connectivity between the genes in **Supplementary Table S12** and a pictorial representation in **Figure 3C**. Based on these observations, we subsequently characterised the PIK3CA-mutant and PIK3CA-wild tumors with respect to their clinical characteristics, transcriptional perturbations and tumor immune microenvironment.

### Association of PIK3CA mutation with the clinicopathological characteristics, viral factors and tumor mutation burden (TMB) in CaCx patients

Based on the PIK3CA mutation status, we categorised the HPV16-positive CaCx patients (n=49) into 2 groups, PIK3CA-mutant (n=21) and PIK3CA-wild (n=28) tumors and evaluated their association with (i) clinical characteristics, (ii) viral factors and (iii) TMB. There was no evidence of the association of PIK3CA mutations with the age at diagnosis (p _chi-square test_=0.42), FIGO stage (p _chi-square test_ =0.8433) or lymph node involvement (p _chi-square test_ =0.145)-summarised in **Supplementary Table S13;** or patient overall survival (log rank p value = 0.9)-represented in **Supplementary Figure S2A**.

Since the focus of our study is on the most prevalent HPV16-driven CaCx, therefore, we further evaluated the association of PIK3CA mutation with the viral factors like HPV16 E7 oncoprotein expression, HPV16 viral load and physical status. We failed to observe any significant difference in the level of HPV16 E7 expression between PIK3CA-mutant and PIK3CA-wild tumors, based on RNA-seq analysis (TPM-values: mean ± SDs, 176.74 ± 107 vs 209.81 ± 133.85, respectively, p _t-test_ =0.46). Consistent with the RNA-seq findings, qRT-PCR analysis also showed no significant difference in the level of E7 expression between the 2 CaCx subsets (Relative E7 expression: mean ± SD, 15.35 ±5.22 vs 12.83±5.09, p _t-test_=0.11), results depicted in **Supplementary Figure S2 B and C**. When we categorised the CaCx patients into high and low HPV16 viral load (based on the median value of log transformed viral load), we recorded that a greater proportion (14 out of 20; 70%) of PIK3CA-mutant tumors portrayed significantly (p _chi-square test_ =0.02) lower viral load, compared to a smaller proportion (8 out of 23; 34.78%) of the PIK3CA-wild tumors. Conversely, more PIK3CA-wild tumors (15 out 23; 65.21%) portrayed high viral load compared to PIK3CA-mutant tumors (6 out of 20; 30%), as per our depiction in **Supplementary Figure S2D**. The viral genome predominantly existed in the episomal form in both groups, with no significant difference in viral physical status. Specifically, the episomal form was observed in 18 PIK3CA-mutant and 19 PIK3CA-wild tumors, whereas the integrated form was detected in 2 and 3 tumors, respectively.

Further, we observed that PIK3CA-mutant tumors (mean total TMB= 7.8±10.11) showed significantly higher (p_t-test_ =0.006) TMB compared to PIK3CA-wild tumors (mean total TMB =2.31±2.00), as depicted in **Supplementary Figure S2E**. Overall, we also recorded significantly higher number of somatic mutations in PIK3CA-mutant tumors as compared to *PIK3CA-wild* tumors (PIK3CA-mutant vs PIK3CA-wild tumors =4641 vs 1827, p_chi-square test_ <0.0001). Among the different types of mutations recorded in our study, PIK3CA-mutant tumors showed a significantly higher proportion of missense mutations (73.5% vs. 26.4%, p _chi-square test_ < 0.001), nonsense mutations (70.8% vs. 29.1%, p _chi-square test_ < 0.001), frameshift deletions (68.75% vs. 31.25%, p _chi-square test_ < 0.001), and splice-site mutations (63.38% vs. 36.61%, p _chi-square test_ = 0.009) compared with PIK3CA-wild-type tumors.

### Comparative analysis of PIK3CA-mutant and PIK3CA-wild tumors between HPV16-positive Indian CaCx and TCGA-CESC patient cohorts

We compared the frequency of PIK3CA mutation and other associated clinical and viral characteristics between HPV16-positive Indian CaCx and TCGA-CESC cohorts. Out of 165 HPV16-positive TCGA-CESC patients, WES was performed on 103 patients (7); details are provided in **Supplementary Table S14.** PIK3CA was mutated in 27 out of 103 patients (26.21%) in the TCGA-CESC cohort, which was significantly (Fisher’s Exact Test; p = 0.0427) lower than that observed in the Indian patient cohort (42.85%).

To further compare the PIK3CA mutation frequency in diverse clinical settings in the Indian and TCGA cohort of patients, we classified the HPV16-positive PIK3CA-mutant tumors of both the cohorts based on FIGO staging.

A distinct stage-wise distribution was observed between the 2 cohorts. In the Indian cohort, the majority of the PIK3CA-mutant tumors (n=21) were diagnosed at advanced stages with 16 of 21 cases (76.19%) classified as late stage (FIGO ≥IIB) and only 5 (23.8%) as early stage (FIGO <IIB, **Supplementary Table S13**). In contrast, the TCGA cohort (n=27) predominantly comprised early-stage PIK3CA-mutant tumors, with 16 of 27 cases (59.25%) being early stage and 11 (40.74%) late stage (**Supplementary Table S15**).

Comparative analysis considering the early-stage PIK3CA-mutant tumors revealed a significantly higher proportion in the TCGA-CESC cohort than in the Indian cohort (59.3% vs. 23.8%; p _chi-square test_ = 0.014). Conversely, late-stage PIK3CA-mutant tumors were significantly more frequent in the Indian cohort than in the TCGA-CESC cohort (76.2% vs. 40.74%; p _chi-square test_ = 0.014). These findings suggest that unlike the TCGA-CESC cohort where PIK3CA mutations are more commonly detected in early-stage disease, the Indian cohort exhibits a significant association with advanced tumor stages. This clearly reflects a population-specific phenomenon that underscores PIK3CA mutations as relevant therapeutic targets for Indian patients who mostly present with advanced-staged disease.

In both the cohorts, all PIK3CA mutations were of missense type and located in the cancer hotspot regions, encoding p.E542K and p.E545K (**Supplementary Figure S3A and Figure 3A)**. In the TCGA-CESC cohort, presence of PIK3CA mutations was significantly associated with absence of node involvement (p_chi-square test_ =0.041; **Supplementary Table S15)**. However, there was no significant difference between PIK3CA-mutant (n=27) and PIK3CA-wild (n=76) tumors with respect to age (p_t-test_=0.06), tumor-stage (p_chi-square test_=0.946) or patient OS (log rank p value= 0.6); as detailed in **Supplementary Table S15** and **Supplementary Figure S3B**. Further, no significant difference (p_t-test_= 0.7) in HPV16 E7 expression (TPM) was observed between PIK3CA-mutant (n=24, mean ± SD = 12413.58±13740.48) and PIK3CA-wild (n=62, mean ± SD =11330.37±11086.48) tumors. Similar findings were observed in our Indian cohort of patients, indicating that although PIK3CA harbours oncogenic hotspot mutations, the presence of mutations is unlikely to have any prognostic significance in HPV16-positive CaCx.

### Identification of DEGs in HPV16-positive CaCx patients with and without PIK3CA mutations

To understand the pathological role of gain-of-function PIK3CA mutations and to identify genes deregulated in association with PIK3CA mutation, we analysed the RNA-seq data [ENA accession number: PRJEB40877] generated by us, on a subset of HPV16-positive tumor samples (n=29), comprising 12 PIK3CA-wild tumors and 17 PIK3CA-mutant tumors. Considering the FDR-adjusted p<0.05 and |log2(Fold Change) | > 0.6 (corresponding to ≥ 1.5-fold differential expression) as thresholds, we identified 27 differentially expressed genes between PIK3CA-mutant and PIK3CA-wild tumors. Some of these differentially expressed genes (DEGs) included MARCO, STAG3, HOXD13, PRAC2, PTHLH, ADAMTS19, FABP4, PWRN1, etc; listed in **Supplementary Table S16**. Functional annotation revealed that these DEGs were associated with key oncogenic processes like proliferation, cell migration, self-renewal, anti-apoptosis, epidermis development, ECM organisation, etc. Many of these genes have been reported to be involved in various cancers, and their deregulation is associated with cancer progression and metastasis. We, therefore, hypothesize that PIK3CA mutations could be associated with deregulated expression of oncogenic genes contributing to more aggressive cancer phenotypes, compared to PIK3CA-wild tumors.

### Tumor Immune microenvironment (TIME) of PIK3CA-mutant and -wild tumors compared to healthy individuals

Tumor microenvironment (TME) plays an important role in cancer development, progression, and treatment outcome, and is particularly relevant for immunotherapy. The implementation of immunotherapy in CaCx, targeting the PD-1/PD-L1 pathway, prompted us to explore the tumor immune microenvironment (TIME) in such cancers. We therefore compared the TIME of PIK3CA-mutant and -wild tumors, compared to healthy individuals, in respect of 3 biomarkers for immunotherapy-(i) Tumor Immune Infiltrating Cells (TIICs), (ii) immune active component through Tertiary Lymphoid Structures (TLS) enrichment and maturation, and (iii) the immune suppressive axis, mediated by immune checkpoint PD-1/PD-L1 axis.

### A. Immune cell infiltration in the tumor milieu of PIK3CA-mutant and -wild tumors compared to healthy individuals

Immune cell composition and the crosstalk between different populations of immune cells in the TME play an important role in modulating the therapeutic response to immunotherapy (34). Therefore, we further evaluated the proportion of immune cells in PIK3CA-mutant (n=17) and -wild tumors (n=12), compared to cervical tissues from healthy individuals (n=34), employing CIBERSORT-abs from bulk RNA-seq data. Out of 22 immune cell types, we found significantly (p _t-test_ <0.05) higher proportions of CD4+ memory activated T cells, Tregs, M0 and M1 macrophages and resting Dendritic cells (DC) in both PIK3CA-wild and PIK3CA-mutant tumors, compared to non-tumor tissues of healthy individuals. In addition, a significantly higher proportion of other immune cells, such as memory B cells, Plasma B cells, CD8+ T cells, activated NK cells, M2 macrophages, and resting mast cells, was observed in the PIK3CA-wild type tumors, compared to healthy individuals. PIK3CA-mutant tumors, on the other hand, did not show any significant difference in the proportions of CD8+ T cells, compared to healthy individuals. We have represented the proportion of all 22 cell types in PIK3CA-mutant tumors, PIK3CA-wild tumors and normal tissues from healthy individuals in **Figure 4A**. The findings thereby suggest the existence of an immune-enriched TME in PIK3CA-wild tumors over that of PIK3CA-mutant tumors, when compared to healthy individuals.

**Figure 4:**
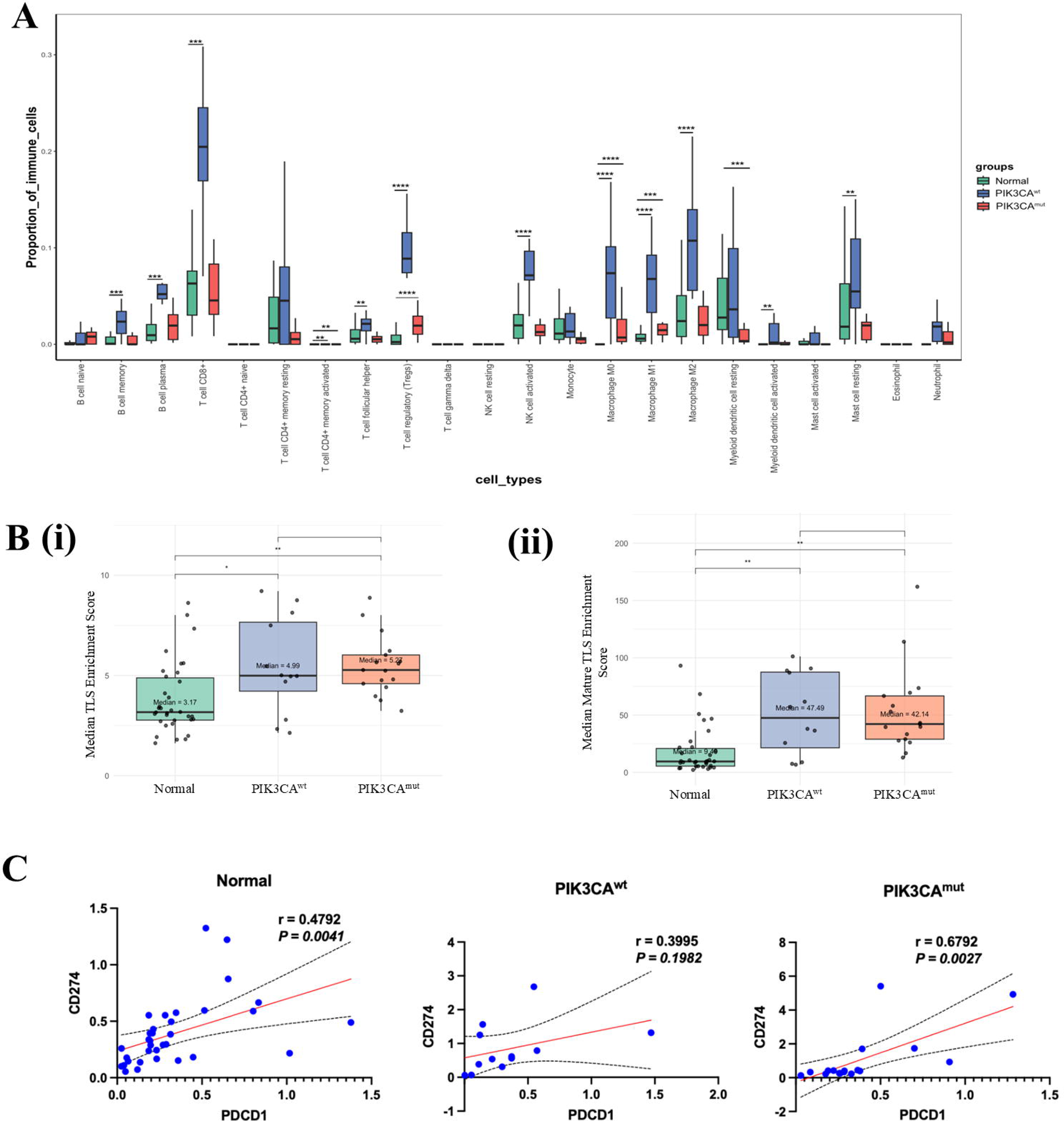
**(A)** Box plot representing the distribution of proportion of 22 TIICs in normal cervical tissues from healthy individuals (Normal; n=34), PIK3CA-wild tumors (PIK3CA^wt^; n=12) and PIK3CA-mutant tumors (PIK3CA^mut^; n=17). The Y axis represents the proportion of different immune cell types and X axis represents the 22 immune cell types. “*” represents significant p < 0.05. “**” represents significant p < 0.01, “***” represents p<0.001 **(B)** Box plot representing **(i)** overall median TLS enrichment score distribution and **(ii)** median Mature-TLS enrichment score in Normal, PIK3CA^wt^ tumors and PIK3CA^mut^ tumors. The TLS score was derived by calculating the geometric means of the TLS signature genes. “*” represents significant p value < 0.05. “**” represents significant p value < 0.01**. (C)** Sample-wise correlation plots depicting the correlation between PDCD1 and CD274 expression (TPM) in Normal, PIK3CA^wt^ tumors and PIK3CA^mut^ tumors. The red line indicates the linear regression fit, while the black line represents the 95% confidence interval. Correlation coefficients (*r*) and corresponding *p* values were calculated using Pearson’s correlation analysis.

### B. TLS enrichment and maturation in PIK3CA-mutant and -wild tumors compared to healthy individuals

In order to determine whether the infiltrating immune cells are organized and functionally active, we analysed the spatial aggregation of TLS within the TME, which reflects an ongoing anti-tumor immune response, in CaCx patients (n = 29) and normal cervical tissues from healthy individuals (n=34) by calculating the TLS enrichment scores. Among the tumors, which included both PIK3CA-wild (n = 12) and PIK3CA-mutant (n = 17) tumors, 51.7% were classified as TLS-High, 37.9% as TLS-Intermediate and 10.3% as TLS-Low. Comparing the 2 subtypes, we observed an enrichment of TLS-High tumors in PIK3CA-mutant group (58.8%) than in -wild group (41.7%), although this difference was not statistically significant. In contrast, we observed a markedly different distribution in the healthy individuals (n = 34) with 44.1% exhibiting TLS-Low phenotype, 32.4% as TLS-Intermediate, and only 23.5% as TLS-High.

When we compared the total TLS enrichment score between the subtypes, we observed significantly (p _t test_ <0.01) higher TLS enrichment in both PIK3CA-mutant (median = 5.27) and PIK3CA-wild tumors (median = 4.99) compared to healthy individuals (median= 3.17), while no significant difference was observed between the 2 tumor subtypes; data represented in **Figure 4B(i)**.

The density and maturation status of TLS critically influences tumor progression and response to immunotherapy (35). Hence, we assessed the stage-specific enrichment scores for early, intermediate and mature TLS after Z-score normalisation to allow relative comparison across samples of different groups i.e, PIK3CA-mutant, PIK3CA-wild and healthy individuals. We did not observe any significant differences between the groups with respect to early TLS (median scores: PIK3CA-mutant = 3.01, PIK3CA-wild = 2.73, Normal = 2.71; *p*_t test_ > 0.05) or intermediate TLS (PIK3CA-mutant = 2.02, PIK3CA-wild = 1.94, Normal = 1.80; *p*_t test_ > 0.05). In contrast, we found marked enrichment of the mature TLS among tumor tissues, including both PIK3CA-mutant (median= 42.14) and -wild tumors (median= 47.49) compared to healthy individuals (median=9.49; *p*_t test_ < 0.01; **Figure 4B(ii)**). Although PIK3CA-wild tumors had a higher median mature TLS score than PIK3CA-mutant tumors, the difference was not statistically significant (p _t test_ > 0.05).

Our findings highlight that HPV16-positive CaCx tumors are characterised by enhanced TLS enrichment, particularly mature TLS compared to the normal cervical tissues, regardless of PIK3CA mutation status, supporting the notion that TLS generation is tumor-associated, perhaps driven by HPV16, rather than mutation-specific. We observed comparable levels of TLS enrichment and maturation in the two tumor subtypes, irrespective of the fact that the PIK3CA-wild tumours portrayed higher TIICs infiltration than the mutant subtype. Thus, the PIK3CA-mutant tumors may harbor fewer immune cells but still support comparable TLS-driven immune activation as the PIK3CA-wild tumors.

### C. PD-L1 and PD-1 expression in PIK3CA-mutant and -wild tumors compared to healthy individuals

Emerging evidence (36,37) has demonstrated a strong association between PIK3CA mutations, HPV16 infection, and PD-L1 overexpression, at both the mRNA and protein levels in CaCx. These observations suggest that a convergent host oncogenic and viral mechanisms may promote immune evasion mechanism through the PD-1/PD-L1 axis. Although PIK3CA-mutant and PIK3CA-wild tumor subtypes showed differences in immune cell infiltration, they exhibited comparable TLS-mediated immune activation. We, therefore, investigated whether these immune responses are functionally constrained by immune checkpoint signalling, specifically the PD-1/PD-L1 axis.

Compared to healthy individuals, CD274 (encoding PD-L1) expression was significantly upregulated in both PIK3CA-mutant tumors (|log_2_(FC)| = 1.14, FDR corrected *p*=0.005) and PIK3CA-wild tumors (|log_2_(FC)| = 1.09, FDR corrected *p*=0.022). However, PDCD1 (encoding PD-1) expression was selectively upregulated in PIK3CA-mutant tumors (|log_2_(FC)| = 0.88, FDR corrected *p*=0.03) compared to healthy individuals, whereas no significant differential expression was observed in PIK3CA-wild tumors (|log_2_(FC)| = 0.24, FDR corrected *p*=0.68). The transcript levels of both PDCD1 and CD274 did not differ significantly between the two tumor subtypes.

Further to examine the coordinated expression of PD-1/PD-L1 axis, we determined the correlation between the expression levels of PD-1 and PD-L1 transcripts across the 3 groups using TPM data of RNA-seq. We observed a significant positive correlation between PDCD1 and CD274 expression in healthy individuals (Pearson’s correlation coefficient, r=0.47, p=0.004) and PIK3CA-mutant tumors (r=0.67, p=0.002), whereas this correlation did not reach statistical significance in PIK3CA-wild tumors (r=0.39, p=0.198); represented in **Figure 4C**. Such significant correlation may reflect coordinated engagement of the PD-1/PD-L1 axis, which may have translational relevance in PIK3CA-mutant tumors. Notably, as PD-1/PD-L1 signalling is influenced by the abundance of CD8+ T cells in the TME, we interpreted these findings in the context of immune cell infiltration analysis. While PIK3CA-wild tumors exhibited enhanced CD8+T cells infiltration, the PIK3CA-mutant tumors portrayed CD8+T cell levels comparable to normal tissues. Taken together, our observation points towards distinct immune regulatory landscapes prevailing in the two tumor subtypes.

### Subtype-specific chemokine signaling implicates CCL3-CCR1 axis as a potential driver of immune modulation in PIK3CA-mutant tumors

Having characterized the immune landscape in terms of TIICs composition, TLS-mediated immune organisation, and PD-1/PD-L1 axis driven immune suppression, we sought to draw subtype-specific molecular cues in PIK3CA-mutant and -wild tumors that could potentially underlie these differences. Since chemokines are the key regulators of immune cell recruitment within the TME, we investigated the chemokine-mediated CCL3–CCR1 (C-C motif chemokine ligand 3, CC chemokine receptor 1) signalling axis which is known to modulate both anti-tumor immunity and immunosuppressive cell infiltration (38–40). We recorded a significant upregulation of the expression (TPM) of both CCL3 and CCR1, specifically within the PIK3CA-mutant tumors in comparison with the healthy individuals (**Figure 5A (i) and (ii)**). The PIK3CA-wild tumors did not portray any such alterations in the CCL3 and CCR1 expression levels.

**Figure 5:**
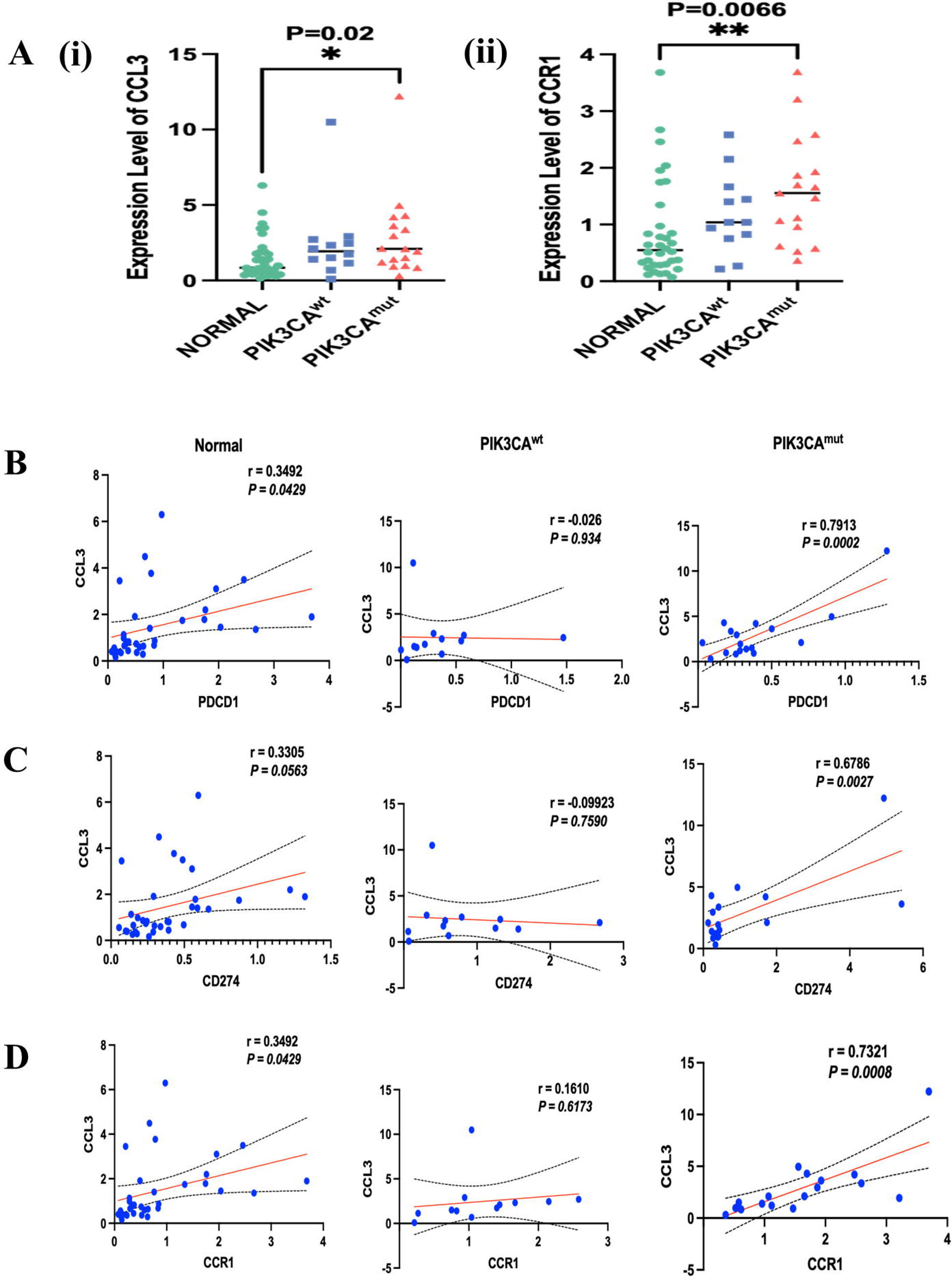
**(A)** Scatter dot plots portraying the relative expression levels of **(i)** CCL3 and **(ii)** CCR1 in Normal, PIK3CA^wt^ tumors and PIK3CA^mut^ tumors. Statistically significant differences between groups are indicated by the corresponding p values above the plots; p < 0.05 was considered significant. Sample-wise correlation plots depicting the correlation between expression levels (TPM) of CCL3 and **(B)** PDCD1, **(C)** CD274 and **(D)** CCR1 in Normal, PIK3CA^wt^ tumors and PIK3CA^mut^ tumors. The red line indicates the linear regression fit, while the black line represents the 95% confidence interval. *r* and *p* values were calculated using Pearson’s correlation analysis.

Further to determine whether the CCL3-CCR1 axis is linked to PD-1/PD-L1 axis, we assessed the correlation between expression (TPM) of CCL3 and PDCD1/CD274. We observed a strong correlation between expression of CCL3 and CD274 (r=0.68, p=0.003), CCL3 and PDCD1 (r=0.79, p=0.0002), and CCL3 and CCR1 (r=0.73, p=0.0008), within the PIK3CA-mutant tumors (**Figure 5B-D**), in contrast to a weak correlation in PIK3CA-wild tumors. Such observations suggest a coordinated regulation of CCL3/CCR1/PD-1/PD-L1 axis specifically in the PIK3CA-mutant subtype.

To further elucidate the functional relevance of CCL3 in shaping the immune landscape of PIK3CA-mutant tumors, we examined its association with immune-activating cytokines and co-stimulatory molecules together with key cytotoxic, myeloid, Treg, and interferon-γ–related immune gene signatures. Compared to the healthy individuals, we observed a significant downregulation of the expression of IL-2, CD40LG, and IL12B in the PIK3CA-mutant tumors (**Supplementary Figure S4 A-D**). We further observed a strong positive correlation of CCL3 expression with cytotoxic T-cell markers (CD8A, GZMB, PRF1 and IFNG), interferon-γ–related immune signatures (STAT1, IRF1, CXCL9 and CXCL10), and myeloid lineage markers (CD68 and CD163), but not with Treg-associated markers (FOXP3 and IL2RA). The data is presented in **Supplementary Figures S5-S8**. Collectively, these findings establish CCL3 as a central mediator of a subtype-specific immune remodelling program in PIK3CA-mutant tumors, characterised by coordinated inflammatory cytotoxic signaling, immune checkpoint and myeloid-associated immunoregulatory remodelling, rather than preferential Treg enrichment.

## Discussion

Although HR-HPV infection is indispensable for CaCx development, the role of host genetic factors in the disease pathogenesis cannot be disregarded. Despite several large-scale genomic studies that have identified the mutational landscape of CaCx in various populations (6,7,31,33,41), the genomic characterization of Indian CaCx patients remains largely underrepresented in publicly available databases, including the TCGA database, which incorporated only 20 Asian patients and no Indian patients. Therefore, our study provides the first comprehensive characterisation of HPV16-positive CaCx patients of Indian origin, integrating genomic, transcriptomic, and immunogenomic analyses revealing both shared and population-specific molecular features with important implications for therapeutic stratification.

Our study, employing WES analysis, revealed that most of the non-synonymous mutations were functionally relevant and the affected genes were involved in cancer-related biological processes indicating that the key biological pathways involved in driving other cancers are also frequently mutated in HPV16-positive CaCx. Among these, PI3K-Akt signalling emerged as the most significantly altered pathway. Consistent with the previous reports on CaCx (7,42) PIK3CA gene was also the most frequently altered gene in our patient cohort. Notably, the prevalence of PIK3CA mutations recorded by us (42%) was also significantly higher than that reported in the HPV16-positive TCGA-CESC cohort (26%) as well as another study (8) with South Indian CaCx patient cohort (34%). Such differential observations appear to be suggestive of potential population-specific differences in the molecular evolution of HPV16-positive CaCx. Therefore, our findings underscore the importance of studying geographically and ethnically distinct patient populations.

As comprehensive molecular characterization of PIK3CA-mutant versus -wild tumors in HPV-driven CaCx remains limited, in this study, we evaluated the molecular profiles of PIK3CA-mutant and -wild tumors. Consistent with some previous reports on solid tumors (43,44) and our observations from TCGA-CESC dataset analysis, PIK3CA mutation did not reveal any significant association with clinicopathological parameters, patient survival or E7 oncoprotein expression. Importantly, unlike TCGA-CESC cohort, where PIK3CA alterations were predominantly associated with early-stage tumors, our cohort demonstrated that PIK3CA mutations were associated with advanced-stage disease, suggesting that PIK3CA mutations may contribute not only to tumor initiation but also to disease progression and acquisition of aggressive phenotypes in Indian patients.

Furthermore, PIK3CA-mutant tumors exhibited significantly lower HPV16 viral loads and higher TMB compared with their wild-type counterparts. This observation may reflect a shift from viral oncogene dependence toward host genome-driven tumor evolution during disease progression. Such a transition has been proposed in several HPV-associated malignancies (7,45), wherein the accumulation of somatic alterations progressively assumes a dominant role in sustaining tumor growth and survival. The elevated TMB observed in PIK3CA-mutant tumors is indicative of increased genomic instability and may further contribute to tumor evolution and aggressive clinical behaviour (46). Consistent with this notion, transcriptomic analysis further revealed enrichment of genes in PIK3CA-mutant tumors associated with cell proliferation, migration, resistance to apoptosis, epidermal development, and extracellular matrix organization. Together, these findings suggest that gain-of-function mutations in PIK3CA, in conjunction with an increased mutational burden, may potentiate HPV-driven oncogenesis by promoting a more aggressive tumor phenotype.

The reduced viral load observed in mutant tumors are also likely to result from impaired viral replication under the impact of loss of tumor cell differentiation. Such changes clearly reflect an abortive HPV life cycle, leading to reduced virion production and consequently lower viral burden. This phenomenon is consistent with the well-established dependence of the HPV replicative cycle on epithelial differentiation, whereby cellular pathways regulating epithelial maturation also influence the transcription of viral late genes essential for productive infection (47).

Tumor genomic profiling is being widely used in recent times for the selection of FDA-approved and investigational therapies for different cancers (48). For late-stage CaCx patients, standard therapeutic regimens lead to therapy resistance and disease recurrence, highlighting the need for alternative therapeutic regimens. The high frequency of actionable alterations in PIK3CA and other curated driver genes in our cohort has important therapeutic implications. About 59% (29 out of 49 patients) of the patients harboured potentially actionable alterations, supported by OncoKB evidence, with some already linked to FDA-approved agents in breast cancer. This opens avenues for repurposing PI3K pathway inhibitors such as PIK3CA inhibitors (Alpelisib), Akt inhibitors, mTOR inhibitors (Temsirolimus, Everolimus, Rapamycin) etc., in advanced CaCx, especially in Indian populations.

Tumor genomic profiling is further well established to characterize the TIME. Various studies, including one from our laboratory (15), highlighted the immune heterogeneity among the HPV16-positive CaCx patients from eastern India. Such findings portray the fact that not all patients are likely to benefit from a single therapeutic regimen. Therefore, in this study, we moved forward to gain a comprehensive understanding of the TIME in the context of novel therapeutic approaches, specifically immunotherapy. We characterised the TIME of PIK3CA-mutant and -wild tumors and compared with the normal tissues from healthy individuals with respect to multiple immune markers for better prediction of therapeutic response. Our findings unfurled intriguing features of PIK3CA-mutant tumors. Despite their aggressive molecular characteristics, these tumors failed to exhibit evidence of immune exclusion. Mature TLS were preserved and Immune-cell infiltration, including CD8+ T cells, remained at comparable levels to healthy tissues. These findings suggest that immune recognition of the tumor persists and that tumor progression is unlikely to result from a complete failure of immune recruitment.

Our observation of coexistence of immune cell infiltration with elevated PD-L1 and PD-1 transcript expression and a statistically significant PD-1/PD-L1 transcriptional coupling in PIK3CA-mutant tumors, points toward the presence of a functionally suppressed or dysregulated immune microenvironment through immune checkpoint activation; while PIK3CA-wild tumors exhibited immune infiltration without a significant PD-1/PD-L1 transcriptional association. Interestingly, such observations are further supported by a recent single-cell transcriptomic data on CaCx TME (49), which demonstrated that PIK3CA-mutant tumors exhibit a dual immune phenotype characterized by pronounced T-cell inflammation together with increased CD8+ T-cell exhaustion. Additionally, the study identified enhanced PD-L1/PD-1 ligand–receptor signaling between malignant epithelial cells and T-cell populations in PIK3CA-mutant tumors, with significantly stronger signaling interactions than those observed in PIK3CA-wild tumors. Further, the authors showed that CD8+ T cells in PIK3CA-mutant tumors preferentially portrayed a terminally exhausted state, while malignant epithelial cell subsets expressing PD-L1 were spatially associated with PD-1+ CD8+ T cells.

Therefore, such independent single-cell observations (49) provide robust support to our finding of significant PD-1/PD-L1 transcriptional coupling in PIK3CA-mutant tumors reflecting a biologically relevant checkpoint-mediated immune regulation within the TME. Taken together, these data indicate that PIK3CA-mutant tumors possess a T-cell inflamed but exhausted TME, in which immune infiltration persists and coexists with coordinated checkpoint activation. This may represent an alternative mechanism of immune evasion associated with PIK3CA mutation and warrants further functional investigation. Such tumors, thereby, present as promising candidates for immune checkpoint therapy targeting PD-1 and consequently the PD-1/PD-L1 axis.

Another notable finding recorded in our study was the selective activation of the CCL3–CCR1 signaling axis in PIK3CA-mutant tumors. The positive correlation of this pathway with immune checkpoint molecules, cytotoxic, interferon-response, myeloid markers suggest the presence of a complex inflammatory microenvironment characterized by concurrent immune activation and immune suppression. Emerging evidence from other malignancies supports a critical role for CCR1-mediated signaling in shaping tumor immune landscapes. In Gram-negative microflora-infected esophageal squamous cell carcinoma (ESCC), activation of the CCL3/CCL5– CCR1–MAPK–PD-L1 axis was shown to promote immune evasion and tumor progression through enhanced PD-L1 expression (50). Similarly, a study on melanoma demonstrated spatial colocalization of CCR1⁺ macrophages with exhausted CD8⁺ T cells, indicative of an immunosuppressive tumor niche (51).

In the context of our findings, the selective activation of the CCL3–CCR1 axis together with enhanced PD-1/PD-L1 transcriptional coupling in PIK3CA-mutant tumors raises the possibility that CCR1-dependent signaling contributes to the establishment of a functionally suppressed immune microenvironment despite preserved immune-cell infiltration. Furthermore, previous studies have implicated the CCL3–CCR1 axis in the recruitment of myeloid-derived suppressor cells, macrophage polarization, and maintenance of chronic inflammatory states that facilitate tumor progression (52). The observed downregulation of IL2, CD40LG, and IL12B provides additional evidence of impaired maintenance of effective anti-tumor immunity in PIK3CA-mutant tumors. These molecules are central regulators of T-cell activation, antigen presentation, and sustained effector immune responses. Their reduced expression suggests that although immune cells are recruited to the tumor microenvironment, the signals required for durable and productive anti-tumor activity may be compromised. This may contribute to the paradoxical coexistence of immune infiltration and aggressive molecular phenotypes observed in PIK3CA-mutant tumors. Although the precise mechanistic relationship between PIK3CA activation and CCR1 signaling in CaCx remains to be elucidated, to the best of our knowledge, our data appears to be the first report to establish the CCL3–CCR1 pathway as a potentially important link between PIK3CA mutation and immune dysregulation. This warrants further functional investigation and exploration as a therapeutic target.

Some recent molecular classifications (53) have identified PIK3CA-mutant CaCx as a distinct genomic subtype essentially based on tumor-intrinsic molecular characteristics. However, our data suggest that mutational status alone may not fully capture the biological heterogeneity associated with PI3K pathway activation. In the HPV16-positive cohort of patients from eastern India, based on integrative genomic and transcriptomic analyses, PIK3CA-mutant tumors exhibited a significant enrichment of these mutations in advanced-stage tumors. Besides, such tumors portrayed a combination of high mutational burden, reduced viral load, and a uniquely immunomodulated TME that has not been emphasized in previous subtype-defining studies. These findings indicate that PIK3CA mutations may serve as both genomic and immune determinants of disease behaviour, thereby emphasizing the importance of population-specific molecular characterization in cervical cancer.

While the strength of our study is based on the integration of whole-exome and transcriptomic profiling to characterize both tumor-intrinsic and immune-related alterations in a clinically relevant HPV16-positive cohort, there are some limitations that need to be addressed. The modest sample size, particularly for transcriptomic analyses, requires validation in larger independent cohorts. Therefore, further functional experiments seem to be necessary to establish causal relationships between PIK3CA mutations, immune dysregulation, and activation of the CCL3–CCR1 pathway. Besides, studies incorporating single-cell and spatial transcriptomics, along with longitudinal clinical data and therapeutic response information will further clarify the prognostic and predictive value of these molecular features.

In conclusion, our study unfurls PIK3CA-mutant HPV16-positive CaCx as a molecularly aggressive and immunologically distinct subtype in Indian patients. The combination of increased mutation frequency, association with advanced-stage disease, altered immune checkpoint signaling, and evidence of functional immune suppression through CCL3-CCR1 axis highlights the potential utility of PIK3CA mutational status as a biomarker for patient stratification and precision therapeutic approaches. These findings also emphasize the importance of population-specific cancer genomics studies towards uncovering biologically relevant differences that may influence disease management and treatment outcomes. Most importantly, our study justifies the potential of PIK3CA mutation as a predictive biomarker for combination therapy involving suitable PI3K inhibitors and immunotherapy targeting the PD-1 and PD-L1 axis in HPV16-positive CaCx.

## Declarations

### Authors’ contributions

AG^1^ and SSG^1,5^ conceived the study; AG^1^ performed the experiments, AG^1^, NKB^1^ and SSG^1,5^ wrote the manuscript; NKB^1^ and AG^^1^ developed the Whole exome and RNA-seq data analysis pipeline and was involved in raw data processing including identification of somatic mutations and differential gene expression analysis. AG^1^, AS^1a^, SM^4^ and SSG^1,5^ performed downstream data analysis, SM^2^, JB^2^, AM^3^ coordinated patient recruitment, clinical assessments and collection of data on patients and provided patient materials; AM^1^ coordinated massively parallel sequencing experiments; SM^4^, KM^5^ and AG^1^ conducted the analysis and interpretation of the data on TLS and chemokine signalling analyses; PPM^6^ provided guidance on the study design and analysis and SSG^1,5^ provided overall guidance of the study; all authors contributed towards reviewing the manuscript.

### Ethics statement

All samples, from both healthy individuals and cancer patients, were collected with written informed consent. The study was approved by the Institutional Ethics Committees of the collaborating institutions-College of Medicine and Jawaharlal Nehru Memorial Hospital, Kalyani (Ref No.: F-24/PR/COMJNM/IEC/20/50/1 dated February 7, 2020) and Tata Medical Center, Kolkata (Ref No.: EC/GOVT/18/17 dated January 3, 2018). As National Institute of Biomedical Genomics (NIBMG), Kalyani did not independently collect the clinical samples, the Institutional Ethical Committee (IEC) of NIBMG was satisfied that members of NIBMG can participate in the research, as IECs of the two primary institutions College of Medicine and Jawaharlal Nehru Memorial Hospital, Kalyani and Tata Medical Center, Kolkata collecting data and clinical samples, had approved the undertaking of this research.

### Data availability statement

Raw Exome sequence data (BAM files) of individual patients included in the study have been deposited in European Nucleotide Archive (ENA) via accession code: PRJEB71014 and secondary accession code: ERP155903. Raw RNA seq BAM files of individual subjects included in the study are available in ENA via accession number: PRJEB40877 and secondary accession number: ERP124576.

## Acknowledgements

We are grateful to the Government of India, Department of Biotechnology, Ministry of Science and Technology, for financial support to this study, through the Systems Medicine Cluster (SyMeC) project (BT/Med-II/NIBMG/SyMeC/2014/Vol. II). We are also grateful to all participating members of SyMeC for their inputs and contributions to this study. We acknowledge Tata Medical Center, Kolkata, and College of Medicine and J.N.M. Hospital, Kalyani, for providing us with the clinical samples for the study; all the members of the Core Technology Research Initiative (CoTeRI) of the National Institute of Biomedical Genomics, Kalyani, India for their technical support during the work; and special thanks to Council of Scientific and Industrial Research (CSIR), India, for providing research fellowship to Abhisikta Ghosh (JRF and SRF), Department of Science and Technology (INSPIRE), Govt. of India for providing research fellowship to Abarna Sinha (JRF and SRF). We also acknowledge ICMR-SRF (GENOMICS-BMS/2021-10619) and RCB (RCB/NIBMG-PhD/2022-23/A/394/1002) for providing fellowship to Arnab Ghosh.

## Supplementary Figure Legends

**Supplementary Figure S1-**Amplification plot of **(A)** viral oncogene HPV16 E7 and **(B)** 18srRNA (house-keeping gene) by TaqMan-based qRT-PCR

**Supplementary Figure S2-Association of clinicopathological features and viral factors with PIK3CA mutation in our cohort of HPV16 positive CaCx patients. (A)** Kaplan Meier plot depicting the overall survival (OS) of the HPV16 CaCx patients with (n=21) and without (n=28) mutations in PIK3CA gene (PIK3CA^mut^ vs PIK3CA^wt.^) The Y axis represents the probability of OS of patients belonging to different groups and X axis represents the time of survival (in months). **(B)** Box plot representing mean expression (based on TPM values of RNA-seq) and **(C)** Relative expression (based on qRT-PCR) of viral oncoprotein HPV16 E7 in PIK3CA^mut^ vs PIK3CA^wt^ tumors. Relative gene expression reflects expression of E7 minus expression of the house-keeping gene (18srRNA). The data is represented as mean ± SD. **(D)** Box plot representing HPV16 viral load (natural log transformed) and **(E)** total TMB/Mb of the genome in PIK3CA^mut^ vs PIK3CA^wt^ tumors. “**” represents significant p value < 0.01 (two-tailed Students’ t-test) TMB: Tumor Mutation Burden; PIK3CA^mut:^ PIK3CA-mutant tumors; PIK3CA^wt^: PIK3CA-wild tumors

**Supplementary Figure S3-PIK3CA mutational landscape and its association with overall survival (OS) in HPV16-positive TCGA-CESC patients (A)** Lollipop plot representing the overview of mutations in PIK3CA in HPV16 positive TCGA-CESC cohort. *The plot was downloaded from cBioportal*. **(B)** Kaplan-Meier plot depicting the patient OS of patient cohort with and without mutations in PIK3CA gene. The Y axis represents the probability of OS of patients belonging to different groups and X axis represents the time of survival (in months).

**Supplementary Figure S4-Differential expression of immune-related genes based on PIK3CA mutation status:** Scatter dot plots portraying the expression levels (TPM) of **(A)** IL2, **(B)** CD40LG**, (C)** IL24, and **(D)** IL12B in normal cervical tissues (Normal), PIK3CA-wild tumors (PIK3CA^wt^), and PIK3CA-mutant tumors (PIK3CA^mut^). Statistically significant differences between groups are indicated by the corresponding p values above the plots. P (t-test) < 0.05 considered significant.

**Supplementary Figure S5. Sample-wise correlation plots depicting the correlation between expression levels (TPM) of CCL3 with cytotoxic T-cell markers: (A–C)** CD8A, **(D-F)** GZMB, **(G–I)** PRF1 and **(J–L)** IFNG in normal cervical tissues (Normal), PIK3CA-wild tumors (PIK3CA^wt^), and PIK3CA-mutant tumors (PIK3CA^mut^) respectively. The red line indicates the linear regression fit, while the black line represents the 95% confidence interval. Correlation coefficients (*r*) and corresponding *p* values were calculated using Pearson’s correlation analysis.

**Supplementary Figure S6-Sample-wise correlation plots depicting the correlation between expression levels (TPM) of CCL3 with interferon-γ–related immune signatures: (A–C)** STAT1, **(D–F)** IRF1, **(G–I)** CXCL9, and **(J–L)** CXCL10 in normal cervical tissues (Normal), PIK3CA-wild tumors (PIK3CA^wt^), and PIK3CA-mutant tumors (PIK3CA^mut^) respectively in each panel. The red line indicates the linear regression fit, while the black line represents the 95% confidence interval. Correlation coefficients (*r*) and corresponding *p* values were calculated using Pearson’s correlation analysis.

**Supplementary Figure S7-Sample-wise correlation plots depicting the correlation between expression levels (TPM) of CCL3 with myeloid lineage markers: (A–C)** CD68, **(D–F)** CD163, and **(G–I)** CSF1R in normal cervical tissues (Normal), PIK3CA-wild tumors (PIK3CA^wt^), and PIK3CA-mutant tumors (PIK3CA^mut^) respectively in each panel. The red line indicates the linear regression fit, while the black line represents the 95% confidence interval. Correlation coefficients (*r*) and corresponding *p* values were calculated using Pearson’s correlation analysis.

**Supplementary Figure S8-Sample-wise correlation plots depicting the correlation between expression levels (TPM) of CCL3 with Treg-associated markers: (A–C)** FOXP3 and **(D–F)** IL2RA in normal cervical tissues (Normal), PIK3CA-wild tumors (PIK3CA^wt^), and PIK3CA-mutant tumors (PIK3CA^mut^) respectively in each panel. The red line indicates the linear regression fit, while the black line represents the 95% confidence interval. Correlation coefficients (*r*) and corresponding *p* values were calculated using Pearson’s correlation analysis.

## Supplementary Tables

**Supplementary Table S1:** Clinicopathological characteristics of patients (n=49) involved in the study

**Supplementary Table S2:** Details of coverage and insert size for all samples (n=49) subjected to Whole Exome Sequencing (WES)

**Supplementary Table S3:** Details of sequencing quality metrices for all samples (n=29) included in total RNA sequencing

**Supplementary Table S4:** Gene signatures for classification of Tertiary Lymphoid Structures (TLS) maturity

**Supplementary Table S5:** Details of different types of mutations in each HPV16-positive CaCx patients (n=49)

**Supplementary Table S6:** Summary statistics of the mutations recorded in HPV16-positive CaCx patients (n=49)

**Supplementary Table S7:** Significant Kegg Pathways associated with the mutated genes having functional impact on the protein function

**Supplementary Table S8:** Significant GO-BP terms associated with the mutated genes having functional impact on the protein function

**Supplementary Table S9:** Non-synonymous protein coding somatic mutations identified in known Significantly Mutated Genes (SMGs) and driver genes in HPV16-positive CaCx patients of Indian cohort

**Supplementary Table S10:** Comparison of the frequency of mutations in the known Significantly Mutated Genes (SMGs) and driver genes between HPV16-positive CaCx patients of Indian cohort and TCGA-CESC PanCanAtlas Cohort

**Supplementary Table S11:** Details of different PIK3CA mutations observed in HPV16-positive PIK3CA-mutant CaCx patients (n=21)

**Supplementary Table S12:** Degree of connectivity of Significantly Mutated Genes (SMGs) in protein-protein interaction network based on STRING database

**Supplementary Table S13:** Comparison of the clinical characteristics between PIK3CA-mutant (n=21) and PIK3CA-wild (n=28) tumors in HPV16-positive CaCx cohort

**Supplementary Table S14:** Details of the PIK3CA mutation status in HPV16-positive TCGA-CESC patients profiled for Whole Exome Sequencing (n=103)

**Supplementary Table S15:** Association of clinical parameters with PIK3CA-mutant (n=27) and PIK3CA-wild tumors (n=76) in HPV16-positive TCGA-CESC cohort

**Supplementary Table S16:** List of Differentially Expressed Genes (DEGs) in *PIK3CA-mutant* tumors (n=17) compared to *PIK3CA-wild* tumors (n=12) in Indian HPV16-positive CaCx cohort, portraying expression difference of fold change cut-off |log2(FC) | > 0.6 and False Discovery Rate (FDR) corrected p<0.05

## Abbreviations

BAM: Binary Alignment Map
CaCx: Cervical Cancer
CESC: Cervical squamous cell carcinoma and endocervical adenocarcinoma
CIBERSORT: Cell-type Identification by Estimating Relative Subsets of RNA Transcripts
DAVID: Database for Annotation, Visualisation and Integrated Discovery
DEG: Differentially Expressed Gene
FDA: Food and Drug Administration
FDR: False Discovery Rate
FIGO: International Federation of Gynecology and Obstetrics
GDC: Genomic Data Commons
GLOBOCON: Global Cancer Observatory
GO-BP: Gene Ontology-Biological Processes
HPV: Human Papillomavirus
IGV: Integrative Genomics Viewer
INDELS: Insertions and deletions
LMICs: Low and Middle income countries
PPI: Protein-Protein Interaction
RIN: RNA Integrity Number
SIFT: Sorting Intolerant from Tolerant
SMGs: Significantly Mutated Genes
SNVs: Single Nucleotide Variations
STRING: Search Tool for the Retrieval of Interacting Genes/Proteins
TCGA: The Cancer Genome Atlas
TIICs: Tumor Infiltrating Immune Cells
TIME: Tumor Immune microenvironment
TLS: Tertiary Lymphoid Structures
TMB: Tumor Mutation Burden
TPM: Transcripts Per Million
WES: Whole Exome Sequencing

