## Supplementary material for "Integrated Genomic and Immune Profiling Reveals PIK3CA-Mutant HPV16-Positive Cervical Cancer as a Clinically Aggressive and Immunologically Distinct Subtype in Patients from Eastern India": Supplemtary Tables

**Supplementary Table S1: Clinicopathological characteristics of patients involved in the study**

| Clinical parameters | | # patients | % of patients |
|---|---|---|---|
| **FIGO Stage** | Early (<IIB) | 11 | 22.44 |
| | Late (≥IIB) | 38 | 77.55 |
| **Lymph node involvement** | Pelvic Lymph node involved (pN+) | 24 | 50.00 |
| | Pelvic Lymph node not involved (pN-) | 24 | 50.00 |
| **Histopathology** | Squamous Cell Carcinoma (SCC) | 41 | 87.23 |
| | Adenocarcinoma | 5 | 10.64 |
| | Adenosquamous | 1 | 2.13 |
| **Histological Grade** | Moderately differentiated | 27 | 57.45 |
| | Poorly differentiated | 7 | 14.89 |
| | Well differentiated | 3 | 6.38 |
| | Not known | 10 | 21.28 |

**Supplementary Table S2: Details of coverage and insert size for all samples (n=49) subjected to Whole Exome Sequencing (WES)**

| Sample ID | Mean coverage (X) | | Median insert size (bp) | |
|---|---|---|---|---|
| | Tumor tissue | Blood | Tumor tissue | Blood |
| 17-369 | 106.65 | 131.84 | 155 | 153 |
| 17-397 | 109.1 | 124.71 | 155 | 156 |
| 17-415 | 94.26 | 108.82 | 173 | 184 |
| 17-417 | 155.86 | 123.4 | 156 | 153 |
| 17-491 | 70.23 | 109.42 | 174 | 185 |
| 17-499 | 141.52 | 136.98 | 158 | 153 |
| 17-507 | 132.53 | 120.34 | 166 | 165 |
| 17-555 | 127.95 | 129.85 | 159 | 160 |
| 17-574 | 95.18 | 106.96 | 190 | 173 |
| 17-580 | 140.44 | 145.88 | 167 | 170 |
| 17-590 | 114.61 | 139.08 | 155 | 155 |
| 17-599 | 132.34 | 142.18 | 154 | 153 |
| 17-607 | 114.46 | 96.65 | 180 | 178 |
| 17-633 | 141.67 | 122.09 | 165 | 165 |
| 17-641 | 131.31 | 194.32 | 152 | 160 |
| 17-652 | 190.23 | 211.34 | 165 | 170 |
| 17-673 | 150.64 | 161.84 | 156 | 158 |
| 17-736 | 127.72 | 160.28 | 158 | 159 |
| 17-782 | 143.82 | 143.18 | 158 | 158 |
| 17-819 | 96.76 | 159.52 | 160 | 164 |
| 17-857 | 117.89 | 158.91 | 157 | 159 |
| 18-335 | 139.2 | 155.11 | 158 | 160 |
| 18-336 | 72.46 | 91.35 | 159 | 175 |
| 18-354 | 97.82 | 113.67 | 158 | 157 |
| 18-373 | 100.78 | 125.52 | 156 | 160 |
| 18-636 | 139.01 | 167.23 | 165 | 165 |
| 18-815 | 132.11 | 152.94 | 158 | 157 |
| 18-932 | 135.21 | 141.42 | 160 | 156 |
| 18-1062 | 85.27 | 97.71 | 175 | 177 |
| 18-1102 | 137.84 | 145.88 | 163 | 162 |
| 18-1151 | 142.33 | 147.46 | 162 | 160 |
| 18-1201 | 119.61 | 133.16 | 163 | 164 |
| 18-1291 | 128.03 | 131.31 | 154 | 158 |
| 18-1861 | 118.46 | 104.69 | 156 | 160 |
| 18-2081 | 114.46 | 119.62 | 162 | 160 |
| 18-2158 | 112.93 | 130.87 | 157 | 160 |
| 18-2178 | 139 | 111.01 | 162 | 159 |
| 18-2302 | 123.34 | 114.58 | 156 | 156 |
| 18-2312 | 131.7 | 131.51 | 164 | 165 |

| | | | | |
|---|---|---|---|---|
| 18-2316 | 117.43 | 125.68 | 156 | 163 |
| 19-003 | 127.67 | 94.04 | 160 | 156 |
| 19-042 | 131.28 | 114.58 | 158 | 157 |
| 19-078 | 113.78 | 131.27 | 154 | 157 |
| 19-253 | 114.04 | 142.88 | 159 | 159 |
| 19-309 | 129.61 | 124.19 | 157 | 156 |
| 19-780 | 101.53 | 126.48 | 160 | 160 |
| 19-974 | 99.9 | 102.47 | 161 | 158 |
| 19-1042 | 114.43 | 105.52 | 158 | 158 |
| 19-1236 | 117.74 | 115.09 | 164 | 160 |

**Supplementary Table S3: Details of sequencing quality metrices for all samples (n=29) included in total RNA sequencing**

| Sample ID | RNA-seq sample ID | Total number of sequenced reads | Total number of uniquely mapped reads (GRCh37) | RNA Integrity Number (RIN) | Ratio of exon-mapped reads to total uniquely mapped reads (Expression Profile Efficiency) (GRCh37, Ensemble_v87) | Total number of detected transcripts [genes] with reads ≥ 1 [total number of genes (ensembl.v87) = 55638] |
|---|---|---|---|---|---|---|
| 18-1861 | 109 | 65287385 | 58200441 | 7.3 | 0.619 | 41567 |
| 18-2158 | 114 | 54865942 | 48538702 | 8.2 | 0.793 | 32206 |
| 18-2081 | 118 | 197199661 | 177503079 | 7 | 0.851 | 34247 |
| 18-2312 | 131 | 174661264 | 159544695 | 6.5 | 0.615 | 40465 |
| 19-078 | 135 | 177125851 | 160360533 | 8.1 | 0.811 | 38903 |
| 19-253 | 143 | 54021319 | 47441633 | 8.1 | 0.727 | 31876 |
| 19-780 | 144 | 60865383 | 53554893 | 8.1 | 0.839 | 31749 |
| 19-1042 | 147 | 128067988 | 116647473 | 6.9 | 0.702 | 43431 |
| 19-1236 | 148 | 178370875 | 159750526 | 7.4 | 0.738 | 39998 |
| 17-397 | 17-397 | 84573947 | 69727734 | 7.4 | 0.785 | 32113 |
| 17-417 | 17-417 | 72037683 | 46277527 | 7 | 0.783 | 28862 |
| 17-491 | 17-491 | 47852894 | 17100794 | 6.7 | 0.417 | 24941 |
| 17-499 | 17-499 | 65100117 | 33949555 | 7.5 | 0.633 | 29231 |
| 17-580 | 17-580 | 84469811 | 67456478 | 8 | 0.753 | 31992 |
| 17-590 | 17-590 | 93777517 | 78957240 | 8.5 | 0.834 | 31181 |
| 17-599 | 17-599 | 96189519 | 42146322 | 8.3 | 0.573 | 28657 |
| 17-633 | 17-633 | 49929082 | 29165923 | 8.6 | 0.731 | 26597 |
| 17-641 | 17-641 | 47761144 | 24813303 | 7.8 | 0.662 | 25143 |
| 17-652 | 17-652 | 88187381 | 46484328 | 7.6 | 0.613 | 27983 |
| 17-736 | 17-736 | 49153118 | 17884269 | 8.1 | 0.447 | 23676 |
| 17-782 | 17-782 | 83153927 | 57124294 | 8.7 | 0.818 | 29136 |
| 17-857 | 17-857 | 57360379 | 46789723 | 7.5 | 0.770 | 31464 |
| 18-1062 | 18-1062 | 103973584 | 53345812 | 8.5 | 0.688 | 28552 |
| 18-335 | 18-335 | 58305378 | 36864999 | 8.4 | 0.749 | 28362 |
| 18-373 | 18-373 | 121202000 | 97402532 | 7.4 | 0.741 | 33773 |
| 18-636 | 18-636 | 82551531 | 56350662 | 8.4 | 0.802 | 29032 |
| 18-815 | 18-815 | 104862467 | 60473302 | 7.8 | 0.666 | 31115 |
| 18-1102 | 96 | 71235281 | 59232908 | 8.3 | 0.775 | 38202 |
| 18-1201 | 99 | 56999332 | 49407295 | 7 | 0.575 | 43068 |

**Supplementary Table S4: Gene signatures for classification of Tertiary Lymphoid Structures (TLS) maturity**

| Gene names | Method of identification | Functional role in TLS formation | TLS maturity stage | Reference |
|---|---|---|---|---|
| *CCL2* | mRNA Microarray analysis | Chemokine involved in monocyte recruitment | Early | [17], [20] |
| *CCL3* | mRNA Microarray analysis | Chemokine involved in T cell and monocyte recruitment | Early | [17], [20] |
| *CCL4* | mRNA Microarray analysis | Chemokine involved in T cell and monocyte recruitment | Early | [17], [20] |
| *CCL5* | mRNA Microarray analysis | Chemokine involved in T cell recruitment | Early | [17], [20] |
| *CCL8* | mRNA Microarray analysis | Chemokine involved in monocyte recruitment | Early | [17], [20] |
| *CCL18* | mRNA Microarray analysis | Chemokine involved in attracting naïve T cells | Early | [17], [20] |
| *CCL19* | mRNA Microarray analysis | Chemokine involved in dendritic cell and T cell recruitment | Intermediate | [17], [20] |
| *CCL21* | mRNA Microarray analysis | Chemokine involved in dendritic cell and T cell recruitment | Intermediate | [17], [20] |
| *CXCL9* | mRNA Microarray analysis | Chemokine involved in T cell recruitment | Early | [17], [20] |
| *CXCL10* | mRNA Microarray analysis | Chemokine involved in T cell recruitment | Early | [17], [20] |
| *CXCL11* | mRNA Microarray analysis | Chemokine involved in T cell recruitment | Early | [17], [20] |
| *CXCL12* | mRNA Microarray analysis | Chemokine involved in B cell recruitment | Intermediate | [17] |

| | | | | |
|---|---|---|---|---|
| *CXCL13* | mRNA Microarray analysis/qRT-PCR/ RNA seq analysis | Chemokine involved in B cell recruitment | All Stages | [18], [19], [20] |
| *CCL20* | mRNA Microarray analysis | Chemokine involved in Th17 cell recruitment | Intermediate | [17] |
| *CCR5* | mRNA Microarray analysis | Receptor for CCL3, CCL4, and CCL5; involved in T cell migration | Intermediate | [17] |
| *CCR6* | RNA seq analysis | Receptor for CCL20; involved in Th17 cell migration | Intermediate | [17] |
| *CXCR3* | mRNA Microarray analysis | Receptor for CXCL9, CXCL10, and CXCL11; involved in T cell migration | Intermediate | [17] |
| *CD4* | mRNA Microarray analysis | Marker for helper T cells | All Stages | [17], [22] |
| *IL-1R1* | mRNA Microarray analysis | Receptor for IL-1; involved in inflammatory response | Early | [17] |
| *IL-1R2* | mRNA Microarray analysis | Decoy receptor for IL-1; regulates inflammatory response | Early | [17] |
| *IL-10* | mRNA Microarray analysis | Anti-inflammatory cytokine; regulates immune response | Intermediate | [17] |
| *CSF2* | mRNA Microarray analysis | Cytokine involved in dendritic cell maturation | Intermediate | [17] |
| *TRAF6* | mRNA Microarray analysis | Adapter protein in TLR and cytokine signaling | Early | [17], [22] |
| *STAT5A* | mRNA Microarray analysis | Transcription factor involved in cytokine signaling | Intermediate | [17] |
| *LUM* | Spatial transcriptomics | Extracellular matrix protein; involved in tissue remodeling | Intermediate | [17] |
| *C1QA* | Spatial transcriptomics | Complement component; involved in immune complex clearance | Intermediate | [17] |
| *C7* | Spatial transcriptomics | Complement component; involved in membrane attack complex formation | Intermediate | [17] |
| *CD52* | Spatial transcriptomics | Surface protein on lymphocytes; involved in immune regulation | All Stages | [17] |

| | | | | |
|---|---|---|---|---|
| *APOE* | Spatial transcriptomics | Lipid transport protein; involved in immune modulation | Intermediate | [17] |
| *PTGDS* | RNA seq analysis | Enzyme involved in prostaglandin synthesis; modulates inflammation | Intermediate | [17] |
| *RBP5* | RNA seq analysis | Retinol-binding protein; involved in vitamin A transport | Intermediate | [17] |
| *CD1D* | RNA seq analysis | Antigen-presenting molecule; presents lipid antigens to NKT cells | Intermediate | [17] |
| *CD5* | mRNA Microarray analysis | Surface protein on T and B cells; modulates signaling | All Stages | [17], [22] |
| *CD38* | mRNA Microarray analysis | Surface protein on activated lymphocytes; involved in cell adhesion | Intermediate | [17], [22] |
| *CD40* | mRNA Microarray analysis | Co-stimulatory protein on B cells; essential for their activation | Intermediate | [17], [22] |
| *IL2RA* | mRNA Microarray analysis | Alpha chain of IL-2 receptor; involved in T cell proliferation | Intermediate | [17] |
| *IL7R* | Spatial transcriptomics | Receptor for IL-7; involved in lymphocyte development | Early | [17] |
| *CD200* | qRT-PCR | Surface glycoprotein; delivers inhibitory signals to immune cells | Intermediate | [17], [18] |
| *ICOS* | qRT-PCR | Co-stimulatory molecule on T cells; important for T cell activation | Intermediate | [17], [18] |
| *LAT* | RNA seq analysis | Adapter protein in T cell receptor signaling | Early | [17] |
| *SKAP1* | RNA seq analysis | Adapter protein in T cell receptor signaling | Early | [17] |
| *SGPP2* | qRT-PCR | Enzyme involved in sphingolipid metabolism; modulates immune responses | Intermediate | [17], [18], [22] |
| *SH2D1A* | qRT-PCR | Adapter protein in T and NK cells; involved in signaling | Intermediate | [17], [18], [22] |

| | | | | |
|---|---|---|---|---|
| *TIGIT* | qRT-PCR | Inhibitory receptor on T cells; regulates immune responses | Intermediate | [17], [18] |
| *PDCD1* | qRT-PCR | Programmed cell death protein 1; inhibitory receptor on T cells | Intermediate | [17], [18], [22] |
| *IGSF6* | mRNA Microarray analysis | Immunoglobulin superfamily member; function not well characterized | Intermediate | [17] |
| *CD79A* | Spatial transcriptomics | Part of B cell receptor complex; essential for B cell activation | Intermediate | [17] |
| *CD79B* | RNA seq analysis | Part of B cell receptor complex; essential for B cell activation | Intermediate | [17] |
| *FBLN7* | qRT-PCR | Extracellular matrix protein; involved in cell adhesion | Intermediate | [17], [18] |
| *PIM2* | Spatial transcriptomics | Serine/threonine kinase; involved in cell survival and proliferation | Intermediate | [17] |
| *DERL3* | Spatial transcriptomics | Involved in endoplasmic reticulum-associated degradation | Intermediate | [17] |
| *EIF1AY* | RNA seq analysis | Translation initiation factor; involved in protein synthesis | Intermediate | [17] |
| *CETP* | RNA seq analysis | Cholesteryl ester transfer protein; involved in lipid transport | Intermediate | [17] |
| *PLTP* | Spatial transcriptomics | Phospholipid transfer protein; involved in lipid metabolism | Intermediate | [17] |
| *SSR4* | Spatial transcriptomics | Signal sequence receptor subunit; involved in protein translocation | Intermediate | [17] |
| *XBP1* | Spatial transcriptomics | Transcription factor; involved in unfolded protein response | Intermediate | [17] |
| *SDC1* | mRNA Microarray analysis | Syndecan-1; involved in cell-cell and cell-matrix interactions | Intermediate | [17], [22] |
| *MS4A1* | mRNA Microarray analysis | Also known as CD20; B cell surface molecule | Intermediate | [17], [22] |

| | | | | |
|---|---|---|---|---|
| *TNFRSF17* | NanoString gene expression analysis | Also known as BCMA; receptor on plasma cells | Mature | [17], [21] |
| *IGJ* | NanoString gene expression analysis | Joining chain of multimeric immunoglobulins | Mature | [17] |
| *IGHG1* | Spatial transcriptomics | Immunoglobulin heavy constant gamma 1 | Mature | [17] |
| *IGHG2* | Spatial transcriptomics | Immunoglobulin heavy constant gamma 2 | Mature | [17] |
| *IGHG3* | Spatial transcriptomics | Immunoglobulin heavy constant gamma 3 | Mature | [17] |
| *IGHG4* | Spatial transcriptomics | Immunoglobulin heavy constant gamma 4 | Mature | [17] |
| *IGHM* | Spatial transcriptomics | Immunoglobulin heavy constant mu | Mature | [17] |
| *IGHA1* | Spatial transcriptomics | Immunoglobulin heavy constant alpha 1 | Mature | [17] |
| *IGHGP* | Spatial transcriptomics | Immunoglobulin heavy constant gamma pseudogene | Mature | [17] |
| *IGKC* | Spatial transcriptomics | Immunoglobulin kappa constant | Mature | [17] |
| *IGLC1* | Spatial transcriptomics | Immunoglobulin lambda constant 1 | Mature | [17] |
| *IGLC2* | Spatial transcriptomics | Immunoglobulin lambda constant 2 | Mature | [17] |
| *IGLC3* | Spatial transcriptomics | Immunoglobulin lambda constant 3 | Mature | [17] |
| *FCRL5* | Spatial transcriptomics | Fc receptor-like protein 5; involved in B cell regulation | Mature | [17] |
| *MZB1* | Spatial transcriptomics | Marginal zone B and B1 cell-specific protein; involved in plasma cell differentiation | Mature | [17] |
| *GFI1* | mRNA Microarray analysis | Transcriptional repressor; involved in hematopoiesis | Intermediate | [17] |
| *TRBC2* | Spatial transcriptomics | T cell receptor beta constant 2 | Intermediate | [17] |

***N.B-*** Reference numbers are same as main text list

**Supplementary Table S5: Details of different types of mutations in each HPV16-positive CaCx patients (n=49)**

| patient ID | Missense Mutation | In Frame Del | Frame Shift Del | Nonsense Mutation | Silent | Splice Site | Frame Shift Ins | De novo Start InFrame | De novo Start OutOfFrame | Start Codon SNP | In Frame Ins | Nonstop Mutation | Stop Codon Del | Total |
|---|---|---|---|---|---|---|---|---|---|---|---|---|---|---|
| 17-369 | 34 | 2 | 2 | 2 | 15 | 1 | 0 | 0 | 0 | 0 | 0 | 0 | 0 | 56 |
| 17-397 | 165 | 2 | 52 | 10 | 59 | 4 | 6 | 1 | 0 | 0 | 0 | 0 | 0 | 299 |
| 17-415 | 67 | 0 | 5 | 6 | 28 | 5 | 2 | 0 | 0 | 0 | 0 | 0 | 0 | 113 |
| 17-417 | 65 | 1 | 4 | 4 | 38 | 9 | 1 | 0 | 0 | 0 | 0 | 0 | 0 | 122 |
| 17-491 | 18 | 0 | 0 | 1 | 4 | 1 | 0 | 0 | 0 | 0 | 0 | 0 | 0 | 24 |
| 17-499 | 39 | 0 | 1 | 2 | 16 | 1 | 0 | 0 | 0 | 0 | 0 | 0 | 0 | 59 |
| 17-507 | 0 | 1 | 0 | 0 | 0 | 0 | 0 | 0 | 0 | 0 | 0 | 0 | 0 | 1 |
| 17-555 | 1 | 1 | 0 | 0 | 2 | 0 | 0 | 0 | 0 | 0 | 0 | 0 | 0 | 4 |
| 17-574 | 0 | 1 | 0 | 0 | 1 | 0 | 0 | 0 | 0 | 0 | 0 | 0 | 0 | 2 |
| 17-580 | 46 | 1 | 1 | 1 | 20 | 1 | 1 | 0 | 0 | 0 | 0 | 0 | 0 | 71 |
| 17-590 | 124 | 1 | 3 | 5 | 48 | 4 | 0 | 0 | 0 | 0 | 0 | 0 | 0 | 185 |
| 17-599 | 142 | 2 | 0 | 9 | 55 | 2 | 0 | 0 | 0 | 0 | 0 | 0 | 0 | 210 |
| 17-607 | 32 | 0 | 2 | 5 | 18 | 1 | 1 | 0 | 0 | 0 | 0 | 0 | 0 | 59 |
| 17-633 | 443 | 2 | 5 | 49 | 209 | 11 | 1 | 0 | 1 | 1 | 0 | 0 | 0 | 722 |
| 17-641 | 74 | 0 | 1 | 3 | 30 | 4 | 1 | 0 | 0 | 0 | 0 | 0 | 0 | 113 |
| 17-652 | 49 | 0 | 2 | 6 | 41 | 2 | 1 | 0 | 0 | 0 | 0 | 0 | 0 | 101 |
| 17-673 | 78 | 2 | 2 | 3 | 26 | 4 | 1 | 0 | 1 | 0 | 1 | 0 | 0 | 118 |
| 17-736 | 28 | 1 | 1 | 4 | 14 | 2 | 2 | 0 | 0 | 0 | 0 | 0 | 0 | 52 |
| 17-782 | 108 | 0 | 4 | 12 | 51 | 0 | 4 | 0 | 0 | 0 | 0 | 0 | 0 | 179 |
| 17-819 | 80 | 3 | 2 | 2 | 34 | 1 | 1 | 0 | 0 | 0 | 0 | 1 | 0 | 124 |
| 17-857 | 6 | 1 | 1 | 0 | 4 | 0 | 1 | 0 | 0 | 0 | 0 | 0 | 0 | 13 |
| 18-335 | 99 | 4 | 10 | 8 | 36 | 5 | 2 | 0 | 0 | 0 | 0 | 0 | 0 | 164 |
| 18-336 | 120 | 0 | 3 | 13 | 47 | 7 | 0 | 0 | 0 | 2 | 1 | 0 | 0 | 193 |
| 18-354 | 63 | 0 | 1 | 7 | 20 | 4 | 1 | 0 | 0 | 0 | 1 | 0 | 0 | 97 |
| 18-373 | 5 | 2 | 0 | 0 | 4 | 3 | 0 | 0 | 0 | 0 | 0 | 0 | 0 | 14 |
| 18-636 | 37 | 2 | 2 | 4 | 18 | 4 | 3 | 0 | 0 | 0 | 0 | 0 | 0 | 70 |
| 18-815 | 11 | 0 | 1 | 1 | 3 | 0 | 0 | 0 | 0 | 0 | 0 | 0 | 0 | 16 |
| 18-932 | 14 | 3 | 0 | 2 | 9 | 0 | 1 | 0 | 0 | 0 | 1 | 0 | 0 | 30 |
| 18-1062 | 114 | 0 | 3 | 11 | 41 | 5 | 0 | 0 | 0 | 0 | 0 | 0 | 0 | 174 |
| 18-1102 | 137 | 2 | 0 | 7 | 52 | 5 | 1 | 1 | 0 | 0 | 0 | 0 | 0 | 205 |
| 18-1151 | 2 | 1 | 0 | 0 | 0 | 0 | 0 | 0 | 0 | 0 | 0 | 0 | 0 | 3 |
| 18-1201 | 69 | 1 | 1 | 1 | 36 | 1 | 0 | 0 | 0 | 0 | 0 | 0 | 0 | 109 |
| 18-1291 | 207 | 2 | 1 | 19 | 73 | 9 | 0 | 0 | 0 | 0 | 0 | 0 | 0 | 311 |
| 18-1861 | 190 | 1 | 2 | 13 | 59 | 3 | 2 | 0 | 0 | 1 | 0 | 0 | 0 | 271 |
| 18-2081 | 43 | 2 | 0 | 10 | 18 | 3 | 0 | 0 | 0 | 0 | 0 | 0 | 0 | 76 |
| 18-2158 | 61 | 1 | 1 | 7 | 17 | 4 | 3 | 0 | 0 | 0 | 0 | 0 | 0 | 94 |
| 18-2178 | 60 | 1 | 1 | 9 | 31 | 4 | 2 | 0 | 0 | 0 | 1 | 0 | 0 | 109 |

| | | | | | | | | | | | | | |
|---|---|---|---|---|---|---|---|---|---|---|---|---|---|
| 18-2302 | 58 | 0 | 2 | 5 | 24 | 3 | 0 | 0 | 0 | 0 | 0 | 0 | 0 | 92 |
| 18-2312 | 29 | 0 | 1 | 4 | 12 | 1 | 2 | 0 | 0 | 0 | 0 | 0 | 1 | 50 |
| 18-2316 | 0 | 1 | 0 | 0 | 0 | 0 | 0 | 0 | 0 | 0 | 0 | 0 | 0 | 1 |
| 19-003 | 46 | 5 | 3 | 8 | 25 | 0 | 1 | 1 | 0 | 0 | 1 | 0 | 0 | 90 |
| 19-042 | 19 | 2 | 0 | 2 | 14 | 0 | 0 | 1 | 0 | 0 | 0 | 0 | 0 | 38 |
| 19-078 | 833 | 4 | 3 | 66 | 349 | 21 | 1 | 0 | 0 | 1 | 0 | 2 | 0 | 1280 |
| 19-253 | 123 | 2 | 2 | 16 | 63 | 3 | 0 | 0 | 0 | 0 | 1 | 0 | 0 | 210 |
| 19-309 | 1 | 0 | 0 | 0 | 1 | 0 | 0 | 0 | 0 | 0 | 0 | 0 | 0 | 2 |
| 19-780 | 12 | 0 | 0 | 1 | 2 | 1 | 0 | 0 | 0 | 0 | 0 | 0 | 0 | 16 |
| 19-974 | 13 | 2 | 1 | 2 | 9 | 0 | 0 | 0 | 0 | 0 | 0 | 0 | 0 | 27 |
| 19-1042 | 14 | 1 | 0 | 1 | 7 | 1 | 0 | 0 | 0 | 0 | 0 | 0 | 0 | 24 |
| 19-1236 | 45 | 1 | 2 | 2 | 22 | 2 | 1 | 0 | 0 | 0 | 0 | 0 | 0 | 75 |
| Total | 4024 | 61 | 128 | 343 | 1705 | 142 | 43 | 4 | 2 | 5 | 7 | 3 | 1 | 6468 |

**Supplementary Table S6: Summary statistics of the mutations recorded in HPV16 positive CaCx patients (n=49)**

| Statistic | Minimum | Maximum | Mean | Median |
|---|---|---|---|---|
| Total No. of mutations | 1 | 1280 | 132 | 90 |
| Total no. of mutations, excluding synonymous mutations | 1 | 931 | 97.2 | 60 |
| Ratio of non-synonymous to synonymous mutations | 1 | 7 | 2.83 | 2.701 |
| Mutation rate per Mb (Total) | 0 | 44.9 | 4.66 | 3.01 |
| Mutation rate per Mb, excluding synonymous mutations | 0 | 32.4 | 3.22 | 1.94 |

**Supplementary Table S7: Significant Kegg Pathways associated with the mutated genes having functional impact on the protein function**

| Kegg pathway ID:term | Gene Count | Genes | FDR |
|---|---|---|---|
| hsa04814: Motor proteins | 51 | *KIF17, ACTB, KIF15, ACTR1A, KIF5C, KIF5A, KIF13B, KIF21A, KIF21B, KIF1A, DYNC2H1, TPM4, TPM3, MYH15, TNNC1, DYNLL1, TUBG1, MYL4, MYH1, MYO3B, TNNT1, KIFC1, MYO3A, MYH8, MYH4, KIF20B, MYH6, DNAH1, DNAH2, DNAH7, TUBAL3, DNAH8, DNAH5, DNAH6, DNAH9, KLC1, MYO6, MYH13, MYH14, DNAI1, MYH7B, DYNC1H1, DNAH17, DNAH14, MYO16, MYO1E, CENPE, ACTC1, KIF26B, KIF4A, MYO5C* | 2.50E-05 |
| hsa05412: Arrhythmogenic right ventricular cardiomyopathy | 28 | *RYR2, LAMA2, ITGB4, LAMA1, ATP2A3, ATP2A2, CACNA1C, CACNA1F, SLC8A1, ACTB, CTNNA1, DMD, CTNNA3, CTNNA2, CACNG3, SGCG, DSP, TCF7L1, JUP, ACTN2, CACNA2D3, CACNB1, SLC8A3, DES, ITGA8, ITGA7, ITGA5, DSC2* | 2.50E-05 |
| hsa04510: Focal adhesion | 51 | *MYLK2, ITGB4, PTEN, ARHGAP5, ELK1, ACTB, ARHGAP35, IGF1R, MYLK, AKT3, KDR, AKT1, TNR, VAV3, PPP1R12A, VWF, PRKCB, ACTN1, COL2A1, COL4A2, PIK3CA, ITGA8, COL4A5, COL6A3, ITGA7, COL6A6, COL6A5, ITGA5, LAMA2, ROCK1, LAMA1, PXN, LAMA3, XIAP, PIK3R1, THBS1, EGFR, RELN, ERBB2, FLNA, MAPK1, FLNC, PAK2, LAMB3, LAMB2, BRAF, LAMB1, COL1A1, COL1A2, COL9A3, BCAR1* | 6.78E-05 |
| hsa05165: Human papillomavirus infection | 72 | *RB1, ITGB4, PTEN, UBE3A, IKBKB, CASP8, PPP2R1B, HEY1, CASP3, AKT3, AKT1, TNR, EP300, MAGI1, APC2, PRKCI, VWF, HLA-B, HLA-C, TSC1, HLA-A, WNT16, HLA-F, TUBG1, CREB3, COL2A1, COL4A2, PIK3CA, CCNE1, ITGA8, COL4A5, COL6A3, ITGA7, COL6A6, COL6A5, ITGA5, MAML3, ATP6V1B1, RBPJL, NOTCH1, LAMA2, LAMA1, NOTCH4, PXN, LAMA3, PIK3R1, PTGS2, THBS1, EGFR, RELA, NRAS, RELN, DVL3, MAPK1, BAK1, FZD1, TCF7L1, FZD5, LAMB3, LAMB2, STAT2, MX1, LAMB1, MTOR, COL1A1, DLG2, COL1A2, APC, GNAS, MDM2, COL9A3, ATR* | 8.24E-05 |
| hsa04512: ECM-receptor interaction | 28 | *LAMA2, ITGB4, LAMA1, LAMA3, THBS1, SV2C, RELN, TNR, LAMB3, VWF, LAMB2, GP1BB, LAMB1, HSPG2, COL1A1, COL2A1, FRAS1, COL1A2, COL4A2, ITGA8, COL4A5, COL6A3, ITGA7, COL9A3, COL6A6, COL6A5, ITGA5, FREM1* | 2.53E-04 |
| hsa05200: Pathways in cancer | 99 | *RB1, IL5RA, CALML3, ETS1, ELK1, IGF1R, FGF9, CDH1, AKT3, AKT1, EP300, SKP2, TPM3, DAPK1, PRKCB, RUNX1, COL4A2, MSH2, MSH3, CCNE1, AGTR1, COL4A5, CSF1R, NOTCH1, EPAS1, NOTCH4, CUL2, MGST1, LPAR4, PIK3R1, HSP90B1, TPR, DVL3, PLCG1, FZD1, ZBTB17, GSTM3, SMAD3, JUP, FZD5, BRAF, NFKB2, BMP2, LPAR5, GNB1, MDM2, GNAS, CALM3, FGFR3, NFE2L2, ALK, HSP90AB1, FLT3, PTEN, BRCA2, GLI1, IKBKB, CASP8, MECOM, HEY1, CASP3, JAK2, APC2, DCC, PLEKHG5, F2, WNT16, PLCB3, TRAF4, PIK3CA, TRAF5, PLCB2, LAMA2, ROCK1, LAMA1, LAMA3, XIAP, ADCY2, PTGS2, RELA, EGFR, DLL4, RXRB, NRAS, ERBB2, CTNNA1, E2F2, MAPK1, CTNNA3, BAK1, CTNNA2, TCF7L1, LAMB3, LAMB2, STAT2, STAT3, LAMB1, MTOR, APC* | 6.86E-04 |
| hsa05410: Hypertrophic cardiomyopathy | 27 | *RYR2, LAMA2, ITGB4, LAMA1, PRKAG1, ATP2A3, ATP2A2, CACNA1C, CACNA1F, SLC8A1, ACTB, TTN, DMD, CACNG3, SGCG, TPM4, TPM3, TNNC1, CACNA2D3, CACNB1, SLC8A3, ACTC1, DES, ITGA8, ITGA7, ITGA5, MYH6* | 7.14E-04 |

| | | | |
|---|---|---|---|
| hsa05414: Dilated cardiomyopathy | 28 | *RYR2, LAMA2, ITGB4, LAMA1, ATP2A3, ATP2A2, ADCY2, CACNA1C, CACNA1F, SLC8A1, ACTB, TTN, DMD, CACNG3, SGCG, TPM4, TPM3, TNNC1, CACNA2D3, CACNB1, SLC8A3, ACTC1, DES, ITGA8, GNAS, ITGA7, ITGA5, MYH6* | 7.42E-04 |
| hsa05222: Small cell lung cancer | 27 | *RB1, LAMA2, LAMA1, LAMA3, PTEN, XIAP, PIK3R1, PTGS2, RELA, IKBKB, RXRB, CASP3, AKT3, E2F2, AKT1, BAK1, SKP2, ZBTB17, LAMB3, LAMB2, LAMB1, COL4A2, TRAF4, PIK3CA, CCNE1, TRAF5, COL4A5* | 8.50E-04 |
| hsa05213: Endometrial cancer | 19 | *APC2, TCF7L1, PTEN, BRAF, PIK3R1, ELK1, EGFR, NRAS, APC, PIK3CA, CDH1, AKT3, ERBB2, CTNNA1, AKT1, MAPK1, CTNNA3, BAK1, CTNNA2* | 0.0034684 |
| hsa04218: Cellular senescence | 36 | *RB1, TRAF3IP2, SERPINE1, PTEN, ITPR2, CALML3, PIK3R1, ETS1, RELA, ZFP36L2, CCNB3, NRAS, AKT3, E2F2, AKT1, MAPK1, SMAD3, HLA-B, NFATC3, HLA-C, NFATC2, TSC1, HLA-A, HLA-F, SIRT1, CDC25A, MTOR, PIK3CA, CCNE1, MDM2, CALM3, RAD1, SLC25A31, SLC25A5, PPID, ATR* | 0.006215 |
| hsa04611: Platelet activation | 30 | *MYLK2, ROCK1, ITPR2, ADCY2, PIK3R1, ARHGAP35, ACTB, MYLK, AKT3, AKT1, MAPK1, PRKG1, PRKCI, PLA2G4F, PTGIR, PPP1R12A, VWF, NOS3, GP1BB, F2, APBB1IP, COL1A1, PLCB3, COL1A2, PIK3CA, JMJD7-PLA2G4B, TBXAS1, GNAS, PLCB2, FERMT3* | 0.0090879 |
| hsa04151: PI3K-Akt signaling pathway | 66 | *HSP90AB1, CSF1, FLT3, ITGB4, PTEN, IGF1R, GHR, GYS2, IKBKB, TCL1B, STK11, PPP2R1B, FGF9, AKT3, KDR, AKT1, TNR, JAK2, MAGI1, VWF, TSC1, YWHAZ, EREG, CREB3, COL2A1, COL4A2, PIK3CA, CCNE1, ITGA8, COL4A5, COL6A3, ITGA7, COL6A6, COL6A5, ITGA5, CSF1R, PHLPP1, LAMA2, LAMA1, LAMA3, LPAR4, PIK3R1, THBS1, EGFR, RELA, HSP90B1, NRAS, RELN, ERBB4, ERBB2, MAPK1, PCK1, LAMB3, LAMB2, NOS3, INSR, LAMB1, MTOR, GH2, COL1A1, COL1A2, LPAR5, GNB1, MDM2, COL9A3, FGFR3* | 0.0101039 |
| hsa05215: Prostate cancer | 25 | *RB1, HSP90AB1, PTEN, PIK3R1, RELA, EGFR, HSP90B1, IGF1R, IKBKB, NRAS, AKT3, ERBB2, E2F2, AKT1, MAPK1, EP300, TCF7L1, BRAF, MTOR, CREB3, ZEB1, PIK3CA, CCNE1, MDM2, ERG* | 0.0101039 |
| hsa04520: Adherens junction | 24 | *TCF7L1, SMAD3, ROCK1, CSNK2A1, ACTN1, INSR, ARHGAP29, PTPRM, IQGAP1, NLK, PTPRF, EGFR, ACTB, IGF1R, TJP1, PTPRB, CDH1, ERBB2, CTNNA1, EP300, MAPK1, CTNNA3, CTNNA2, SSX2IP* | 0.0123072 |
| hsa04015:Rap1 signaling pathway | 43 | *CSF1R, DOCK4, CSF1, CALML3, ADCY2, LPAR4, PIK3R1, SIPA1L2, ITGAL, THBS1, RAP1GAP, EGFR, ACTB, IGF1R, GRIN2A, NRAS, FGF9, CDH1, AKT3, KDR, AKT1, MAPK1, PLCG1, MAGI1, VAV3, PRKCI, PRKCB, MAGI3, INSR, BRAF, GRIN1, APBB1IP, PLCB3, ADORA2A, LPAR5, PIK3CA, ID1, GNAS, CALM3, PLCB2, FGFR3, RAPGEF6, BCAR1* | 0.0141229 |
| hsa04921:Oxytocin signaling pathway | 34 | *RYR1, RYR2, MYLK2, ROCK1, PRKAG1, ITPR2, CALML3, ADCY2, CACNA1C, CACNA1F, PTGS2, ELK1, EGFR, RYR3, ACTB, MYLK, TRPM2, NRAS, MAPK1, CACNG3, KCNJ3, PLA2G4F, PPP1R12A, PRKCB, NOS3, NFATC3, CACNA2D3, NFATC2, CACNB1, PLCB3, JMJD7-PLA2G4B, GNAS, CALM3, PLCB2* | 0.0141229 |
| hsa00562:Inositol phosphate metabolism | 20 | *ISYNA1, PTEN, OCRL, MTMR4, MTMR7, MTM1, FIG4, INPP4A, ITPKC, INPP4B, PLCB3, PIK3CA, INPP5F, IMPA1, INPP5K, PIK3C3, PLCH1, PLCH2, PLCG1, PLCB2* | 0.0151365 |

| | | | |
|---|---|---|---|
| hsa04931:Insulin resistance | 26 | *PRKAG1, PTEN, SLC2A2, PPP1R3A, IRS2, PIK3R1, ACACB, RELA, PTPRF, IKBKB, GYS2, RPS6KA6, AKT3, AKT1, PCK1, SREBF1, PRKCB, NOS3, PRKCE, INSR, STAT3, MTOR, CREB3, TBC1D4, PIK3CA, SLC27A2* | 0.0160115 |
| hsa00310:Lysine degradation | 18 | *KMT2E, KMT2D, ECHS1, KMT2A, SETDB1, KMT2C, SETD1B, SETD7, DOT1L, DLST, ASH1L, DHTKD1, MECOM, ALDH1B1, SMYD3, COLGALT1, EZH1, EZH2* | 0.0160115 |
| hsa04020:Calcium signaling pathway | 49 | *RYR1, RYR2, MYLK2, ATP2A3, ATP2A2, CALML3, SLC8A1, GRM1, RYR3, MYLK, HTR6, CYSLTR2, FGF9, KDR, LHCGR, PRKCB, TNNC1, PHKA1, TACR1, ITPKC, PLCB3, ADORA2A, AGTR1, SLC25A5, PLCB2, PDE1C, ITPR2, ADCY2, CACNA1C, CACNA1F, EGFR, CACNA1H, GRIN2A, ERBB4, ERBB2, PLCG1, NOS3, NTRK3, NFATC3, NFATC2, PHKB, GRIN1, SLC8A3, P2RX4, GRIN3A, GNAS, CALM3, SLC25A31, FGFR3* | 0.0164556 |
| hsa05205:Proteoglycans in cancer | 41 | *ROCK1, PXN, ITPR2, PIK3R1, IQGAP1, THBS1, ELK1, EGFR, ACTB, IGF1R, NRAS, ERBB4, CASP3, AKT3, ERBB2, KDR, GPC3, FLNA, AKT1, MAPK1, FLNC, PLCG1, FZD1, VAV3, PPP1R12A, FZD5, PRKCB, STAT3, ANK2, BRAF, ANK3, WNT16, HSPG2, ANK1, MTOR, COL1A1, TFAP4, COL1A2, PIK3CA, MDM2, ITGA5* | 0.0226165 |
| hsa04933:AGE-RAGE signaling pathway in diabetic complications | 24 | *SMAD3, PRKCB, NOS3, PRKCE, STAT3, SERPINE1, CYBB, PIK3R1, RELA, COL1A1, PLCB3, NRAS, COL1A2, COL4A2, PIK3CA, CASP3, AKT3, AGTR1, COL4A5, AKT1, MAPK1, PLCG1, JAK2, PLCB2* | 0.0229725 |
| hsa04910:Insulin signaling pathway | 30 | *PRKAG1, PPP1R3A, IRS2, CALML3, PIK3R1, ELK1, ACACB, HK2, PTPRF, IKBKB, GYS2, LIPE, NRAS, PRKAR2A, AKT3, AKT1, MAPK1, PCK1, SREBF1, PRKCI, INSR, PHKA1, PHKB, TSC1, BRAF, MTOR, PPP1R3F, PIK3CA, FASN, CALM3* | 0.0239456 |
| hsa05223:Non-small cell lung cancer | 19 | *RB1, ALK, PRKCB, STAT3, BRAF, PIK3R1, EGFR, RXRB, NRAS, PIK3CA, KIF5C, AKT3, ERBB2, KIF5A, E2F2, AKT1, MAPK1, BAK1, PLCG1* | 0.0239456 |
| hsa04371:Apelin signaling pathway | 30 | *RYR1, RYR2, MYLK2, SERPINE1, PRKAG1, ITPR2, CALML3, ADCY2, SLC8A1, RYR3, MYLK, LIPE, NRAS, MAP1LC3C, CDH1, AKT3, AKT1, MAPK1, SMAD3, NOS3, PRKCE, MTOR, SLC8A3, MYL4, PLCB3, GNB1, AGTR1, CALM3, PIK3C3, PLCB2* | 0.027354 |
| hsa01521:EGFR tyrosine kinase inhibitor resistance | 20 | *PRKCB, STAT3, PTEN, BRAF, NRG1, PIK3R1, EGFR, MTOR, IGF1R, NRAS, PIK3CA, AKT3, ERBB2, KDR, NF1, AKT1, MAPK1, PLCG1, JAK2, FGFR3* | 0.027354 |
| hsa04724:Glutamatergic synapse | 26 | *GRIA1, GRIK5, ITPR2, ADCY2, CACNA1C, GRM1, GRIN2A, GRM8, MAPK1, SLC17A6, DLGAP1, SLC17A8, LZTS3, KCNJ3, PLA2G4F, PRKCB, GRIN1, PLCB3, GRIN3A, DLG4, JMJD7-PLA2G4B, GNB1, GNAS, PLCB2, SHANK3, SHANK1* | 0.0280552 |
| hsa04144:Endocytosis | 47 | *SH3GL3, RAB5B, WIPF1, SH3KBP1, WIPF3, CLTB, AP2A1, ASAP2, EGFR, IGF1R, SNX2, PSD2, KIF5C, KIF5A, CLTCL1, RAB11FIP2, LDLR, ARFGEF1, SH3GLB1, HSPA8, PRKCI, SMAD3, IQSEC3, HLA-B, ARAP2, HLA-C, EPS15L1, STAM, HLA-A, HLA-F, IGF2R, DNM1, ARFGAP2, RNF41, DNM2, DAB2, ACAP2, ZFYVE16, ARPC2, NEDD4, RUFY2, CHMP2B, MDM2, VPS45, CHMP7, FGFR3, FOLR1* | 0.0303303 |
| hsa05214:Glioma | 19 | *RB1, PRKCB, PTEN, BRAF, CALML3, PIK3R1, EGFR, MTOR, IGF1R, NRAS, PIK3CA, AKT3, MDM2, E2F2, AKT1, MAPK1, CALM3, BAK1, PLCG1* | 0.0314194 |

| | | | |
|---|---|---|---|
| hsa04022:cGMP-PKG signaling pathway | 34 | *MYLK2, ROCK1, ATP2A3, ATP2A2, ITPR2, IRS2, CALML3, ADCY2, CACNA1C, CACNA1F, SLC8A1, MYLK, AKT3, CNGA1, AKT1, MAPK1, PRKG1, PPP1R12A, NOS3, PRKCE, INSR, NFATC3, NFATC2, SLC8A3, CREB3, PLCB3, KCNU1, KCNMA1, AGTR1, CALM3, SLC25A31, SLC25A5, PLCB2, MYH6* | 0.0314194 |
| hsa04934:Cushing syndrome | 32 | *RB1, KMT2D, SCARB1, KMT2A, ITPR2, ADCY2, CACNA1C, CACNA1F, CACNA1H, EGFR, MC2R, CYP11A1, DVL3, E2F2, MAPK1, PDE8A, LDLR, MEN1, FZD1, APC2, TCF7L1, FZD5, BRAF, WNT16, NR5A1, CREB3, PLCB3, APC, CCNE1, GNAS, AGTR1, PLCB2* | 0.0325254 |
| hsa05225:Hepatocellular carcinoma | 34 | *RB1, SMARCD2, PTEN, MGST1, PIK3R1, ELK1, EGFR, ACTB, IGF1R, NRAS, AKT3, DVL3, E2F2, AKT1, MAPK1, BAK1, PLCG1, ARID2, FZD1, APC2, GSTM3, PBRM1, TCF7L1, SMAD3, FZD5, PRKCB, BRAF, ARID1A, WNT16, MTOR, SMARCA4, APC, PIK3CA, NFE2L2* | 0.0325254 |
| hsa05017:Spinocerebellar ataxia | 30 | *GRIA1, RYR1, RBPJL, KCNC3, ATP2A3, ATP2A2, ITPR2, AMBRA1, PIK3R1, GRM1, ATXN3, PSMD6, GRIN2A, RELN, PSMD3, AKT3, AKT1, SPTBN2, PRKCB, PUM1, PDYN, MTOR, GRIN1, PLCB3, GRIN3A, PIK3CA, PIK3C3, SLC25A31, SLC25A5, PLCB2* | 0.0331764 |
| hsa05417:Lipid and atherosclerosis | 41 | *HSP90AB1, NCF2, CALML3, PIK3R1, RELA, HSP90B1, IKBKB, RXRB, NRAS, CASP8, CYP2B6, IRAK1, CASP6, CASP3, AKT3, AKT1, MAPK1, OLR1, NLRP3, PLCG1, JAK2, APOB, LDLR, VAV3, HSPA8, POU2F1, NOS3, STAT3, MIB2, NFATC3, CYBB, NFATC2, APOA4, POU2F3, MIB1, PLCB3, PIK3CA, IRF7, CALM3, PLCB2, NFE2L2* | 0.0344615 |
| hsa04730:Long-term depression | 16 | *GRIA1, RYR1, PLA2G4F, PRKCB, ITPR2, BRAF, GRM1, IGF1R, PLCB3, NRAS, PPP2R1B, JMJD7-PLA2G4B, GNAS, MAPK1, PLCB2, PRKG1* | 0.0375559 |
| hsa04935:Growth hormone synthesis, secretion and action | 26 | *ITPR2, IRS2, ADCY2, CACNA1C, PIK3R1, CACNA1F, GHR, NRAS, AKT3, AKT1, MAPK1, EP300, PLCG1, JAK2, JUNB, POU1F1, PRKCB, STAT3, MTOR, GH2, CREB3, PLCB3, PIK3CA, GNAS, PLCB2, BCAR1* | 0.0382362 |
| hsa05221:Acute myeloid leukemia | 17 | *CSF1R, TCF7L1, JUP, FLT3, STAT3, BRAF, PIK3R1, MPO, MTOR, RELA, RUNX1, IKBKB, NRAS, PIK3CA, AKT3, AKT1, MAPK1* | 0.0433785 |
| hsa05224:Breast cancer | 30 | *RB1, NOTCH1, NOTCH4, PTEN, PIK3R1, BRCA2, EGFR, IGF1R, DLL4, NRAS, FGF9, HEY1, AKT3, ERBB2, DVL3, E2F2, AKT1, MAPK1, BAK1, FZD1, APC2, TCF7L1, FZD5, BRAF, WNT16, MTOR, NFKB2, APC, PIK3CA, PGR* | 0.0433785 |
| hsa02010:ABC transporters | 13 | *ABCC3, ABCA2, ABCC2, ABCC8, ABCB7, ABCB5, ABCA4, TAP2, ABCC9, ABCA8, ABCB11, ABCA13, ABCG2* | 0.045152 |

**Supplementary Table S8: Significant GO-BP terms associated with the mutated genes having functional impact on the protein function**

| GO-BP Terms | Gene Count | Genes | FDR |
|---|---|---|---|
| GO:0007155~cell adhesion | 115 | *COL16A1, COL12A1, CDH8, PNN, TNR, SVEP1, HABP2, MAGI1, DSCAM, ACTN2, PRKCE, OMD, CASK, ADAM2, FLG2, ADAM17, BCAM, OBSCN, CLDN8, SPECC1L, PKP1, DSC2, FREM3, NTM, PXN, PCDH15, PCDH19, KLC1, NID2, PCDH17, EFNB1, PDZD2, PCDHA2, PCDHA7, JUP, SIGLEC12, CADM2, PCDH20, PTPRD, COL1A1, PCDHGA11, FAT1, CNTN1, CNTN4, PCDHB3, PTPRT, CNTNAP2, SRPX, PCDHGB7, ITGB4, TGFB1I1, STAB2, PTPRM, ATP2A2, ITGAL, ARHGAP5, HAPLN3, PTPRF, ROBO1, ADAMTSL1, CHL1, CLDN25, EMILIN2, EDIL3, NEO1, PCDHGA8, VWF, PCDHGA4, GP1BB, CLDN11, VCAN, MMRN1, MMRN2, ITGA8, COL6A3, ITGA7, COL6A6, CDHR5, COL6A5, ITGA5, DSCAML1, SSX2IP, CNTNAP5, CNTNAP4, NLGN3, LAMA2, LAMA1, LAMA3, THBS1, TM9SF4, ACAN, RELN, CTNNA1, CTNNA3, CTNNA2, OPCML, JAM3, NLGN4X, LAMB3, PCDHGC5, LAMB2, PCDHGC3, PCDH8, PCDHB14, PCDHB13, LAMB1, PCDHB12, NFASC, NPHS1, APC, COL7A1, AGGF1, SIGLEC8, FOLR1, BCAR1* | 2.02E-08 |
| GO:0045944~positive regulation of transcription from RNA polymerase II promoter | 196 | *RB1, FOXI1, ELK1, SOX3, SOX1, ZNF729, SOX18, ZNF606, KAT8, AKT1, HOXA1, JUNB, TP63, MED1, SLC11A1, RUNX1, HCFC1, RFX6, MAML3, CLOCK, AIRE, EPAS1, GATA6, PIK3R1, GATA2, PRDM10, ZNF827, DHX36, MYO6, HLTF, HIVEP1, TP53BP1, ZNF148, ZNF268, ZNF267, ZNF384, JUP, FZD5, BCL11B, NFATC3, NFATC2, NR1D2, NR1D1, SMARCA1, ST18, SMARCA4, SETX, ZFAT, ZNF814, REL, CDK13, EZH1, TP73, SETD4, TOP2A, GLI1, HOXC11, PHF8, MED12, IKBKB, MED13, MECOM, HEY1, HNF4A, JAK2, TEAD2, TEAD4, SREBF1, ANXA2, NCOA6, TET1, PAX3, EN1, AP3B1, ASH1L, SIRT1, NR5A1, TOX3, ZEB2, ZEB1, ZNF780B, MPND, ZNF594, TMPRSS6, SATB2, DOT1L, GPS2, ASCL2, RELA, EGFR, RXRB, GBX2, ZNF229, TBL1X, ZNF224, BRD4, IL33, STAT2, MCRS1, STAT3, WWP2, POU4F2, GRIN1, SKI, ZNF33A, TFAP4, NFIB, TCEA2, PRDM4, NRL, ARID4B, UBE3A, ETS1, SALL2, STK16, EP300, MEN1, MLIP, CASK, POU3F2, RNF40, ZNF91, MAF, KAT6A, PGR, PHIP, ERG, MZF1, KMT2D, DLX2, NOTCH1, KMT2A, TNKS, KMT2C, RLF, NEUROD2, DVL3, NLRP3, SMYD3, OTX1, NKX6-1, ZBTB17, AKIRIN1, ZFHX3, POU2F1, SMAD3, AUTS2, POU2F3, NFKB2, DAB2, BMP2, LHX2, ZNF658, PIN1, BMPR1A, NFE2L2, ZNF770, DDX3X, CHD8, CHD6, PSIP1, NR2E1, ZIC3, NFKBIZ, ZIC1, NCK2, POU1F1, PPP1R12A, PROP1, ATRX, FOXL2, CREB3, THRAP3, ELF4, HAND2, IRF7, ITGA8, ZNF750, RBPJL, CEBPB, HLF, NLRC5, HDAC8, ATAD2B, IFI16, E2F2, E2F7, ATOH1, EYA1, IRX6, FOXJ1, TBX5, ASXL1, KLF6, ASXL3, TAF7* | 2.91E-06 |
| GO:0007156~homophilic cell adhesion via plasma membrane adhesion molecules | 45 | *PTPRT, ROBO3, PCDHGB7, PCDH15, PTPRM, CELSR1, PCDH19, CELSR3, CDH8, PCDH17, MYPN, ROBO1, CDH7, CDH1, PCDHA2, CDH26, HMCN1, PCDHA7, PCDHGA8, DSCAM, PCDHGA4, PCDHGC5, CADM2, PCDH20, PCDHGC3, PCDH8, VSTM2L, PCDHB14, PCDHB13, PCDHB12, IGFN1, PCDHGA11, SDK1, DCHS2, FAT1, FAT2, DSG1, DCHS1, CDHR5, FAT3, PCDHB3, CNTN4, DSCAML1, DSC2, CDH18* | 4.53E-05 |

| | | | |
|---|---|---|---|
| GO:0006325~chromatin organization | 61 | *RB1, KDM5C, CHD9, PHF1, CHD8, SETD7, CHD6, JMJD1C, IKZF1, CHD3, PHF8, SOX1, TLK2, KAT8, EP300, HMG20A, JAK2, MEN1, BRD1, KDM2B, PRKCB, ATRX, TET1, ASH1L, SAFB, SIRT1, RNF40, RNF168, DNAJC2, DPPA3, WAC, NUP98, BCORL1, KMT2E, KMT2D, KMT2A, KMT2C, DOT1L, HDAC8, HDAC9, RELA, HLTF, EP400, BRD4, TCF7L1, KDM4B, SETDB1, EYA1, EYA2, SETD1B, LRWD1, ZZEF1, SMARCA4, PARP10, ASXL1, KANSL1, BCOR, EZH1, BAP1, FAM50A, EZH2* | 1.46E-04 |
| GO:0030534~adult behavior | 16 | *NLGN3, ALK, CNTNAP2, ZFHX2, NLGN4X, PTEN, NRXN3, NRXN2, PCDH17, ADAM2, NAGLU, SHANK3, BBS4, SHANK1, ATP6V1B1, SPTBN2* | 2.33E-04 |
| GO:0007018~microtubule-based movement | 28 | *DNAH1, DNAH2, DNAH7, DNAH8, DNAH5, DNAH6, DNAH9, KLC1, KIF17, KIF15, KIF5C, KIF5A, KIF13B, KIF21A, KIF21B, KIF1A, DNAI1, DYNC2H1, DYNC1H1, DNAH17, DNAH14, TCTE1, KTN1, CENPE, KIFC1, KIF4A, KIF26B, KIF20B* | 2.33E-04 |
| GO:0098609~cell-cell adhesion | 46 | *VEZT, TENM2, COL13A1, LAMA3, PRICKLE1, CELSR1, ITGAL, LPP, CELSR3, EGFR, CDH8, CDH7, CNN3, BAIAP2L1, CDH1, CTNNA1, NPHP4, CTNNA3, CTNNA2, NEO1, JAM3, DSP, CD93, JUP, ANXA2, CNTN5, DCC, TJP1, NFASC, OBSCN, DLG2, NPHS1, DCHS2, DLG4, LRRC7, ITGA8, FAT1, CNTN1, PKP1, FAT2, ITGA7, DSG1, ELMO2, FAT3, ITGA5, DSC2* | 2.43E-04 |
| GO:0007411~axon guidance | 45 | *CSF1R, ROBO3, NRP1, TENM2, TRIO, NOTCH1, LAMA2, EPHA10, SEMA3D, LAMA1, LAMA3, MYCBP2, PTPRO, NRXN3, SEMA3E, KALRN, ARHGAP35, MYPN, ADAMTSL1, EFNB1, RELN, GBX2, CHL1, KIF5C, PLEKHG4B, KIF5A, TNR, ARHGEF40, NEO1, ATOH1, PLXNA3, SEMA6B, EPHA5, DSCAM, CNTN5, LAMB2, DCC, SEMA4C, VSTM2L, POU4F2, NFASC, LHX2, CNTN1, CNTN4, DSCAML1* | 2.59E-04 |
| GO:0086010~membrane depolarization during action potential | 11 | *KCNH2, SCN10A, SCN11A, SCN8A, SCN9A, SCN4A, SCN5A, SCN2A, SCN3B, SCN3A, CACNA1H* | 4.68E-04 |
| GO:0019228~neuronal action potential | 16 | *KCND2, KCNA1, KCNA2, ANK3, MTOR, CACNA1H, SCN10A, P2RX4, SCN11A, SCN8A, SCN9A, MYH14, SCN4A, SCN5A, SCN2A, SCN3A* | 5.00E-04 |
| GO:0000122~negative regulation of transcription from RNA polymerase II promoter | 152 | *RB1, ZNF177, MYT1L, MAGEB16, WWC1, ATN1, ARID4B, PRDM1, FOXQ1, SOX3, SOX1, SALL2, FGF9, SOX18, WWC3, EP300, ZNF568, MAGEA6, TP63, MEN1, MED1, ZNF560, MLIP, MORC3, KDM2B, ESX1, HIC1, RUNX1, FOXP1, HCFC1, ZNF91, MAF, ZNF93, THAP7, CCNE1, MAGEB2, TXNIP, BCORL1, MZF1, PPID, MAGEB5, ZNF431, ZBTB8A, DLX2, MAGEA12, NOTCH1, MAGED1, NOTCH4, GATA6, GATA2, PCBP3, TPR, HIVEP1, MAGEC1, ZNF148, NKX6-1, ZNF268, ZBTB17, PLK3, ZFHX3, SMAD3, MAGEE1, NFATC3, NFATC2, NR1D2, NR1D1, SCRT1, SMARCA4, DAB2, BMP2, CNOT1, ID1, MDM2, NFE2L3, SIM2, EZH1, NFE2L1, CC2D1B, EZH2, KDM5A, ZNF512B, TENM2, BTG2, ZNF491, CHD8, CHD3, NR2E1, SDR16C5, SIN3B, HEY1, DNMT3B, HMG20A, JARID2, LMCD1, ZNF587B, DACT1, SREBF1, POU1F1, PROP1, RFC1, TET1, EN1, FOXL2, YWHAZ, SIRT1, ETV7, NCOR2, ZEB2, ZEB1, CRY2, IRF7, STRAP, MCPH1, ZNF750, CEBPB, TMPRSS6, SATB2, GPS2, HDAC8, CXXC5, HDAC9, ASCL2, RELA, DLL4, CUX2, IFI16, TBX22, TBL1X, ZNF224, ZIM2, E2F7, SPEN, IL33, SKOR1, USP9X, STAT3, ZNF180, FOXJ1, WWP2, KLF3, POU4F2, TBX18, SKI, PER3, NFIB, HNRNPA2B1, ZNF217, TAF7, TRIM37, BCOR, TRIM33, TAF1* | 6.78E-04 |

| | | | |
|---|---|---|---|
| GO:0007275~multicellular organism development | 36 | *ALK, CSF1R, HLF, FLT3, GREB1L, FGD1, EGFR, CDH8, IGF1R, CDH7, ZMYM3, ERBB4, CDH1, ZMYM4, ERBB2, KDR, HOXA1, CDH26, ADAMTS9, ZNF267, TIE1, NTRK3, INSR, GREB1, PCOLCE, MERTK, KLF3, MMP11, VCAN, BMP1, ROR2, CDK13, FGFR3, CHST2, CDH18, ATR* | 8.11E-04 |
| GO:0016477~cell migration | 56 | *SH3KBP1, ITGB4, WWC1, ATN1, PTEN, FMN2, PTPRF, ARHGAP35, SYDE2, WWC3, KDR, NCK2, NDEL1, APC2, VAV3, PRKCI, PROP1, SHROOM2, GFRA1, RHOC, ATRN, PIK3CA, ELMO2, DOCK2, PHLDB2, ENPEP, BRAT1, PXN, LAMA3, IQGAP1, THBS1, CLN3, ERBB4, GPC3, CTNNA1, CTNNA3, PLCG1, CTNNA2, GPC6, JAM3, NTNG1, JUP, LAMB3, USP9X, LAMB2, NFATC2, LAMB1, CDC42BPB, MERTK, SORL1, CCDC88A, APC, TAOK2, FAT1, PLXNB1, BCAR1* | 9.28E-04 |
| GO:0034765~regulation of ion transmembrane transport | 34 | *CATSPER1, CATSPER4, KCNA1, KCNC3, KCNA2, CACNA1C, CACNA1F, CACNA1H, KCNV2, SCN11A, SCN9A, SCN5A, SCN3B, SCN3A, KCNJ3, KCNH2, KCNF1, KCND1, KCND2, CACNA2D3, KCNJ15, CYBB, KCNJ16, KCNK16, WWP2, CACNB1, SCN10A, KCNU1, NEDD4, SCN8A, KCNMA1, KCNQ2, SCN4A, SCN2A* | 0.001 |
| GO:0006468~protein phosphorylation | 92 | *SMG1, TRIO, MAST2, PRKAG1, AATK, MYLK, IKBKB, LIPE, STK11, RPS6KA6, PPP4R1, AKT3, TLK2, AKT1, PIM3, JAK2, MAP3K6, PSKH2, PRKG1, EPHA5, MAP4K2, CDKL4, PRKCI, MORC3, CDKL5, CSNK2A1, PRKCB, DAPK1, PRKCE, CASK, KSR2, YWHAZ, NRBP1, CIT, LATS1, FGR, OBSCN, CCNE1, BMP2K, PIK3C3, ALPK3, TNIK, ERG, ROR2, ALPK1, ROCK1, EPHA10, LAMA1, WNK4, PIK3R1, NLK, KALRN, TTBK1, CAMKK1, TTN, CLN3, DGUOK, IRAK1, ERBB2, MAPK1, CSK, MARK4, TRPM6, PAK2, NEK8, SLC12A3, PLK3, DMPK, LIMK2, TIE1, NTRK3, INSR, PHKB, BRAF, CDC42BPB, MERTK, MTOR, CLK3, SRPK1, BMP2, MAP3K15, TAOK3, WNK1, PRKRA, TAOK1, TAOK2, WNK3, TAF7, CDK13, BMPR1A, ATR, TAF1* | 0.00104 |
| GO:0007420~brain development | 53 | *CNTNAP2, IFT172, CHD8, ROGDI, AATK, UBE3A, IRS2, BRCA2, UBE2L3, PHF8, RGS4, SIX6, ZIC2, FPGS, ZIC1, PPFIA1, CCIN, MACROD2, CDK5RAP2, MEN1, MED1, CCDC39, NCOA6, SHROOM2, HSPG2, EML1, ITGA8, DSCAML1, DLX2, RTN1, PCDH19, GRIN2A, RELN, OXCT1, DHX37, CSK, BAK1, SLC17A8, ZFHX3, KDM4B, ZFHX2, CNTN5, XRCC5, CADM2, FOXJ1, SMARCA1, GRIN1, NFASC, NFIB, ID1, NF1, CNTN1, CNTN4* | 0.00159 |
| GO:0006338~chromatin remodeling | 63 | *RB1, KDM5A, SETD4, SMARCAL1, ERCC6L, KDM5C, CHD9, CHD8, CHD6, PSIP1, JMJD1C, CHD3, BRCA2, CHD1, ACTB, PHF8, NFKBIZ, KAT8, JAK2, JARID2, ARID2, TP63, TGM2, PBRM1, BRD1, KDM2B, PRKCB, ATRX, TET1, ASH1L, ARID1A, HCFC1, SLFN11, WAC, MPND, CLOCK, INO80C, SMARCD2, INO80E, PAXIP1, SRCAP, KMT2C, SATB2, BAZ2B, HDAC8, CHRAC1, ZNF827, HLTF, EP400, BRD4, KDM4B, EYA1, EYA2, SETD1B, MCRS1, HUWE1, SMARCA1, SMARCA4, BCOR, EZH1, BAP1, BRDT, TAF1* | 0.00175 |
| GO:0045893~positive regulation of transcription, DNA-templated | 115 | *WWC1, ATN1, ETS1, ELK1, ACTB, UBE2L3, SOX18, CDH1, CAPN3, KAT8, AKT1, EP300, SUPT20H, TP63, RNF111, CDK5RAP2, KCNH2, MED1, ACTN2, PRKCB, ACTN1, POU3F1, RUNX1, HCFC1, RFX6, KAT6A, PHIP, ROR2, NUP98, CLOCK, INO80C, KMT2E, BLM, INO80E, NOTCH1, SRCAP, KMT2A, MAGED1, AIRE, NOTCH4, GATA6, RLF, RAI1, EP400, INPP5K, DVL3, HIVEP3, TP53BP1, ZNF423, FZD1, ZFHX3, SMAD3, INSR, NFATC3, NFATC2, NR1D2, NR1D1, SMARCA1, SMARCA4, COL1A1, BMP2, LHX2, HLA-DRB1, TP73, KDM5A, ZNF496, CHD8, TGFB1I1, CHD3, BRCA2, GLI1, PHF8, IKBKB, MED12, MED13, MECOM, ZIC2, ZIC3, HNF4A, ZIC1, CEP290, TEAD2, RFC1, NCOA6, PAX3, FOXL2, ARID1A, DNM2, DNAJC2, CREB3, ELF4, THRAP3, IRF7, WAC, CEBPB, SMARCD2,* | 0.00211 |

| | | | |
|---|---|---|---|
| | | *RELA, EGFR, RXRB, TBX22, ERBB4, TBL1X, BRD4, IRX6, MCRS1, MGA, STAT3, TBX5, TBX18, KLF6, TFAP4, KANSL1, NFIB, TAF7, TRIM37* | |
| GO:0060285~cilium-dependent cell motility | 10 | *CCDC39, DNAH1, DNAH2, DNAH17, DNAH7, DNAH8, DNAH14, DNAH6, EFHC1, EFHC2* | 0.00276 |
| GO:0007416~synapse assembly | 22 | *GRIA1, NLGN3, GPM6A, FARP1, DSCAM, FZD5, PLXND1, PTEN, NRXN3, NRXN2, PCDHB14, NRG1, PCDHB13, IGFN1, SDK1, PCLO, CDH1, ERBB4, PCDHB3, BSN, SHANK3, SPTBN2* | 0.00374 |
| GO:0046777~protein autophosphorylation | 39 | *ALK, CSF1R, MYLK2, SMG1, FLT3, TTK, NLK, EGFR, IGF1R, STK11, IRAK1, ERBB4, STK16, ERBB2, KDR, AKT1, PIM3, CSK, JAK2, PAK2, MAP3K3, CDKL5, DAPK1, INSR, MTOR, CLK3, UHMK1, FGR, PTPRC, TAOK3, TAOK1, TAOK2, MYO3A, WNK3, CALM3, TNIK, FGFR3, TAF1, ATR* | 0.00522 |
| GO:0007417~central nervous system development | 34 | *SH3GL3, TRIO, ATN1, PTEN, CAPG, HAPLN3, CELSR1, KALRN, DRP2, SOX3, UGT8, ACAN, RPS6KA6, RELN, PTPRZ1, ZIC2, ZIC3, ZIC1, JARID2, ATOH1, SEMA6B, ARSA, LHCGR, SZT2, PROP1, EIF2B2, LIG4, MARCKS, VCAN, COL2A1, ADORA2A, IL1RAPL2, NMUR2, DSCAML1* | 0.00606 |
| GO:0030036~actin cytoskeleton organization | 40 | *SPTBN4, ROCK1, EPB41, FMN2, FGD1, AMOT, SPTA1, CLN3, ATXN3, FGD6, POF1B, FLNA, INPP5K, DMD, NPHP4, CCIN, FLNC, HMCN1, SPTAN1, PRKG1, SPTBN2, PHACTR4, STARD13, ACTN2, LIMK2, ACTN1, IQSEC3, PDE4DIP, FOXJ1, CDC42BPB, TJP1, TBCK, PIK3CA, DAAM2, TAOK2, SPECC1L, NF1, DOCK2, UTRN, SMTNL2* | 0.00736 |
| GO:0043524~negative regulation of neuron apoptotic process | 34 | *NRP1, BTG2, CEBPB, HSP90AB1, ROCK1, PLXND1, GBE1, SEMA3E, AMBRA1, SLC30A10, CLU, IL27RA, FOXQ1, CLN3, PTPRZ1, CHL1, KDR, JAK2, PRKCI, KDM2B, PCDHGC5, PCDHGC3, LIG4, EN1, VSTM2L, BRAF, OXR1, SIRT1, TOX3, GCLC, ADORA2A, MSH2, PIK3CA, RASA1* | 0.01294 |
| GO:0048812~neuron projection morphogenesis | 19 | *GPM6A, CNTNAP2, TRIO, NYAP1, ANKRD27, MYO16, DNM2, UGT8, ADORA2A, SLC9A6, TAOK3, TAOK1, TAOK2, EPB41L3, PLXNB1, DMD, TNIK, KIF20B, GAS7* | 0.01353 |
| GO:0007052~mitotic spindle organization | 18 | *PLK3, DYNC1H1, NUDC, TNKS, TTK, WDR62, TUBG1, CKAP5, EML1, CENPE, CENPH, KIF4A, SBDS, EFHC1, EFHC2, TACC2, PCNT, CLASP2* | 0.01456 |
| GO:0035176~social behavior | 17 | *NLGN3, CNTNAP2, NLGN4X, GRID1, GAD1, CHD8, PTEN, NRXN3, NRXN2, EN1, NR2E1, TH, PTCHD1, DLG4, SHANK3, BBS4, SHANK1* | 0.01542 |
| GO:0098703~calcium ion import across plasma membrane | 14 | *CACNA1C, CACNA1F, CACNA1H, SLC8A1, SLC8A3, TRPM2, SCN10A, SCN11A, SCN8A, SCN9A, SCN4A, SCN5A, SCN2A, SCN3A* | 0.01682 |
| GO:0071625~vocalization behavior | 9 | *NLGN3, CNTNAP2, NLGN4X, DLG4, NRXN3, NRXN2, MYH14, SHANK3, SHANK1* | 0.01739 |

| | | | |
|---|---|---|---|
| GO:0051056~regulation of small GTPase mediated signal transduction | 28 | *ARHGAP11A, DOCK3, TRIO, ARHGAP5, SIPA1L2, KALRN, FGD1, RAP1GAP, ARHGAP35, AMOT, SYDE2, ARHGAP21, ARHGAP20, PLEKHG4B, TAGAP, ARHGEF40, VAV3, STARD13, RALGAPA2, PLEKHG5, ARHGEF19, ARAP2, ARHGAP29, ARHGAP28, OBSCN, ARHGAP30, RALGAPB, DOCK2* | 0.01739 |
| GO:0050808~synapse organization | 18 | *NLGN3, NLGN4X, PCDHGC5, PCDHGC3, LRP4, RAB39B, TSC1, SEMA3E, SLC8A3, DRP2, NFASC, SLC9A6, PPFIA1, TNR, FLNA, PIN1, PPFIA4, PPFIA2* | 0.01952 |
| GO:0006898~receptor-mediated endocytosis | 21 | *NLGN3, CUBN, LRP1, INSR, STAB2, AP2A1, RABEPK, HSPG2, SORL1, EGFR, LRP1B, IGF2R, DNM1, DNM2, CLN3, DAB2, CLTCL1, PLA2R1, FOLR1, LDLR, REPS1* | 0.02241 |
| GO:0007165~signal transduction | 176 | *SH3GL3, OLFML2B, CD83, IL5RA, IRS2, ARR3, ARHGAP35, IGF1R, SYDE2, STK11, RPS6KA6, PPP4R1, RASSF2, OR1S1, GRB14, FGF9, GRAP2, AKT3, PPFIA1, AKT1, MAP3K6, PDE8A, CHGB, MAGI1, UTS2R, CHRNB1, CSNK2A1, DAPK1, PRKCB, MAGI3, PRKCE, OR1J4, PSG1, ANK2, OCRL, ANK3, GABRG3, ANK1, GABRG1, INPP4A, SIGIRR, INPP4B, OLFM1, BCAM, ZFYVE16, PSG6, AKAP9, PKP1, PGR, ERG, ROR2, CLOCK, GRIA1, CSF1R, LRPAP1, ENPEP, PDE1C, EPAS1, CABP4, PXN, WNK4, ITPR2, IQGAP1, PIK3R1, IQGAP2, RASAL3, TRHDE, KALRN, RAP1GAP, TTBK1, ARHGAP12, C3, ARHGAP21, ARHGAP20, TP53BP2, NLRP3, NPHP4, STARD13, DTNA, STAC, TNFSF15, RASSF8, CDC7, BRAF, ARHGAP28, STAM, IGF2R, NFKB2, ARHGAP30, TG, BMP1, WNK1, DLG4, GNB1, NF1, HLA-DRB1, ARHGAP11A, PTPRT, IL21, OR7A10, ALK, NRP1, TENM1, TENM2, GDI2, PRKAG1, PTPRM, DUSP15, ITGAL, ARHGAP5, IMPA1, CHL1, TAGAP, KIF13B, PDE4B, NCK2, JAK2, PRKG1, PPP1R12A, TNFRSF18, ARAP2, GFRA1, GAB3, KSR2, RHOC, YWHAZ, DNM2, APBB1IP, RASA1, IL1RAPL2, TRAF5, RASA2, OR3A1, RBPJL, VAC14, PLEKHH3, PHLPP1, OR1E2, ROCK1, LRRC18, NRXN3, NRXN2, TPTE, EGFR, FGD1, DLL4, ARFRP1, NRAS, CHRNG, ERBB4, ERBB2, LRRC8C, LRRC8D, MAPK1, CHRNA10, PCNT, PAK2, CAP2, GABRQ, SLC12A3, GABRA6, STAT2, TIE1, STAT3, MX1, LILRB2, TSPEAR, CDC42BPB, MKLN1, NLRP12, OPN1SW, PDE10A, P2RX4, PDE7B, SIGLEC8, BCAR1* | 0.02799 |
| GO:0007010~cytoskeleton organization | 33 | *SH3KBP1, MAST2, FGD1, LMNB2, CAMSAP1, CNN2, UGT8, ARHGAP21, FGD6, PCLO, ZMYM3, ZMYM4, EPB41L3, DMD, CTNNA2, CAP2, SVIL, PRKCI, KRT4, MICAL3, SORBS2, ANK2, ANK3, CDC42BPB, ANK1, MTOR, DES, ELMO2, PHIP, TNIK, BRWD1, BRWD3, HLA-DRB1* | 0.02799 |
| GO:0007605~sensory perception of sound | 33 | *SPTBN4, PGAP1, USP53, COL11A1, HEXA, PCDH15, USH1C, OTOA, CASP3, MYO6, OTOG, OTOF, MYH14, HOXA1, SLC17A8, TBL1X, CCDC50, EYA1, CNTN5, PAX3, TSPEAR, NAV2, POU4F2, SERPINB6, TJP1, COL1A1, COL2A1, TH, MYO3B, MYO3A, PKHD1L1, ATP6V1B1, SLC26A5* | 0.04415 |
| GO:0033674~positive regulation of kinase activity | 19 | *ALK, EPHA5, VAC14, CSF1R, EPHA10, FLT3, TIE1, NTRK3, INSR, MERTK, EGFR, ZNF16, IGF1R, ERBB4, ERBB2, KDR, ROR2, FGFR3, HLA-DRB1* | 0.04979 |

**Supplementary Table S9: Non-synonymous protein coding somatic mutations identified in literature-curated driver genes in HPV16-positive CaCx patients of Indian cohort**

| Gene | Patient ID | Chromosome | Start position | End position | Reference allele | Variant allele | Variant type | cDNA change | Protein change |
|---|---|---|---|---|---|---|---|---|---|
| *PIK3CA* | 17- 397 | 3 | 178936091 | 178936091 | G | A | Missense Mutation | c.1633G>A | p.E545K |
| | 17- 397 | 3 | 178936103 | 178936103 | G | T | Missense Mutation | c.1645G>T | p.D549Y |
| | 17- 491 | 3 | 178936091 | 178936091 | G | A | Missense Mutation | c.1633G>A | p.E545K |
| | 17- 499 | 3 | 178916890 | 178916890 | C | T | Missense Mutation | c.277C>T | p.R93W |
| | 17- 499 | 3 | 178936082 | 178936082 | G | A | Missense Mutation | c.1624G>A | p.E542K |
| | 17- 580 | 3 | 178936082 | 178936082 | G | A | Missense Mutation | c.1624G>A | p.E542K |
| | 17- 590 | 3 | 178936092 | 178936092 | A | G | Missense Mutation | c.1634A>G | p.E545G |
| | 17- 599 | 3 | 178952085 | 178952085 | A | G | Missense Mutation | c.3140A>G | p.H1047R |
| | 17- 633 | 3 | 178928079 | 178928079 | G | A | Missense Mutation | c.1357G>A | p.E453K |
| | 17- 633 | 3 | 178936082 | 178936082 | G | A | Missense Mutation | c.1624G>A | p.E542K |
| | 17- 673 | 3 | 178936082 | 178936082 | G | A | Missense Mutation | c.1624G>A | p.E542K |
| | 18- 335 | 3 | 178936082 | 178936082 | G | A | Missense Mutation | c.1624G>A | p.E542K |
| | 18- 815 | 3 | 178936091 | 178936091 | G | A | Missense Mutation | c.1633G>A | p.E545K |
| | 18- 1102 | 3 | 178936082 | 178936082 | G | A | Missense Mutation | c.1624G>A | p.E542K |
| | 18- 1201 | 3 | 178936091 | 178936091 | G | A | Missense Mutation | c.1633G>A | p.E545K |
| | 18- 1201 | 3 | 178938934 | 178938934 | G | A | Missense Mutation | c.2176G>A | p.E726K |
| | 18- 1291 | 3 | 178936091 | 178936091 | G | A | Missense Mutation | c.1633G>A | p.E545K |
| | 18- 1861 | 3 | 178936082 | 178936082 | G | A | Missense Mutation | c.1624G>A | p.E542K |
| | 18- 2081 | 3 | 178936091 | 178936091 | G | A | Missense Mutation | c.1633G>A | p.E545K |
| | 18- 2158 | 3 | 178936082 | 178936082 | G | A | Missense Mutation | c.1624G>A | p.E542K |
| | 18- 2178 | 3 | 178936092 | 178936092 | A | G | Missense Mutation | c.1634A>G | p.E545G |

| | | | | | | | | | |
|---|---|---|---|---|---|---|---|---|---|
| | 18- 2302 | 3 | 178951957 | 178951957 | G | A | Missense Mutation | c.3012G>A | p.M1004I |
| | 19- 078 | 3 | 178936091 | 178936091 | G | A | Missense Mutation | c.1633G>A | p.E545K |
| | 19- 078 | 3 | 178938934 | 178938934 | G | A | Missense Mutation | c.2176G>A | p.E726K |
| | 19- 253 | 3 | 178951994 | 178951994 | G | C | Missense Mutation | c.3049G>C | p.D1017H |
| | 19- 780 | 3 | 178936091 | 178936091 | G | A | Missense Mutation | c.1633G>A | p.E545K |
| *KMT2C* | 17- 369 | 7 | 151873887 | 151873887 | C | T | Missense Mutation | c.8651G>A | p.R2884Q |
| | 17- 633 | 7 | 151882642 | 151882642 | C | A | Splice Site | NA | NA |
| | 17- 633 | 7 | 152027750 | 152027750 | G | A | Nonsense Mutation | c.325C>T | p.Q109* |
| | 18- 1062 | 7 | 151860728 | 151860728 | G | A | Nonsense Mutation | c.9934C>T | p.Q3312* |
| | 18- 1062 | 7 | 151878464 | 151878464 | G | A | Nonsense Mutation | c.6481C>T | p.Q2161* |
| | 18- 1062 | 7 | 151878659 | 151878659 | G | A | Nonsense Mutation | c.6286C>T | p.Q2096* |
| | 18- 1062 | 7 | 151879312 | 151879312 | G | A | Missense Mutation | c.5633C>T | p.S1878L |
| | 18- 2158 | 7 | 152009018 | 152009018 | G | A | Nonsense Mutation | c.604C>T | p.Q202* |
| | 19- 078 | 7 | 151878680 | 151878680 | G | A | Nonsense Mutation | c.6265C>T | p.Q2089* |
| | 19- 253 | 7 | 152012245 | 152012245 | G | A | Nonsense Mutation | c.568C>T | p.R190* |
| | 19- 974 | 7 | 151859678 | 151859678 | C | A | Nonsense Mutation | c.10984G>T | p.E3662* |
| *KMT2D* | 17- 397 | 12 | 49438211 | 49438211 | T | - | Frame Shift Deletion | c.5058delA | p.K1686fs |
| | 17- 415 | 12 | 49442892 | 49442893 | AG | - | Frame Shift Deletion | c.4015_4016delCT | p.L1339fs |
| | 17- 633 | 12 | 49425412 | 49425412 | G | C | Nonsense Mutation | c.13076C>G | p.S4359* |
| | 17- 633 | 12 | 49441780 | 49441780 | G | A | Nonsense Mutation | c.4204C>T | p.Q1402* |
| | 17- 782 | 12 | 49440474 | 49440474 | C | T | Missense Mutation | c.4336G>A | p.D1446N |
| | 19- 042 | 12 | 49422946 | 49422946 | C | A | Nonsense Mutation | c.14149G>T | p.E4717* |
| | 19- 253 | 12 | 49438693 | 49438693 | C | - | Frame Shift Deletion | c.4797delG | p.L1600fs |
| | 17- 590 | 22 | 41573318 | 41573318 | C | A | Missense Mutation | c.5603C>A | p.T1868K |
| | 17- 633 | 22 | 41560094 | 41560094 | C | G | Missense Mutation | c.3766C>G | p.Q1256E |

| | | | | | | | | | |
|---|---|---|---|---|---|---|---|---|---|
| *EP300* | 17- 633 | 22 | 41564473 | 41564473 | G | A | Missense Mutation | c.3895G>A | p.G1299S |
| | 17- 633 | 22 | 41564792 | 41564792 | G | T | Nonsense Mutation | c.4093G>T | p.E1365* |
| | 17- 633 | 22 | 41568503 | 41568503 | G | A | Splice Site | c.4453G>A | p.D1485N |
| | 17- 633 | 22 | 41568527 | 41568527 | G | T | Missense Mutation | c.4477G>T | p.D1493Y |
| | 17- 673 | 22 | 41565529 | 41565529 | G | A | Missense Mutation | c.4195G>A | p.D1399N |
| | 17- 782 | 22 | 41568505 | 41568505 | T | A | Missense Mutation | c.4455T>A | p.D1485E |
| | 18- 2158 | 22 | 41565529 | 41565529 | G | A | Missense Mutation | c.4195G>A | p.D1399N |
| *MACF1* | 17- 633 | 1 | 39851451 | 39851451 | G | A | Missense Mutation | c.14209G>A | p.D4737N |
| | 18- 336 | 1 | 39785406 | 39785406 | C | G | Missense Mutation | c.4031C>G | p.S1344C |
| | 18- 1062 | 1 | 39801107 | 39801108 | AT | - | Frame Shift Deletion | c.8862_8863delAT | p.Y2955fs |
| | 18- 2081 | 1 | 39835719 | 39835719 | C | A | Missense Mutation | c.12971C>A | p.S4324Y |
| | 19- 253 | 1 | 39853429 | 39853429 | C | G | Missense Mutation | c.14930C>G | p.S4977C |
| *HUWE1* | 17- 607 | X | 53564560 | 53564560 | G | A | Missense Mutation | c.12094C>T | p.R4032C |
| | 17- 633 | X | 53576272 | 53576272 | G | C | Missense Mutation | c.9683C>G | p.S3228C |
| | 17- 819 | X | 53621513 | 53621513 | T | C | Missense Mutation | c.3449A>G | p.Y1150C |
| | 18- 1102 | X | 53619434 | 53619434 | C | T | Missense Mutation | c.3896G>A | p.R1299Q |
| | 19- 078 | X | 53581633 | 53581633 | C | T | Missense Mutation | c.8455G>A | p.E2819K |
| *FAT1* | 17- 607 | 4 | 187549491 | 187549491 | - | T | Frame Shift Ins | c.4627_4627insA | p.R1543fs |
| | 17- 641 | 4 | 187629733 | 187629733 | T | C | Missense Mutation | c.1249A>G | p.T417A |
| | 17- 782 | 4 | 187542017 | 187542017 | G | C | Nonsense Mutation | c.5723C>G | p.S1908* |
| | 19- 078 | 4 | 187524565 | 187524565 | C | G | Missense Mutation | c.11115G>C | p.Q3705H |
| | 19- 253 | 4 | 187549883 | 187549883 | C | T | Missense Mutation | c.4358G>A | p.R1453H |
| *MAPK1* | 17- 415 | 22 | 22127164 | 22127164 | C | T | Missense Mutation | c.964G>A | p.E322K |
| | 17- 417 | 22 | 22127164 | 22127164 | C | T | Missense Mutation | c.964G>A | p.E322K |
| | 17- 590 | 22 | 22127164 | 22127164 | C | T | Missense Mutation | c.964G>A | p.E322K |

| | | | | | | | | | |
|---|---|---|---|---|---|---|---|---|---|
| | 19- 078 | 22 | 22127164 | 22127164 | C | T | Missense Mutation | c.964G>A | p.E322K |
| *COL6A6* | 17-652 | 3 | 130284399 | 130284399 | G | A | Missense Mutation | c.1223G>A | p.R408Q |
| | 18-1062 | 3 | 130311541 | 130311541 | G | T | Splice Site | c.4346G>T | p.G1449V |
| | 19-003 | 3 | 130381047 | 130381047 | G | A | Missense Mutation | c.6397G>A | p.A2133T |
| | 19-078 | 3 | 130289890 | 130289890 | A | G | Missense Mutation | c.2630A>G | p.Q877R |
| *SPTA1* | 17- 590 | 1 | 158592846 | 158592846 | C | T | Missense Mutation | c.6047G>A | p.R2016H |
| | 18- 336 | 1 | 158622375 | 158622375 | A | G | Missense Mutation | c.3257T>C | p.L1086S |
| | 18- 354 | 1 | 158612277 | 158612277 | T | G | Missense Mutation | c.4661A>C | p.Q1554P |
| | 18- 636 | 1 | 158589012 | 158589012 | C | T | Splice Site | c.6530G>A | p.R2177K |
| *MED12* | 17- 590 | X | 70357214 | 70357214 | G | A | Missense Mutation | c.5729G>A | p.R1910H |
| | 17- 673 | X | 70339576 | 70339576 | G | A | Missense Mutation | c.245G>A | p.R82H |
| | 18- 2081 | X | 70360490 | 70360490 | C | G | Nonsense Mutation | c.6050C>G | p.S2017* |
| *ASXL1* | 18- 636 | 20 | 31022243 | 31022243 | - | TCAC GTAT CAA | Frame Shift Ins | c.1728_1728insTCACGTAT CAA | p.S577fs |
| | 18- 2302 | 20 | 31022263 | 31022263 | G | A | Nonsense Mutation | c.1748G>A | p.W583* |
| | 19- 253 | 20 | 31022442 | 31022442 | G | - | Frame Shift Deletion | c.1927delG | p.G646fs |
| *APC* | 17- 590 | 5 | 112173738 | 112173738 | C | T | Missense Mutation | c.2447C>T | p.T816I |
| | 18- 1861 | 5 | 112179015 | 112179015 | C | G | Missense Mutation | c.7724C>G | p.S2575C |
| | 19- 078 | 5 | 112175156 | 112175156 | T | C | Missense Mutation | c.3865T>C | p.C1289R |
| *BCOR* | 18- 636 | X | 39921559 | 39921559 | G | C | Missense Mutation | c.4261C>G | p.R1421G |
| | 19- 003 | X | 39923736 | 39923736 | G | A | Nonsense Mutation | c.3355C>T | p.Q1119* |
| | 19- 078 | X | 39913190 | 39913190 | G | A | Missense Mutation | c.4925C>T | p.S1642L |
| *CHD8* | 18- 336 | 14 | 21865986 | 21865986 | C | T | Missense Mutation | c.5047G>A | p.E1683K |
| | 18- 1102 | 14 | 21868692 | 21868692 | G | A | Missense Mutation | c.4450C>T | p.R1484W |
| | 18- 1201 | 14 | 21896304 | 21896304 | C | T | Missense Mutation | c.1325G>A | p.G442E |

| | | | | | | | | | |
|---|---|---|---|---|---|---|---|---|---|
| *HLA-A* | 18- 335 | 6 | 29910797 | 29910797 | G | T | Nonsense Mutation | c.337G>T | p.E113* |
| | 18- 335 | 6 | 29911144 | 29911144 | T | A | Missense Mutation | c.443T>A | p.I148N |
| | 18- 354 | 6 | 29913009 | 29913009 | A | G | Splice Site | NA | NA |
| | 19- 003 | 6 | 29910668 | 29910668 | G | T | Nonsense Mutation | c.208G>T | p.E70* |
| *SHKBP1* | 18- 932 | 19 | 41096310 | 41096310 | G | A | Missense Mutation | c.1750G>A | p.G584S |
| | 19- 253 | 19 | 41094580 | 41094580 | G | A | Missense Mutation | c.1387G>A | p.G463S |
| *LAMA3* | 17-652 | 18 | 21331008 | 21331008 | C | T | Missense Mutation | c.811C>T | p.H271Y |
| | 19-078 | 18 | 21329399 | 21329399 | A | C | Missense Mutation | c.573A>C | p.K191N |
| *MAGI1* | 18-373 | 3 | 65425561 | 65425563 | CTG | - | In Frame Deletion | c.1261_1263delCAG | p.Q421del |
| | 19-078 | 3 | 65456142 | 65456142 | C | T | Missense Mutation | c.775G>A | p.E259K |
| *FREM1* | 19-078 | 9 | 14824025 | 14824025 | G | C | Missense Mutation | c.2167C>G | p.Q723E |
| | 19-078 | 9 | 14859349 | 14859349 | C | T | Missense Mutation | c.463G>A | p.D155N |
| | 19-1236 | 9 | 14756376 | 14756376 | G | C | Missense Mutation | c.5403C>G | p.D1801E |
| *FBXW7* | 19- 253 | 4 | 153247289 | 153247289 | G | A | Missense Mutation | c.1513C>T | p.R505C |
| | 19- 780 | 4 | 153247289 | 153247289 | G | C | Missense Mutation | c.1513C>G | p.R505G |
| *NFE2L2* | 17- 641 | 2 | 178098810 | 178098810 | C | T | Missense Mutation | c.187G>A | p.E63K |
| | 17- 673 | 2 | 178098944 | 178098944 | C | G | Missense Mutation | c.53G>C | p.R18P |
| *MTOR* | 17- 417 | 1 | 11264633 | 11264633 | T | C | Missense Mutation | c.3929A>G | p.Y1310C |
| | 18- 932 | 1 | 11316219 | 11316219 | C | T | Missense Mutation | c.535G>A | p.V179I |
| *PTEN* | 17- 641 | 10 | 89717708 | 89717708 | C | T | Nonsense Mutation | c.733C>T | p.Q245* |
| | 18- 2081 | 10 | 89692923 | 89692923 | G | A | Missense Mutation | c.407G>A | p.C136Y |
| *AKT1* | 18-1062 | 14 | 105246551 | 105246551 | C | T | Missense Mutation | c.49G>A | p.E17K |
| | 19-1236 | 14 | 105246482 | 105246482 | C | G | Missense Mutation | c.118G>C | p.E40Q |
| | 19-1236 | 14 | 105246551 | 105246551 | C | T | Missense Mutation | c.49G>A | p.E17K |
| *ARID1A* | 17- 369 | 1 | 27101621 | 27101621 | A | - | Frame Shift Deletion | c.4252delA | p.I1418fs |

| | | | | | | | | | |
|---|---|---|---|---|---|---|---|---|---|
| *ARID1A* | 17- 736 | 1 | 27057928 | 27057928 | C | T | Nonsense Mutation | c.1636C>T | p.Q546* |
| *ERBB2* | 17- 819 | 17 | 37880220 | 37880220 | T | C | Missense Mutation | c.2264T>C | p.L755S |
| | 19- 974 | 17 | 37868208 | 37868208 | C | T | Missense Mutation | c.929C>T | p.S310F |
| *THBS2* | 18-1102 | 6 | 169637792 | 169637792 | C | T | Missense Mutation | c.1228G>A | p.V410I |
| | 19-1236 | 6 | 169620302 | 169620302 | C | T | Missense Mutation | c.3502G>A | p.E1168K |
| *ZNF750* | 19-253 | 17 | 80789425 | 80789425 | G | C | Nonsense Mutation | c.906C>G | p.Y302* |
| *ZFHX3* | 18- 1102 | 16 | 72831358 | 72831360 | TTG | - | In Frame Deletion | c.5221_5223delCAA | p.Q1741del |
| *CASP8* | 18- 1062 | 2 | 202136339 | 202136339 | G | T | Missense Mutation | c.583G>T | p.D195Y |
| *ZC3H6* | 17-782 | 2 | 113089698 | 113089698 | G | C | Missense Mutation | c.3203G>C | p.G1068A |
| *STK11* | 17- 641 | 19 | 1220629 | 1220629 | C | T | Missense Mutation | c.647C>T | p.S216F |
| *ATXN3* | 17- 417 | 14 | 92549604 | 92549604 | - | A | Splice Site | NA | NA |
| *GH2* | 17-652 | 17 | 61957765 | 61957765 | G | C | Nonsense Mutation | c.570C>G | p.Y190* |
| *RBPJL* | 17-652 | 20 | 43945460 | 43945460 | C | G | Missense Mutation | c.1415C>G | p.S472C |
| *LAMB3* | 19-1236 | 1 | 209791961 | 209791977 | GGCCTCGCTGACCTCCT | - | Frame Shift Deletion | c.2729_2745delAGGAGGTCAGCGAGGCC | p.QEVSEA910fs |
| *TSC1* | 18-1151 | 9 | 135771988 | 135771990 | GCT | - | In Frame Deletion | c.3127_3129delAGC | p.S1043del |
| *GHR* | 17-857 | 5 | 42700027 | 42700027 | G | A | Missense Mutation | c.541G>A | p.A181T |

**Supplementary Table S10: Comparison of the frequency of mutations in the literture-curated driver genes between HPV16-positive CaCx patients of Indian cohort and TCGA-CESC PanCanAtlas Cohort**

| Gene name | Frequency (%) in Indian cohort (N=49) | Frequency (%) in TCGA cohort (N=291) PanCanAtlas |
|---|---|---|
| ***PIK3CA*** | 42.9 | 28.5 |
| *EP300* | 10.2 | 12 |
| ***MAPK1*** | 8.2 | 3.8 |
| ***HLA-A*** | 6.1 | 4.5 |
| ***SHKBP1*** | 4.1 | 1.4 |
| *PTEN* | 4.1 | 7.2 |
| *NFE2L2* | 4.1 | 5.5 |
| *FBXW7* | 4.1 | 12 |
| *ARID1A* | 4.1 | 5.5 |
| *HLA-B* | 2 | 5.2 |
| *CASP8* | 2 | 4.5 |
| *KMT2C* | 14.3 | 18.6 |
| *KMT2D* | 12.2 | 14.1 |
| ***FAT1*** | 10.2 | 7.9 |
| ***MACF1*** | 10.2 | 4.8 |
| ***HUWE1*** | 10.2 | 9.6 |
| ***SPTA1*** | 8.2 | 5.5 |
| *MED12* | 6.1 | 6.2 |
| ***CHD8*** | 6.1 | 3.4 |
| ***BCOR*** | 6.1 | 4.1 |
| ***ATXN3*** | 2 | 0.3 |
| ***ASXL1*** | 6.1 | 1.7 |
| ***APC*** | 6.1 | 4.1 |
| *NOTCH1* | 2 | 6.9 |
| *TP53* | 0 | 7.9 |

***N.B-*** *The genes marked in bold are the SMGs and driver genes that shows more mutation frequency in Indian cohort compared to TCGA-CESC PanCanAtlas cohort*

**Supplementary Table S11: Details of different *PIK3CA* mutations observed in HPV16-positive *PIK3CA* -mutant CaCx patients (n=21)**

| Variant Classification | cDNA Change | Codon Change | Protein Change | No. of patients | Frequency of variants | Protein Domain |
|---|---|---|---|---|---|---|
| Missense | c.1633 G>A | c.(1633-1635) Gag>Aag | p.E545K | 8 | 30.76 | HD |
| Missense | c.1624 G>A | c.(1624-1626) Gaa>Aaa | p.E542K | 8 | 30.76 | HD |
| Missense | c.1634 A>G | c.(1633-1635) gAg>gGg | p.E545G | 2 | 7.69 | HD |
| Missense | c.2176 G>A | c.(2176-2178) Gaa>Aaa | p.E726K | 2 | 7.69 | KD |
| Missense | c.1645 G>T | c.(1645-1647) Gat>Tat | p.D549Y | 1 | 3.84 | HD |
| Missense | c.277 C>T | c.(277-279) Cgg>Tgg | p.R93W | 1 | 3.84 | ABD |
| Missense | c.3140 A>G | c.(3139-3141) cAt>cGt | p.H1047R | 1 | 3.84 | KD |
| Missense | c.1357 G>A | c.(1357-1359) Gaa>Aaa | p.E453K | 1 | 3.84 | C2 |
| Missense | c.3012 G>A | c.(3010-3012) atG>atA | p.M1004I | 1 | 3.84 | KD |
| Missense | c.3049 G>C | c.(3049-3051) Gat>Cat | p.D1017H | 1 | 3.84 | KD |

***N.B:*** *cDNA Change column represents change from reference allele>variant allele; in Codon Change column, the change in allele is represented in capital letter; Protein Change column represents the change in amino acid in protein sequence; Protein Domain column maps the PIK3CA mutations across different funtional domains of* PIK3CA *protein: HD- Helical Domain, KD- Kinase Domain, ABD- Adaptor-Binding Domain, C2 domain*

**Supplementary Table S12: Degree of connectivity of literature-curated driver genes in protein-protein interaction network based on STRING database**

| Gene name | Degree of connectivity |
|---|---|
| *PIK3CA* | 19 |
| *PTEN* | 17 |
| *EP300* | 14 |
| *ERBB2* | 14 |
| *KMT2D* | 14 |
| *ARID1A* | 13 |
| *AKT1* | 13 |
| *MAPK1* | 11 |
| *KMT2C* | 11 |
| *MTOR* | 11 |
| *STK11* | 10 |
| *NFE2L2* | 9 |
| *CASP8* | 8 |
| *MED12* | 8 |
| *FAT1* | 8 |
| *TSC1* | 7 |
| *BCOR* | 6 |
| *APC* | 5 |
| *HUWE1* | 5 |
| *ASXL1* | 4 |
| *GH2* | 3 |
| *MACF1* | 3 |
| *HLA-A* | 3 |
| *GHR* | 3 |
| *ZNF750* | 2 |
| *LAMA3* | 2 |
| *COL6A6* | 2 |
| *MAGI1* | 2 |
| *SHKBP1* | 1 |
| *RBPJL* | 1 |
| *THBS2* | 1 |
| *LAMB3* | 1 |
| *FREM1* | 1 |
| *PCDHGA12* | 0 |
| *PSPC1* | 0 |
| *SLC35G5* | 0 |
| *ZC3H6* | 0 |
| *PCDHA9* | 0 |

**Supplementary Table S13: Comparison of the clinical characteristics between *PIK3CA* -mutant (n=21) and *PIK3CA* -wild (n=28) tumors in HPV16-positive CaCx cohort**

| Characteristics | *PIK3CA* -Mutant tumors (n=21) | *PIK3CA* -wild tumors (n=28) | p value |
|---|---|---|---|
| **Age at diagnosis** | 55.19±9.44 | 56.04±13.47 | 0.8 |
| **FIGO Stage** | | | 0.843 |
| Early (≤IIA) | 5 | 6 | |
| Late (≥ IIB) | 16 | 22 | |
| **Lymph node involvement** | | | 0.145 |
| Involved (pN+) | 13 | 11 | |
| Not Involved (pN-) | 8 | 16 | |

***N.B:*** *For age, mean age ±SD was calculated for PIK3CA-mutant and PIK3CA-wild tumors; and p-value was calculated using the t-test of equality of means. For rest of the clinical parameters, Chi-square test of significance was performed to detect the p value.*

**Supplementary Table S14: Details of the *PIK3CA* mutation status in HPV16-positive TCGA-CESC patients profiled for Whole Exome Sequencing (n=103)**

| Sample ID | *PIK3CA* status | PIK3CA Mutation site | FIGO stage |
|---|---|---|---|
| TCGA-C5-A1BE | Mutated | p.E726K | Stage IIA2 |
| TCGA-C5-A1BJ | Mutated | p.E545Q, p.E600K | Stage IB1 |
| TCGA-C5-A1MH | Mutated | p.E545K, p.E726K | Stage IIB |
| TCGA-C5-A1ML | Mutated | p.E545K | Stage IIB |
| TCGA-C5-A2M1 | Mutated | p.E545K | Stage IIB |
| TCGA-C5-A7CJ | Mutated | p.E542K | Stage IB1 |
| TCGA-DG-A2KK | Mutated | p.E545K | Stage IB1 |
| TCGA-DG-A2KL | Mutated | p.E542K | Stage IIB |
| TCGA-DS-A5RQ | Mutated | p.E545K | Stage IB2 |
| TCGA-EA-A3HT | Mutated | p.E545K | Stage IIB |
| TCGA-EA-A3QE | Mutated | p.H1047R | Stage IVB |
| TCGA-EA-A50E | Mutated | p.E545K | Stage IVB |
| TCGA-EA-A5FO | Mutated | p.E545K | Stage IA2 |
| TCGA-EK-A2RN | Mutated | p.E545K ,p.E726K | Stage IB1 |
| TCGA-EK-A2RO | Mutated | p.E545K | Stage IB1 |
| TCGA-FU-A3HY | Mutated | p.H1047L | Stage IIA2 |
| TCGA-FU-A3WB | Mutated | p.E545K | Stage IB2 |
| TCGA-HG-A2PA | Mutated | p.G106V | Stage IB |
| TCGA-IR-A3LF | Mutated | p.E542K, p.G1007R | Stage II |
| TCGA-IR-A3LI | Mutated | p.E545K | Stage IB1 |
| TCGA-JW-A5VL | Mutated | p.E545K | Stage IIB |
| TCGA-JW-A69B | Mutated | p.E542K | Stage IIIB |
| TCGA-LP-A7HU | Mutated | p.E542K, p.E545K | Stage IVB |
| TCGA-MY-A5BD | Mutated | p.E542K | Stage IB |
| TCGA-MY-A5BE | Mutated | p.E545K | Stage IIB |
| TCGA-Q1-A5R1 | Mutated | p.E545K | Stage IB1 |
| TCGA-Q1-A6DV | Mutated | p.G118D | Stage IB |
| TCGA-BI-A0VR | Wild | | Stage IB1 |
| TCGA-BI-A0VS | Wild | | Stage IIIB |
| TCGA-BI-A20A | Wild | | Stage IIA2 |
| TCGA-C5-A0TN | Wild | | Stage IVA |
| TCGA-C5-A1BI | Wild | | Stage IB1 |
| TCGA-C5-A1BK | Wild | | Stage IIIB |
| TCGA-C5-A1BL | Wild | | Stage IIB |
| TCGA-C5-A1M8 | Wild | | Stage IIIB |

| | | | |
|---|---|---|---|
| TCGA-C5-A1M9 | Wild | | Stage IB1 |
| TCGA-C5-A1MN | Wild | | Stage IB |
| TCGA-C5-A1MP | Wild | | Stage IB2 |
| TCGA-C5-A2LS | Wild | | Stage IVA |
| TCGA-C5-A2LV | Wild | | Stage IIB |
| TCGA-C5-A2LX | Wild | | Stage IB2 |
| TCGA-C5-A2LY | Wild | | Stage IB1 |
| TCGA-C5-A2LZ | Wild | | Stage IB1 |
| TCGA-C5-A2M2 | Wild | | Stage IIB |
| TCGA-C5-A3HD | Wild | | Stage IIA |
| TCGA-C5-A3HL | Wild | | Stage IVB |
| TCGA-C5-A7CH | Wild | | Stage IB1 |
| TCGA-C5-A7CK | Wild | | Stage IIB |
| TCGA-C5-A7CL | Wild | | Stage IIIB |
| TCGA-C5-A7UE | Wild | | Stage IB1 |
| TCGA-DG-A2KM | Wild | | Stage IA |
| TCGA-DR-A0ZL | Wild | | Stage IIB |
| TCGA-DS-A0VK | Wild | | Stage IB1 |
| TCGA-DS-A0VL | Wild | | Stage IB2 |
| TCGA-DS-A0VM | Wild | | Stage IVB |
| TCGA-DS-A0VN | Wild | | Stage IIB |
| TCGA-EA-A1QT | Wild | | Stage IIIB |
| TCGA-EA-A3HQ | Wild | | Stage IIIB |
| TCGA-EA-A3HU | Wild | | Stage IB1 |
| TCGA-EA-A3QD | Wild | | Stage IB2 |
| TCGA-EA-A411 | Wild | | Stage IB1 |
| TCGA-EA-A43B | Wild | | Stage IB1 |
| TCGA-EA-A44S | Wild | | Stage IIIB |
| TCGA-EA-A5O9 | Wild | | Stage IB1 |
| TCGA-EK-A2H1 | Wild | | Stage IB |
| TCGA-EK-A2IP | Wild | | Stage IIIA |
| TCGA-EK-A2PI | Wild | | Stage IVA |
| TCGA-EK-A2RB | Wild | | Stage IB1 |
| TCGA-EK-A2RC | Wild | | Stage II |
| TCGA-EK-A2RE | Wild | | Stage IB |
| TCGA-EK-A2RK | Wild | | Stage IB |
| TCGA-EK-A2RL | Wild | | Stage IB1 |
| TCGA-EK-A3GK | Wild | | Stage IIB |
| TCGA-EK-A3GN | Wild | | Stage IVB |
| TCGA-EX-A1H5 | Wild | | Stage IIIB |
| TCGA-EX-A1H6 | Wild | | Stage IIIB |

| | | | |
|---|---|---|---|
| TCGA-EX-A69L | Wild | | Stage IIIB |
| TCGA-FU-A2QG | Wild | | Stage IIB |
| TCGA-FU-A3EO | Wild | | Stage IB2 |
| TCGA-FU-A3NI | Wild | | Stage IB1 |
| TCGA-FU-A3TQ | Wild | | Stage IB1 |
| TCGA-FU-A3YQ | Wild | | Stage IB1 |
| TCGA-FU-A40J | Wild | | Stage IIIB |
| TCGA-FU-A770 | Wild | | Stage IB |
| TCGA-HM-A3JJ | Wild | | Stage IB2 |
| TCGA-HM-A4S6 | Wild | | Stage IIIB |
| TCGA-IR-A3LC | Wild | | Stage II |
| TCGA-IR-A3LL | Wild | | Stage IB2 |
| TCGA-JW-A5VK | Wild | | Stage I |
| TCGA-JX-A3Q8 | Wild | | Stage IB2 |
| TCGA-JX-A5QV | Wild | | Stage IIA |
| TCGA-LP-A4AW | Wild | | '-- |
| TCGA-LP-A4AX | Wild | | Stage IB1 |
| TCGA-LP-A5U2 | Wild | | Stage II |
| TCGA-LP-A5U3 | Wild | | Stage IB2 |
| TCGA-MU-A51Y | Wild | | Stage IIIB |
| TCGA-MU-A5YI | Wild | | Stage IB1 |
| TCGA-MY-A5BF | Wild | | Stage IB1 |
| TCGA-Q1-A5R2 | Wild | | Stage I |
| TCGA-Q1-A6DT | Wild | | Stage IIIB |
| TCGA-Q1-A6DW | Wild | | Stage IB |
| TCGA-Q1-A73R | Wild | | Stage IIIB |
| TCGA-RA-A741 | Wild | | Stage IB1 |

**Supplementary Table S15: Association of clinical parameters with PIK3CA-mutant (n=27) and PIK3CA-wild tumors (n=76) in HPV16-positive TCGA-CESC cohort**

| Characteristics | PIK3CA-Mutant tumors (n=27) | PIK3CA-wild tumors (n=76) | p value |
|---|---|---|---|
| **Age at diagnosis** | 49.59±15.47 | 44.23±11.62 | 0.06 |
| **FIGO Stage** | | | 0.946 |
| Early (≤IIA) | 16 | 45 | |
| Late (≥ IIB) | 11 | 30 | |
| **Lymph node involvement** | | | 0.041 |
| Involved (pN+) | 3 | 23 | |
| Not Involved (pN-) | 15 | 30 | |

***N.B*** *. For age, mean age ±SD was calculated for PIK3CA-mutant and PIK3CA-wild tumors; p-value was calculated using the t-test of equality of means. While for rest of the clinical parameters, Chi-square test of significance was performed to detect the p value.*

**Supplementary Table S16: List of Differentially Expressed Genes (DEGs) in *PIK3CA* -mutant tumors (n=17) compared to *PIK3CA* -wild tumors (n=12) in Indian HPV16-positive CaCx cohort, portraying expression difference of fold change cut-off |log2(FC) | > 0.6 and False Discovery Rate (FDR) corrected p<0.05**

| Gene Name | log2FC | FDR | Status in *PIK3CA* -mutant compared to *PIK3CA* -wild tumors |
|---|---|---|---|
| *MARCO* | 2.966 | 0.038 | Upregulation |
| *STAG3* | 2.382 | 0.035 | Upregulation |
| *PGC* | 4.684 | 0.01 | Upregulation |
| *CNTD2* | 2.24 | 0.018 | Upregulation |
| *THBS4* | 1.553 | 0.014 | Upregulation |
| *ELF5* | 3.406 | 0.024 | Upregulation |
| *FBXO10* | 0.823 | 0.019 | Upregulation |
| *GAL3ST2* | 2.994 | 0.01 | Upregulation |
| *HOXB13* | 3.713 | 0.01 | Upregulation |
| *AZGP1* | 3.565 | 0.014 | Upregulation |
| *TDRD12* | 3.501 | 0.04 | Upregulation |
| *LINC00471* | 1.272 | 0.01 | Upregulation |
| *PRAME* | 3.353 | 0.04 | Upregulation |
| *HEPACAM2* | 3.276 | 0.025 | Upregulation |
| *PNMA5* | 4.12 | 0.04 | Upregulation |
| *PRAC2* | 3.811 | 0.038 | Upregulation |
| *SPESP1* | 2.957 | 0.04 | Upregulation |
| *RP1-232P20.1* | 3.652 | 0.018 | Upregulation |
| *AFAP1-AS1* | 4.41 | 0.001 | Upregulation |
| *PTHLH* | -2.396 | 0.037 | Downregulation |
| *ADAMTS19* | -3.931 | 0.041 | Downregulation |
| *SOST* | -3.078 | 0.04 | Downregulation |
| *FABP4* | -4.868 | 0 | Downregulation |
| *PNLIPRP3* | -3.584 | 0.035 | Downregulation |
| *PWRN1* | -4.799 | 0.003 | Downregulation |
| *PWRN3* | -4.188 | 0.024 | Downregulation |
| *RP11-680G10.1* | -2.112 | 0.024 | Downregulation |
