## Supplementary material for "Integrated Genomic and Immune Profiling Reveals PIK3CA-Mutant HPV16-Positive Cervical Cancer as a Clinically Aggressive and Immunologically Distinct Subtype in Patients from Eastern India": Supplemtary Figures

**Supplementary Figure S1**

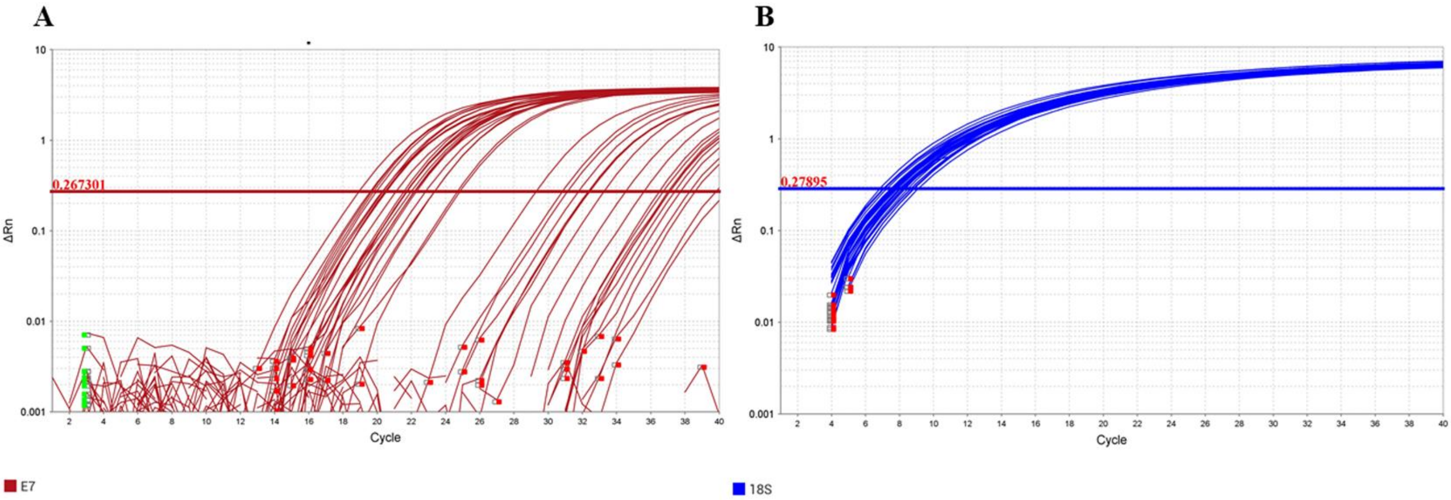


**Supplementary Figure S1:** Amplification plot of **(A)** viral oncogene HPV16 E7 and **(B)** 18srRNA (house-keeping gene) by TaqMan-based qRT-PCR

**Supplementary Figure S2**

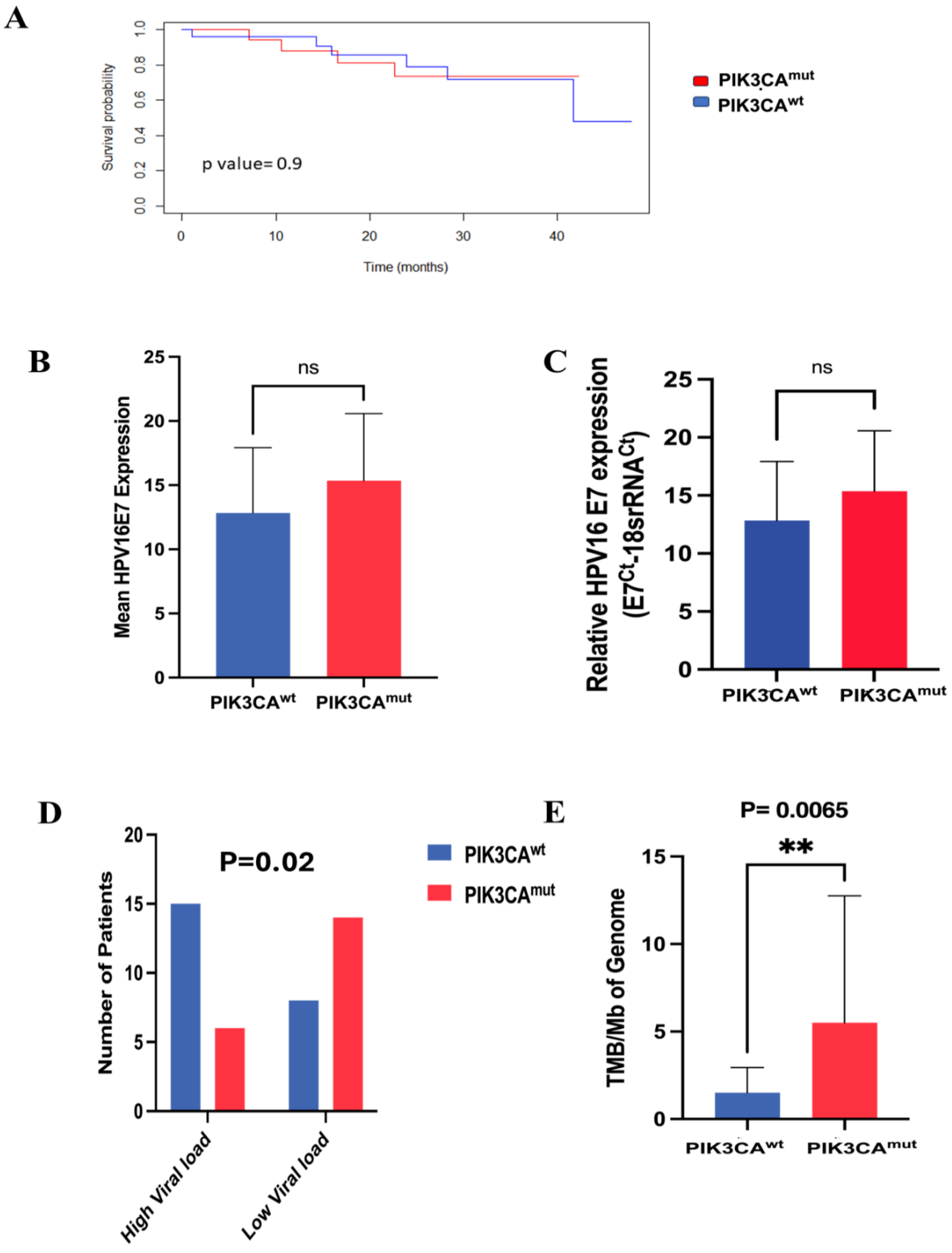

**Supplementary Figure S2: Association of clinicopathological features and viral factors with *PIK3CA* mutation in our cohort of HPV16 positive CaCx patients. (A)** Kaplan Meier plot depicting the overall survival (OS) of the HPV16 CaCx patients with (n=21) and without (n=28) mutations in *PIK3CA* gene (PIK3CA$^{mut}$ vs PIK3CA$^{wt.}$) The Y axis represents the probability of OS of patients belonging to different groups and X axis represents the time of survival (in months). **(B)** Box plot representing mean expression (based on TPM values of RNA-seq) and **(C)** Relative expression (based on qRT-PCR) of viral oncoprotein HPV16 E7 in PIK3CA$^{mut}$ vs PIK3CA$^{wt}$ tumors. Relative gene expression reflects expression of E7 minus expression of the house-keeping gene (18srRNA). The data is represented as mean ± SD. **(D)** Distribution of HPV16 viral load (natural log transformed) and **(E)** total TMB/Mb of the genome in PIK3CA$^{mut}$ vs PIK3CA$^{wt}$ tumors.

"**" represents significant p value < 0.01 (two-tailed Students' t-test)

TMB: Tumor Mutation Burden

PIK3CA$^{mut:}$ *PIK3CA*-mutant tumors

PIK3CA$^{wt}$: *PIK3CA*-wild tumors

**Supplementary Figure S3**

**A**

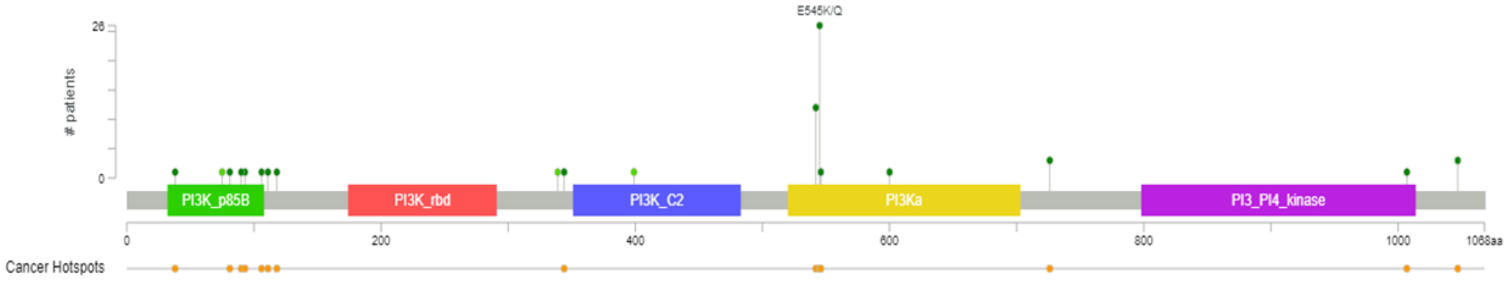


**B**

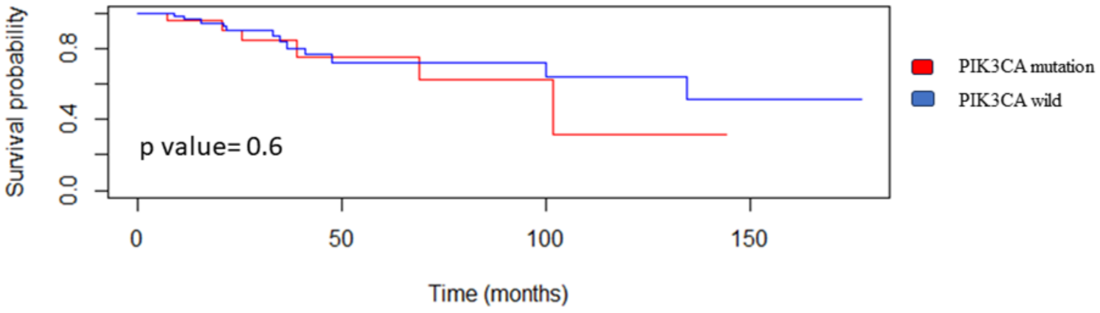


**Supplementary Figure S3: *PIK3CA* mutational landscape and its association with overall survival (OS) in HPV16-positive TCGA-CESC patients (A)** Lollipop plot representing the overview of mutations in *PIK3CA* in HPV16 positive TCGA-CESC cohort**.** *The plot was downloaded from cBioportal*. **(B)** Kaplan-Meier plot depicting the patient OS of patient cohort with and without mutations in *PIK3CA* gene**.** The Y axis represents the probability of OS of patients belonging to different groups and X axis represents the time of survival (in months).

**Supplementary Figure S4**

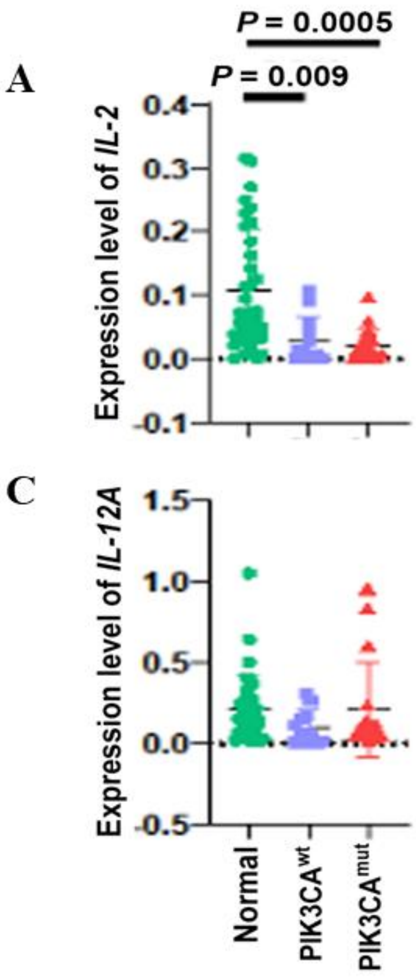


**Supplementary Figure S4: Differential expression of immune-related genes based on *PIK3CA* mutation status:** Scatter dot plots portraying the expression levels (TPM) of **(A)** *IL2*, **(B)** *CD40LG*, **(C)** *IL24*, and **(D)** *IL12B* in normal cervical tissues (Normal), *PIK3CA*-wild tumors ($PIK3CA^{wt}$), and *PIK3CA*-mutant tumors ($PIK3CA^{mut}$). Statistically significant differences between groups are indicated by the corresponding p values above the plots. P (t-test) < 0.05 considered significant.

**Supplementary Figure S5**

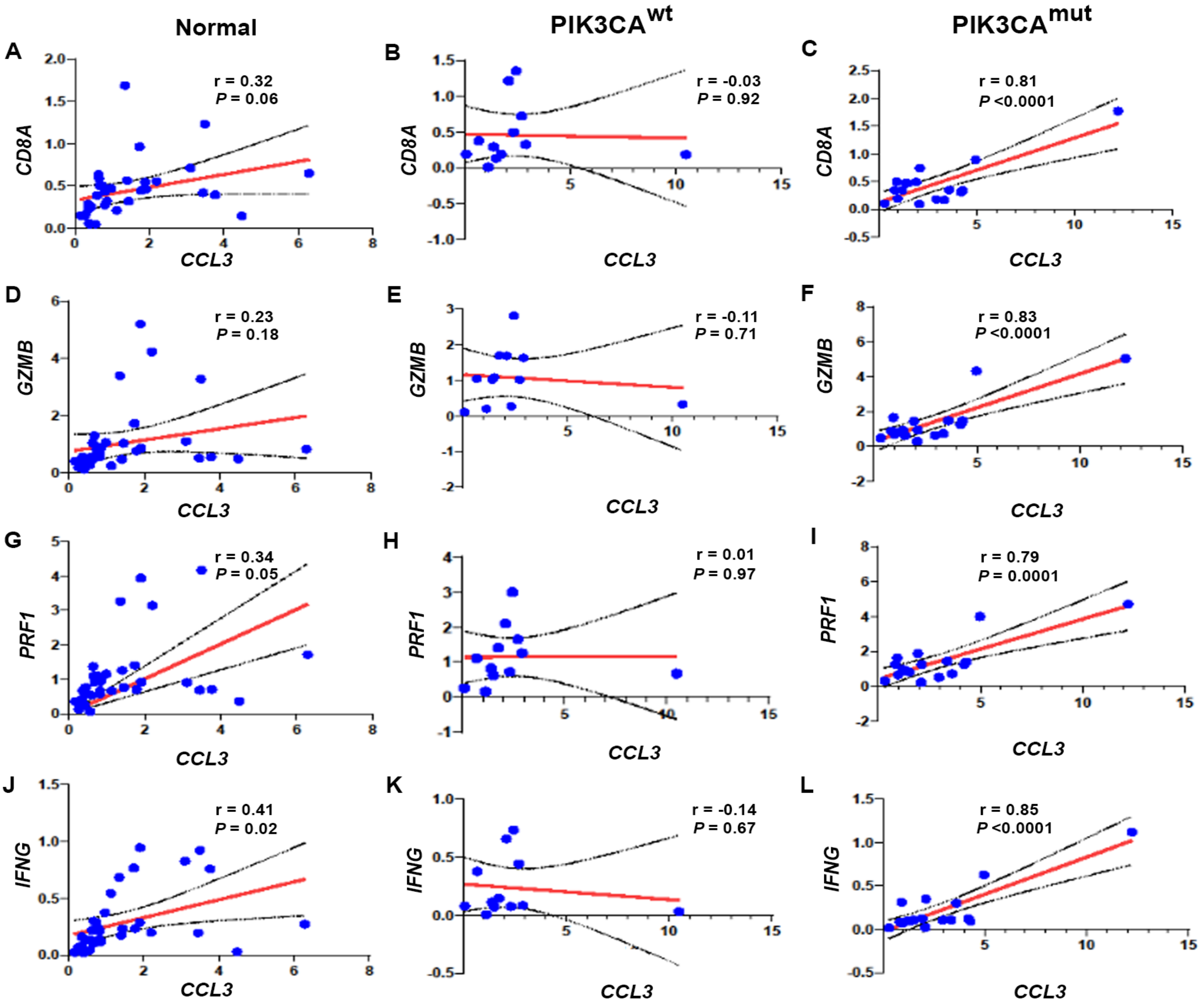

**Supplementary Figure S5: Sample-wise correlation plots depicting the correlation between expression levels (TPM) of *CCL3* with cytotoxic T-cell markers: (A–C)** *CD8A*, **(D-F)** *GZMB*, **(G–I)** *PRF1* and **(J–L)** *IFNG* in normal cervical tissues (Normal), *PIK3CA*-wild tumors ($PIK3CA^{wt}$), and *PIK3CA*-mutant tumors ($PIK3CA^{mut}$) respectively. The red line indicates the linear regression fit, while the black line represents the 95% confidence interval. Correlation coefficients (*r*) and corresponding *p* values were calculated using Pearson's correlation analysis.

**Supplementary Figure S6**

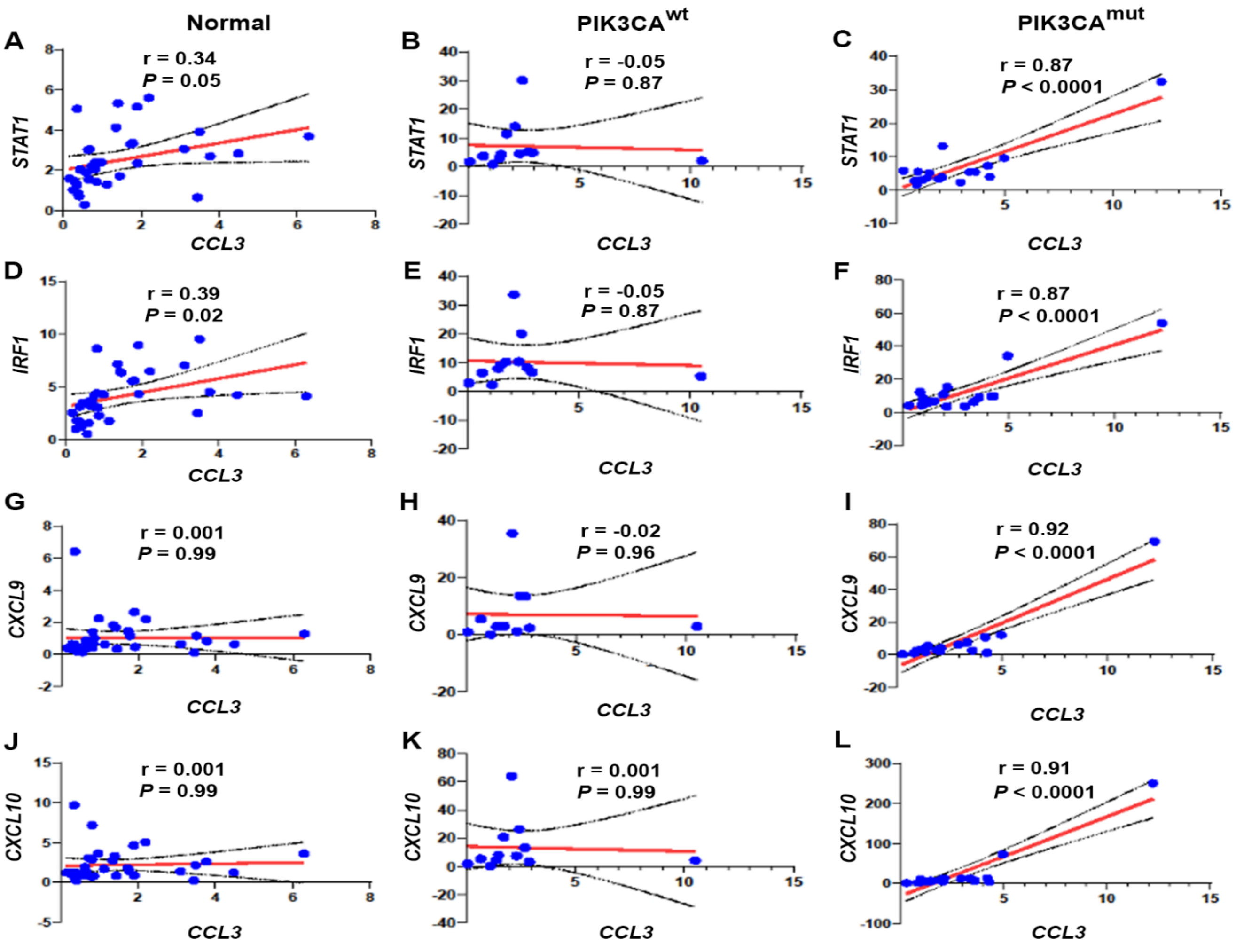


**Supplementary Figure S6: Sample-wise correlation plots depicting the correlation between expression levels (TPM) of *CCL3* with interferon- γ–related immune signatures: (A–C)** *STAT1*, **(D–F)** *IRF1*, **(G–I)** *CXCL9*, and **(J–L)** *CXCL10* in normal cervical tissues (Normal), *PIK3CA*-wild tumors (PIK3CA$^{wt}$), and *PIK3CA*-mutant tumors (PIK3CA$^{mut}$) respectively in each panel. The red line indicates the linear regression fit, while the black line represents the 95% confidence interval. Correlation coefficients (*r*) and corresponding *p* values were calculated using Pearson's correlation analysis.

**Supplementary Figure S7**

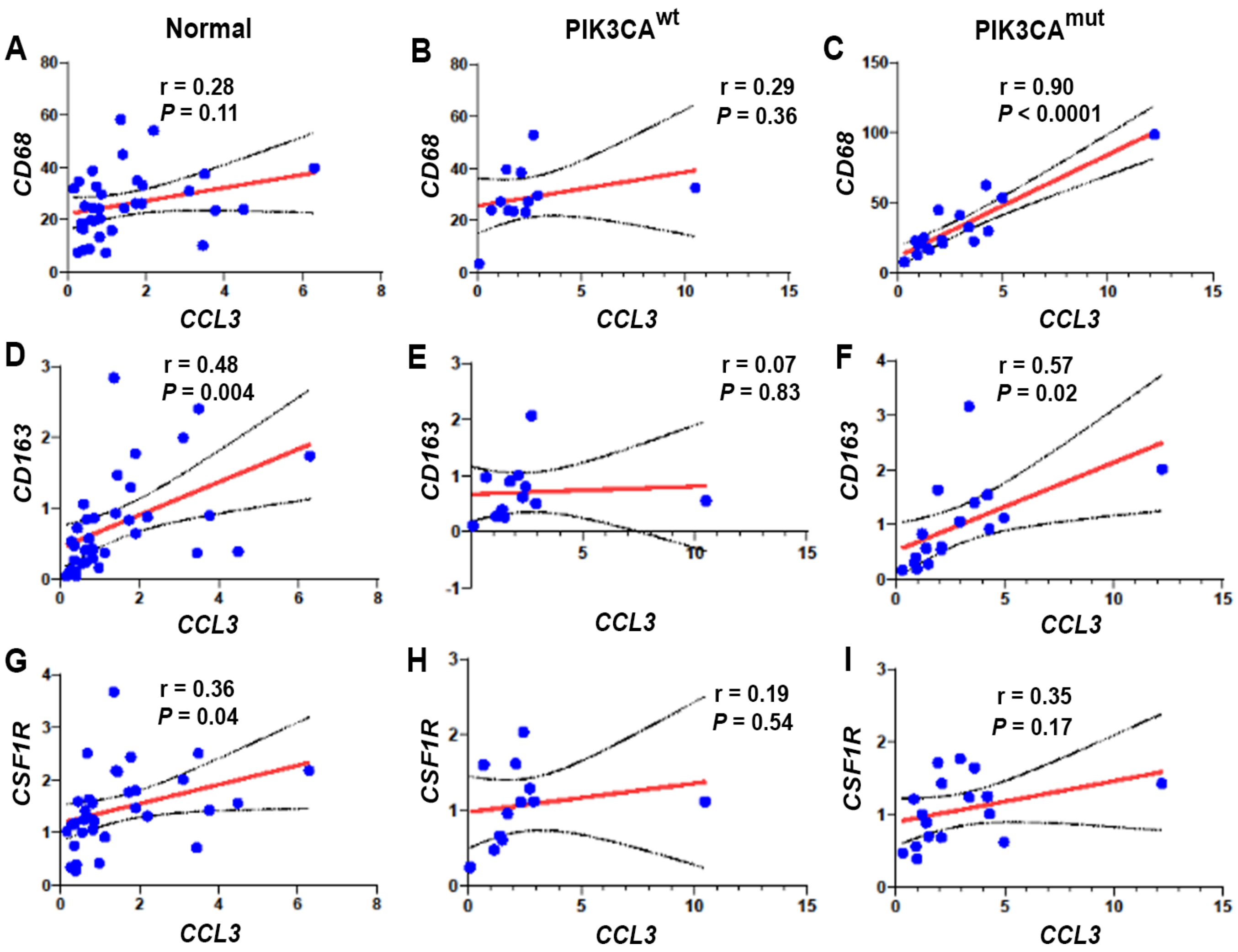


**Supplementary Figure S7: Sample-wise correlation plots depicting the correlation between expression levels (TPM) of *CCL3* with myeloid lineage markers: (A–C)** *CD68*, **(D–F)** *CD163*, and **(G–I)** *CSF1R* in normal cervical tissues (Normal), *PIK3CA*-wild tumors ($PIK3CA^{wt}$), and *PIK3CA*-mutant tumors ($PIK3CA^{mut}$) respectively in each panel. The red line indicates the linear regression fit, while the black line represents the 95% confidence interval. Correlation coefficients (*r*) and corresponding *p* values were calculated using Pearson's correlation analysis.

**Supplementary Figure S8**

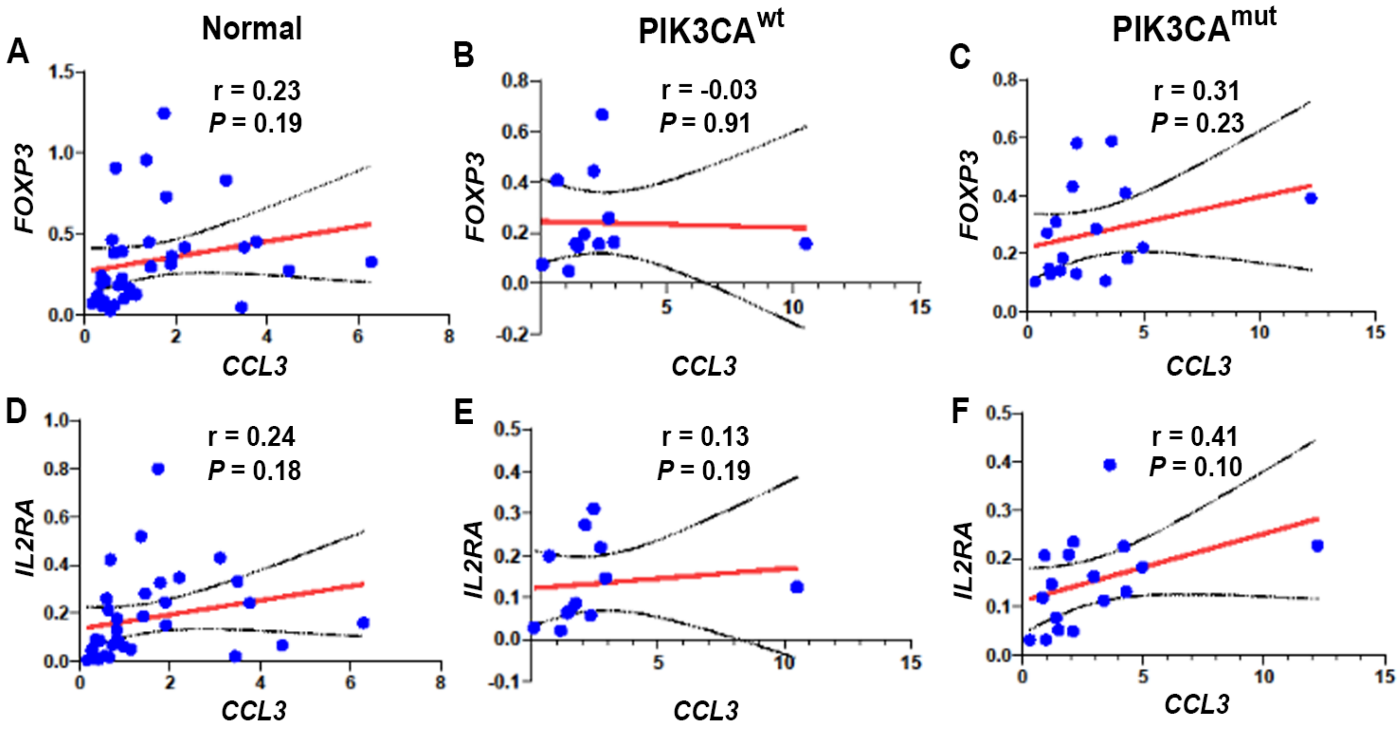


**Supplementary Figure S8: Sample-wise correlation plots depicting the correlation between expression levels (TPM) of *CCL3* with Treg-associated markers: (A–C)** *FOXP3* and **(D–F)** *IL2RA* in normal cervical tissues (Normal), *PIK3CA*-wild tumors (PIK3CA$^{wt}$), and *PIK3CA*-mutant tumors (PIK3CA$^{mut}$) respectively in each panel. The red line indicates the linear regression fit, while the black line represents the 95% confidence interval. Correlation coefficients (*r*) and corresponding *p* values were calculated using Pearson's correlation analysis.
